# Long-term neurological and gastrointestinal sequelae of SARS-CoV-2, influenza, and other neurotropic infections with and without vaccination

**DOI:** 10.1101/2024.07.25.24310990

**Authors:** Bo Konings, Luisa Villatoro, Robert Burns, Guillermo Barahona, Megan McKnight, Ken Hui, Jan Tack, Pankaj Jay Pasricha

## Abstract

COVID-19 increases the risk of neurological and gastrointestinal sequelae, but it is unclear if it does so more than other infections. Using a multicenter record network, we matched 649,478 COVID-19 patients to negative controls (NCs) and patients infected with influenza, human herpesvirusses, and lyme’s disease (LD) to compare new-onset gastrointestinal (GISx), autonomic (ANSx), sensory (SNSx), and motor (MNSx) symptoms 3-12 months after infection. ANSx showed significant increases compared to NCs (odds ratio (OR) 1.34; confidence interval (CI) 1.31-1.36) and most other investigated infections (LD, influenza, infectious mononucleosis, and herpes zoster; OR 1.40, 1.13, 1.11, and 1.05, respectively). SNSx (OR 1.35; CI 1.31-1.39), MNSx (OR 1.32; CI 1.28-1.36) and GISx (OR 1.36; CI 1.33-1.38) were increased but varied more compared with other infections. COVID-19 vaccination reduced the risk of GISx, ANSx, and SNSx. Sequelae frequently ascribed to COVID-19 may manifest with similar or higher frequency after other infections, except ANSx.

## INTRODUCTION

As the urgent challenges posed by the global COVID-19 pandemic are beginning to attenuate, attention is beginning to shift to the post-acute sequelae of SARS-CoV-2 (PASC), more commonly referred to as ‘Long COVID’.^1^ Even when relying on the most conservative estimates, about 10% of individuals infected by COVID-19 may be affected.^2^ Symptoms that have been shown to originate or persist months after the infection include gastrointestinal (GI) symptoms such as constipation, gastro-esophageal reflux disease, abdominal pain, and nausea, as well as neurological symptoms such as brain fog, cognitive dysfunction, sensorimotor symptoms, and neuralgia.^2^

Possible causes for these symptoms include neuronal dysfunction affecting both the central nervous system (CNS) and various divisions of the peripheral nervous system (PNS), including the autonomic (ANS), enteric (ENS) somatosensory (SNS) and motor (MNS) nervous systems, ascribed variously to autoimmunity, microbiota dysbiosis, or vascular abnormalities.^3^ Much of the groundwork on Long-COVID has been established by research about well-known pathogens with significant phenotypical and mechanistic similarities to COVID-19.^3^ Animal studies have shown that SARS-CoV and MERS-CoV demonstrate neuroinvasive properties by entering the brain through olfactory pathways and spreading to thalamic and brainstem neurons.^4^ In humans, neuropathological studies have established indirect evidence of CNS invasion by detecting SARS-CoV-2 RNA and proteins in the brain.^5^ In the PNS, motor-predominant polyneuropathies and myopathies have been described in the post-acute phase of SARS infections.^6^ However, long-lasting systemic effects of infections are by no means a new concept. Viruses from the Human Herpes Virus family (HHV) have been shown to exploit the vicinity of neural tissue in the respiratory and olfactory tract to invade almost any cell type of the nervous system, displaying distinct affinity for the sensory ganglia.^7,8^ Another example is the influenza virus, which has the potential to cause serious neurological complications, especially in children.^9^ Finally, Lyme’s disease associated neuroborreliosis is a classic, yet controversial, example of long-lasting neurological complications arising after a bacterial infection.^10^

Evidence overwhelmingly indicates that COVID-19 has the potential to directly or indirectly affect the CNS and PNS, and the often overlooked second largest collection of neurons in the body, the ENS.^11^ However, no large-scale studies have been performed to directly compare rates of long-term sequelae of COVID-19 to those of other pathogens with established neurotropic properties. Using a nationwide electronic health record (EHR) network, we set up a retrospective cohort-study design to compare new-onset GI symptoms and diagnoses (GISx), and symptoms referrable to the autonomic (ANSx), sensory (SNSx) and motor nervous systems (MNSx) 3 to 12 months after a first diagnosis of COVID-19 to contemporary negative controls (NCs) and patients diagnosed with Influenza, Lyme’s disease (LD), or several HHV, including Cytomegalovirus (CMV), Herpes Zoster (HZ; “shingles”), Varicella (VZ; “chickenpox”), and Infectious Mononucleosis (IM). We also evaluated the significance of anosmia in predicting these sequelae, as this symptom has been suggested to be a harbinger of neurological and GI sequelae.^12^ Finally, we examined the effect of vaccination on the development of outcomes following COVID-19 and influenza.

## RESULTS

### Study population

A total of 649,478 patients with COVID-19 met all selection criteria (**Table 1**). Baseline characteristics for each control population before and after matching are presented in **Supplementary Tables 1-5**.

**Table 1.**
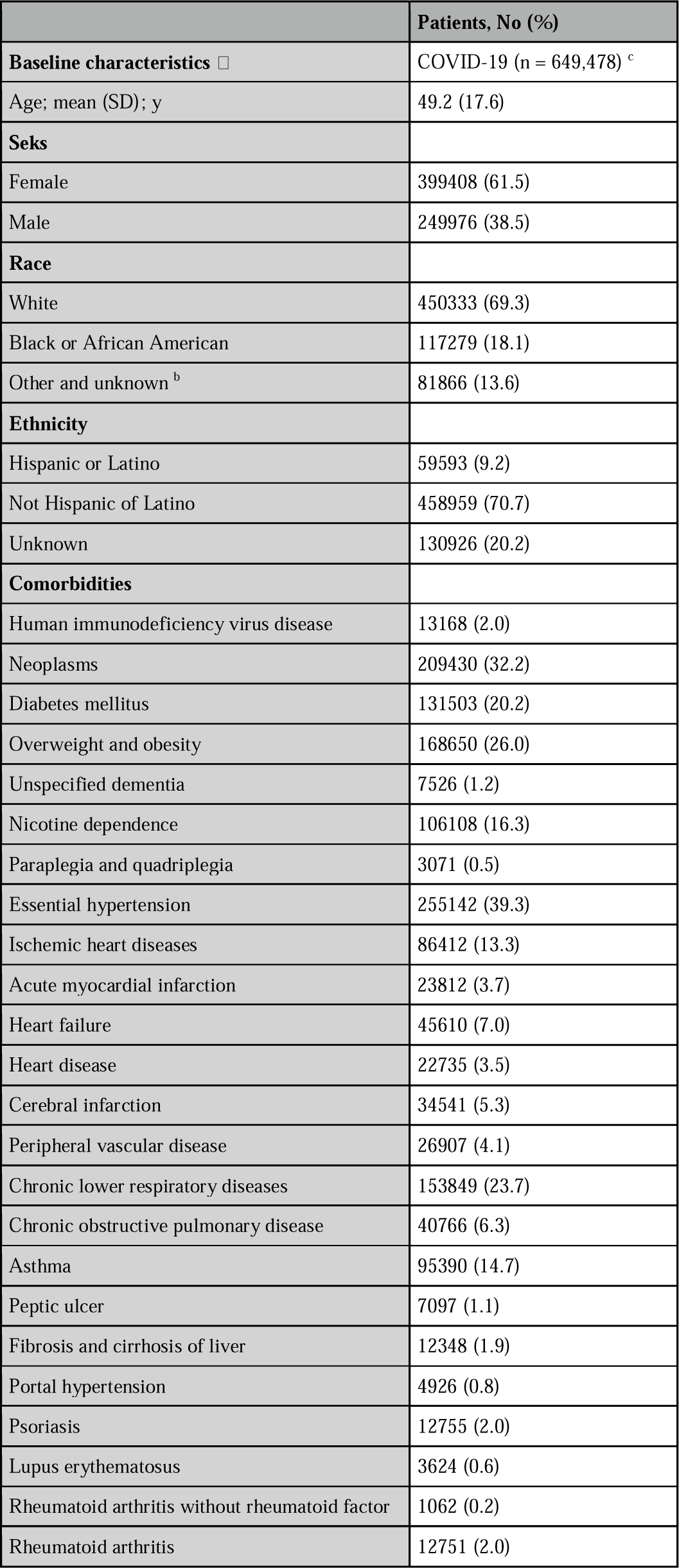
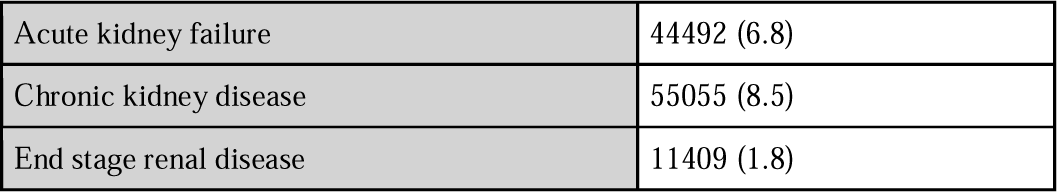
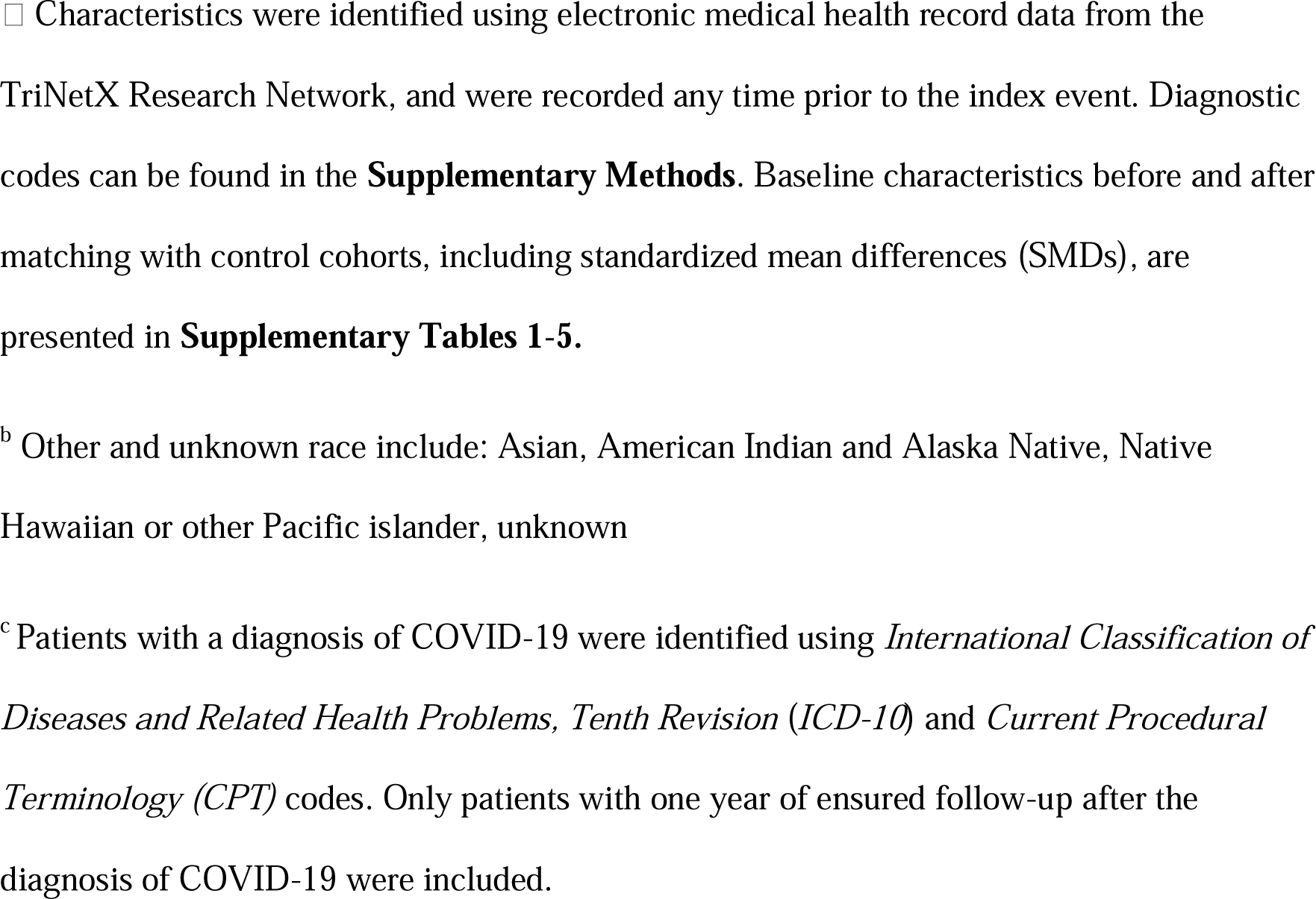
Baseline characteristics of the COVID-19 cohort.

### Neurological and gastrointestinal sequelae

**Figure 1** and **Supplementary Tables 6-9** present the ORs and absolute rates of new-onset primary outcomes after COVID-19 compared to matched contemporary NCs and other infectious diseases. ANSx were significantly (P<0.05) more prevalent after COVID-19 compared with NCs (8.5% vs 6.51; OR 1.34; CI [1.31-1.36]), influenza (OR 1.13; CI [1.11-1.16]), LD (OR 1.21; CI [1.16 – 1.27]), HZ (OR 1.05; CI [1.03-1.08]), and IM (OR 1.11; CI [1.03-1.18]; all P<0.05). No significant differences were observed compared to VZ and CMV (**Fig. 1**). SNSx and MNSx were increased after COVID-19 compared to NCs (OR 1·35 and 1·32; both P<·0001, respectively). However, a diagnosis of influenza (OR 0.84; P<.0001; and 0.94; P 0.004, respectively), HZ (OR 0.92 and 0.88; both P<.0001, respectively), and VZ (OR 0.49 and 0.60; both P<.0001, respectively) showed an even greater risk of SNSx and MNSx. No differences were observed compared to LD, IM, and CMV (**Fig. 1**). The risk of developing any new-onset GISx was significantly greater after COVID-19 compared to contemporary NCs (10.62% vs 8.04%; OR 1.36; CI [1.33-1.38]), LD (1.4 [1.34-1.46]), and VZ (1.17 [1.10-1.24]). No significant differences in the overall prevalence of GISx were observed compared to influenza, HZ, IM, and CMV (**Fig. 1**).

**Figure 1.**
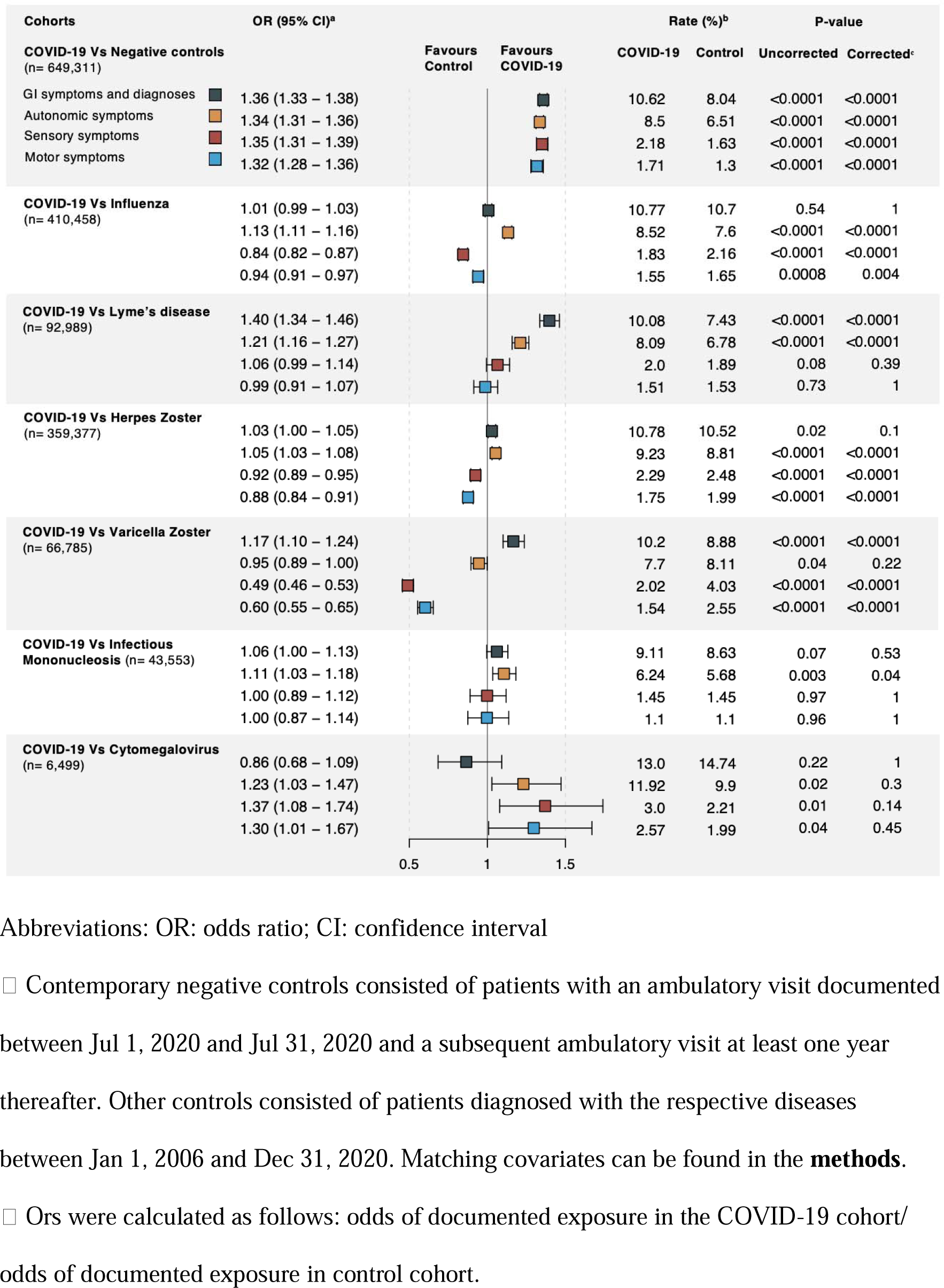
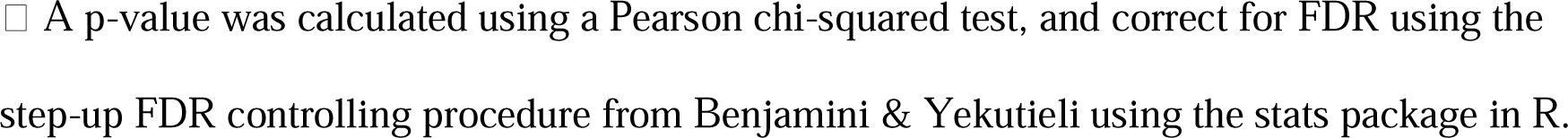
ORs and rates of new-onset outcomes between 3 months and one year after COVID-19, compared to matched contemporary NCs and other infectious diseases.

Individual symptoms after COVID-19 compared to contemporary NCs are presented in **Figure 2**. The increase in ANSx predominantly resulted from an increase in postural symptoms (6.07% vs 4.34%; OR 1.43; CI [1.40-1.45]), urinary symptoms (2.73% vs 2.08%; OR 1.32; CI [1.29-1.35]), and exocrine gland dysfunction (1.88% vs 1.19%; OR 1.58; CI [1.54-1.63]). A significant increase in prescriptions often used for the treatment of ANSx (4.15% vs 3.55%; OR 1.17; CI [1.15-1.20]) provided internal validation. The most prevalent GI symptoms after COVID-19 as compared to NCs were abdominal and pelvic pain (6.43%), diarrhea (3.09%), nausea (2.97%) and constipation (2.91%). The most prevalent ICD-10 GI diagnosis was gastro-esophageal reflux disease (GERD; 3.93%), followed by inflammatory bowel disease (IBD; 1%), functional dyspepsia (FD; 0.34%) and gastroparesis (GP; 0.16%; all P<.0001); a diagnosis of irritable bowel syndrome (IBS) was not increased (0.48% vs 0.46%; OR 1.05; CI [1-1.11]). Although the overall prevalence of new-onset GI symptoms did not differ between COVID-19 and influenza, the risk of developing abdominal and pelvic pain was even higher after influenza (OR 0.95; CI [0.93-0.97]), as was a diagnosis of IBD (OR 0.7; CI [0.68-0.73]; **Supplementary Figure 1**). We confirmed the increased risk of IBD after influenza (1.21% vs 0.65%, respectively; OR 1.87; CI [1.79-1.96]; **Supplementary Table 10-11**) by comparing influenza with matched NCs. Conversely, the risk of a diagnosis of GP was higher after COVID-19 as compared with influenza (OR 1.26; CI [1.12-1.41]). Similarly, GISx such as nausea (OR 1.19; CI [1.16-1.23]), vomiting (OR 1.25; CI [1.20-1.31]), bloating (OR 1.62; CI [1.55-1.70]) and early satiety (OR 1.62; CI [1.43-1.83]) were higher after COVID-19 compared to influenza, as were dysphagia (OR 1.21; CI [1.16-1.26]), and GERD (OR 1.09; CI [1.06-1.12]; **Supplementary Figure 1**).

**Figure 2.**
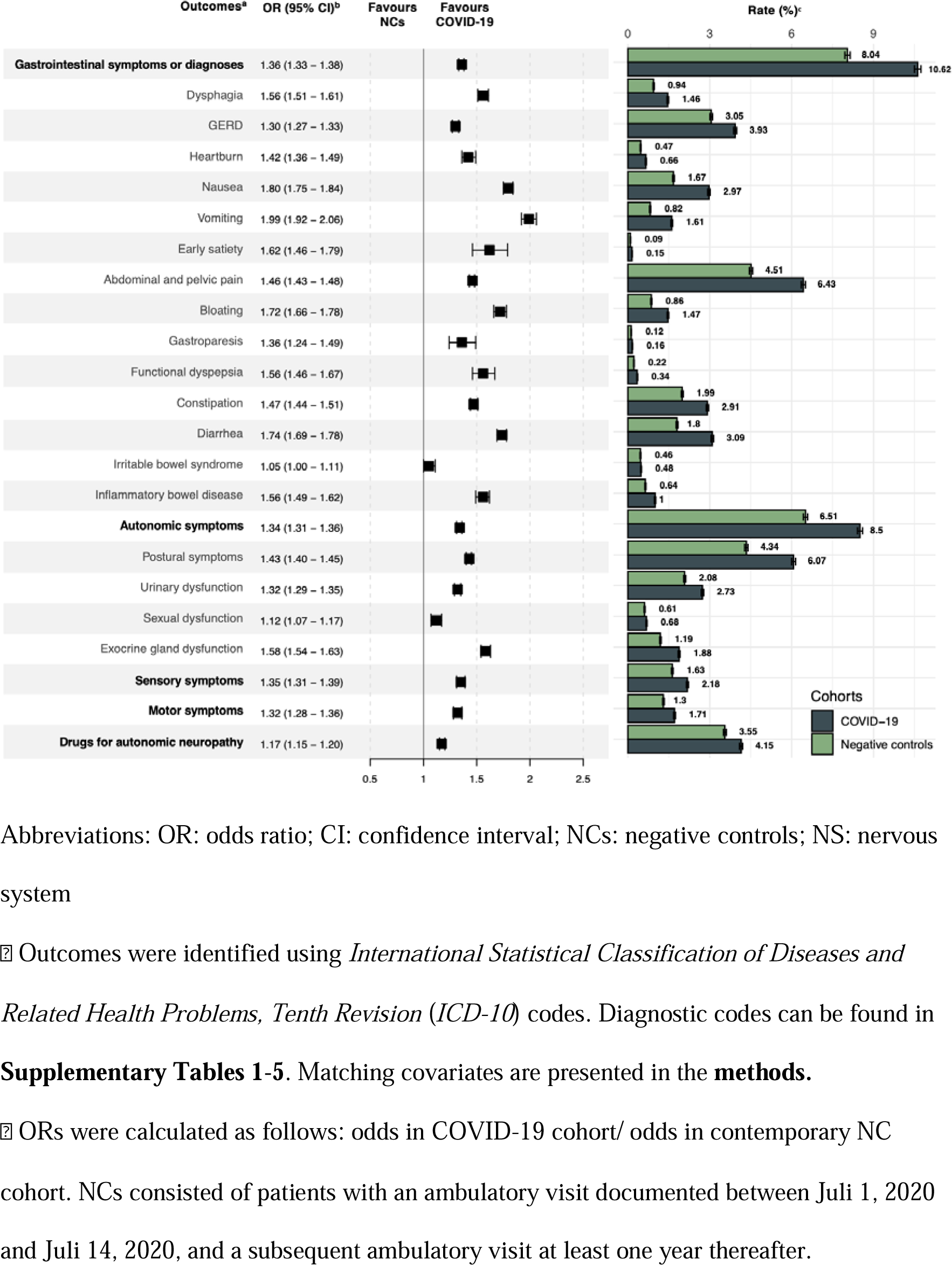
Odds ratios and absolute rates of symptoms and diagnoses between 3 months and one year after COVID-19, compared to matched contemporary NCs.

### Effects of vaccination on neurological and gastrointestinal sequelae

Prior COVID-19 vaccination decreased (P<0.0001) the risk of GISx (OR 0.55; CI [0.47-0.64]), ANSx (OR 0.73; CI [0.64-0.84]) and SNSx (OR 0.64; CI [0.54-0.76]); the decrease in MNSx (OR 0.93; CI [0.79-1.1]) was not significant. For influenza, prior vaccination did not impact the risk of GISx, but it was associated with a higher rate of ANSx (OR 1.12; CI [1.06-1.17]), SNSx (OR 1.12; CI [1.05-1.21]) and MNSx (OR 1.16; CI [1.08-1.24]; **Table 2 and Supplementary Tables 12-14**). Anosmia did not affect any of the primary outcomes (P<0·05; **Supplementary Table 15**). Outcomes did not differ significantly between NCs sampled from different time periods (**Supplementary Tables 16-17)**, and similar trends continued for all major outcomes between 1 and 2 years after the index diagnosis (**Supplementary Table 18**).

**Table 2.** New-onset outcomes between 3 months and 1 year after COVID-19 or influenza, in subjects with prior vaccination compared to without.

|  | After matching for baseline demographics and risk factors |  |  |  |  |
| --- | --- | --- | --- | --- | --- |
| Outcome □ | OR (95% CI) □ | Rate (%) |  | P-value (uncorrected) | P-value (corrected) |
| COVID-19 with prior vaccination vs without prior vaccination (n=18,101) |  |  |  |  |  |
|  |  | Vaccinated | Unvaccinated |  |  |
| Gastrointestinal symptoms or diagnoses | 0.55 (0.47 - 0.64) | 258 / 3,883 (6.73) | 556 / 4,772 (11.65) | <.0001 | <.0001 |
| Autonomic symptoms | 0.73 (0.64 - 0.84) | 378 / 5,400 (7) | 641 / 6,870 (9.33) | <.0001 | <.0001 |
| Sensory symptoms | 0.64 (0.54 - 0.76) | 221 / 12,957 (1.71) | 361 / 13,662 (2.64) | <.0001 | <.0001 |
| Motor symptoms | 0.93 (0.79 - 1.10) | 264 / 13,073 (2.02) | 314 / 14,519 (2.16) | 0.41 | 1 |
| Influenza with prior vaccination vs without prior vaccination (n=87,936) |  |  |  |  |  |
|  |  | Vaccinated | Unvaccinated |  |  |
| Gastrointestinal symptoms or diagnoses | 1.00 (0.95 - 1.05) | 2,703 / 22,703 (11.91) | 3,869 / 32,438 (11.93) | 0.94 | 1 |
| Autonomic symptoms | 1.12 (1.06 - 1.17) | 3,509 / 37,242 (9.42) | 4,095 / 48,024 (8.53) | <.0001 | 0.0004 |
| Sensory symptoms | 1.12 (1.05 - 1.21) | 1,591 / 72,374 (2.20) | 1,521 / 77,535 (1.96) | 0.001 | 0.02 |
| Motor symptoms | 1.16 (1.08 - 1.24) | 1,602 / 75,584 (2.12) | 1,450 / 78,955 (1.84) | 0.0001 | 0.003 |

Abbreviations: OR: odds ratio; CI: confidence interval
□ Outcomes were identified using *International Statistical Classification of Diseases and Related Health Problems, Tenth Revision (ICD-10)* codes. Outcomes and matching covariates can be found in the **methods**.
□ For each OR, the numerator consisted of the cohort with the exposure, the denominator consisted of the cohort without the respective exposure.
□ A p-value was calculated using a Pearson chi-squared test, and correct for FDR using the step-up FDR controlling procedure from Benjamini & Yekutieli using the stats package in R.

## DISCUSSION

Using a nationwide medical record network, we performed a retrospective cohort study to investigate the risk of new-onset GI symptoms and diagnoses, and symptoms referrable to the autonomic, sensory, and motor nervous system following a SARS-CoV-2 infection. By performing direct comparisons with influenza and other pathogens with neurotrophic properties we show that phenomena frequently ascribed to SARS-CoV-2 may manifest with comparable or even higher frequency after other infections, with the notable exception of ANSx.

A summary of the primary outcomes following a SARS-CoV-2 infection compared to NCs and other pathogens are presented in **Table 3**. A SARS-CoV-2 infection was associated with an increased OR of developing GISx, ANSx, SNSx, and MNSx compared to NCs (OR 1.36, 1.34, 1.35, and 1.32; all P<.0001). While these findings suggest that SARS-CoV-2 might be equally pathogenic to all divisions of the PNS in relative terms, most symptoms manifested in the GI system and ANS, affecting 10.6% and 8.5% of individuals, respectively, compared to 2.8% and 1.7% in the SNS and MNS, respectively (**Fig. 2**). These findings provide validation for earlier studies that have characterized PASC.^2^ Having established an association between a SARS-CoV-2 infection with the subsequent development of symptoms in the GI tract and the PNS, we set out to consecutively examine the robustness of these associations relative to other pathogens.

**Table 3.** Summary of differences in the risk of GI and neurological symptoms after a SARS-CoV-2 infection compared to other pathogens.

| Outcome | OR [95% CI] of new-onset symptoms after SARS-CoV-2 versus |  |  |  |  |  |  |
| --- | --- | --- | --- | --- | --- | --- | --- |
|  | Negative controls | Influenza | Human herpesviruses |  |  |  | Lyme's disease |
|  |  |  | Varicella | Herpes Zoster | Infectious Mononucleosis | Cytomegalovirus |  |
| Gastrointestinal symptoms or diagnoses | Increased<br>1.36<br>[1.33-1.38] | No difference | Increased<br>1.17<br>[1.10-1.24] | No difference | No difference | No difference | Increased<br>1.40<br>[1.34-1.46] |
| Autonomic symptoms | Increased<br>1.34<br>[1.31-1.36] | Increased<br>1.13<br>[1.11-1.16] | No difference | Increased<br>1.05<br>[1.03-1.08] | Increased<br>1.11<br>[1.03-1.18] | No difference | Increased<br>1.21<br>[1.16-1.27] |
| Sensory symptoms | Increased<br>1.35<br>[1.31-1.39] | Decreased<br>0.84<br>[0.82-0.87] | Decreased<br>0.49<br>[0.46-0.53] | Decreased<br>0.92<br>[0.89-0.95] | No difference | No difference | No difference |
| Motor symptoms | Increased<br>1.32<br>[1.28-1.36] | Decreased<br>0.94<br>[0.91-0.97] | Decreased<br>0.60<br>[0.55-0.65] | Decreased<br>0.88<br>[0.84-0.91] | No difference | No difference | No difference |

Abbreviations: OR: odds ratio
<sup>a</sup> ORs for statistically significant differences are in parentheses, and were calculated as follows: odds of symptoms after SARS-CoV-2 / odds of symptoms after the comparator pathogen. An OR >1 reflects a higher risk of symptoms after SARS-CoV-2 respective to the comparator cohort, an OR <1 reflects the inverse.
<sup>b</sup> Negative controls refers to subjects without a documented SARS-CoV-2 infection, Influenza refers to patients diagnosed with either an unspecified or specified seasonal variant of Influenza, Lyme's disease refers to those with erythema chronicum migrans due to *Borrelia burgdorferi*, Herpes Zoster refers to shingles/zona, Varicella refers to chickenpox, Infectious Mononucleosis refers to mononucleosis caused by Epstein-Barr Virus, Cytomegalovirus (CMV) refers to an infection with CMV, excluding congenital CMV or mononucleosis. Diagnostic codes and their positive predictive values can be found in the **Supplementary Methods**.

First, we conducted a comparison with Influenza, a virus of significant relevance due to its seasonal recurrence, widespread impact, and clinical similarity to COVID-19 and found a higher incidence of ANSx following COVID-19 (OR 1.13). However, influenza infection demonstrated a higher risk of SNSx and MNSx compared to COVID-19 (OR 0.84 and OR 0.94, respectively). Recently, a cohort study showed that a COVID-19 infection was associated with a lower risk of neurologic diagnoses in the year after infection compared to influenza, among which neuropathy (HR 0.57).^16^ With respect to GISx, we found more variability. We confirmed the increased risk of IBD after influenza (1.21% vs 0.65%, respectively; OR 1.87; CI [1.79-1.96]; **Supplementary Figure 1**) by comparing influenza with matched NCs.

Subsequently, we aimed to compare SARS-CoV-2 to various subtypes of HHV. First, we discuss Varicella (chickenpox) and Herpes Zoster (shingles), as they represent clinical manifestations of the primo-infection and reactivation of the same alpha herpesvirus, the Varicella Zoster Virus (VZV; HHV-3). We observed a higher incidence of new-onset GISx and ANSx, and lower incidence of SNSx and MNSx following COVID-19 compared both VZ and HZ. While a VZV reactivation has been associated with the development of chronic pain (postherpetic neuralgia), a VZV primoinfection has not been as strongly linked to long-term complications.^17^ While some authors report that patients with VZV primo-infection can suffer from long-term cognitive impairment,^18^ our data suggests that both a VZV primoinfection and reactivation may be followed by long-term sensory and motor symptoms. Subsequently, we compared SARS-CoV-2 to IM, the clinical syndrome caused by the gamma herpesvirus EBV (HHV-4). While both infections yielded comparable rates of peripheral neurological symptoms, an increase in new-onset GISx following COVID-19 was observed compared to IM (OR 1.11). While EBV has well-established associations with a multitude of neurological conditions, among which multiple sclerosis and small-fiber neuropathy, the underlying mechanisms remain to be established.^19^ Consequently, we sought to investigate the beta herpesvirus CMV (HHV-5), which has been recognized as a potential causative factor in acute and occasionally severe neurological complications among immunocompromised individuals, and paucisymptomatic infections in immunocompetent individuals.^20^ Our study did not reveal any significant differences between neurological and GI sequelae following COVID-19 and CMV, keeping in mind the smaller sample sizes for this comparison and therefore lower grade of confidence. Our data suggests that COVID-19, EBV and CMV might be equally pathogenic for various parts of the peripheral nervous system.^20^

Finally, COVID-19 was compared with the spirochaete Borrelia Burgdorferi, which usually presents as erythema chronicum migrans, referred to as LD. While LD has been linked to the development of long-lasting neurological symptoms, referred to as neuroborreliosis, it has been surrounded by controversy ever since. Our study showed comparable rates of SNSx and MNSx after both infections. However, COVID-19 yielded a significantly higher risk of ANSx and GISx (OR 1.21 and 1.40, respectively). Neuroborreliosis typically manifests during the early disseminated phase of the disease, characterized by cranial and peripheral nerve involvement, and is usually highly responsive to oral doxycycline.^21^ However, as far as symptoms outlasting the acute illness are concerned, controlled studies have failed to establish an association with long-term neurological complications.^22^ If anything, the increase in ANSx and GISx after a SARS-CoV-2 infection compared to LD reiterates the affinity of SARS-CoV-2 for these symptom clusters.

In summary, all primary outcomes were increased following COVID-19 compared to NCs – confirmatory of the literature to date – but only ANSx appeared to be consistently increased relative to other pathogens (other than CMV and IM).^23^ Specifically, more than 8% of patients developed at least 1 new ANSx, predominantly comprising postural symptoms. Interestingly, our study validated earlier reports of an increase in SNSx and MNSx after COVID-19 compared to non-infected controls but failed to replicate this association compared to other pathogens. Moreover, VZV primoinfection and reactivation consistently yielded a greater risk of developing SNSx and MNSx, as did influenza. In addition, we observed new-onset GISx in approximately 10·6% of COVID-19 patients, which was not significantly different from other prevalent infectious diseases such as influenza and HHV. However, the risk for specific symptoms and diagnoses like GP and related symptoms was higher after COVID-19 than after influenza. Even though these diagnoses were among the least common in this study, and no difference was found for the related syndrome FD, the symptoms comprising these two related syndromes were much more common. Considering the current healthcare burden of these functional GI disorders in both primary and specialist care, even a modest relative increase would further strain the system and necessitate attention. Of note, our study found the risk of IBD to be increased after COVID-19 compared to NCs (OR: 1·56), and an even stronger association was found for influenza (OR: 1.87). While environmental triggers are generally accepted to play a pivotal role in the pathogenesis of IBD, a causal link with infections such as influenza remains elusive.^24^ Our data supports the concept that infections that cause gut dysbiosis might perpetuate high-grade inflammation in susceptible individuals.^24^ Intriguingly, preclinical studies have shown that respiratory influenza infections can induce intestinal injury through T helper 17 cell-dependent mechanisms and by disrupting the integrity of the gut barrier.^25,26^ Large-scale prospective studies are highly necessary to establish the absolute risks for each of these sequelae.

Furthermore, our study found prior vaccination for COVID-19 to be protective against the development of GISx, ANSx and SNSx. Using real-world data, this study provides reassuring evidence in support of earlier studies, which demonstrated that COVID-19 vaccines protect against long-term neurological and GI sequelae following SARS-CoV-2. ^27–29^ The exact mechanism remain unclear, but might involve a reduction in the severity of the infection and the associated inflammatory or immune responses.^28,29^ Notably, an influenza vaccination in our study appeared to predispose to the development of more ANS, SNS and MNS disturbances. Although neurological complications (e.g. transverse myelitis) have been anecdotally described following an influenza vaccine, other complications remain rare.^30^ It is possible that the increase in complications after a vaccination for influenza in our study is partially explained by a form of assignment bias. Despite rigorous matching, outcomes might be confounded by persistent differences in cohort constitution in this real-world dataset, resulting from vaccination program characteristics. During the pandemic, COVID-19 vaccines have been gradually made available to almost the entire population, which is generally not applicable to a seasonal influenza vaccination program in which the indication to receive a vaccine is often made based on premorbid conditions. This way, subjects with comorbidities that predispose to severe outcomes are more likely to have been assigned to the vaccinated cohort, leading to assignment bias. Primary outcomes did not differ in subjects with versus without anosmia in our study.

Owing to its utility, the use of EHR data is constrained by inherent limitations, including the necessity to resort to ICD-10 codes to establish diagnoses. It is also imperative to note that the exact anatomical origin of the neurological symptoms remains speculative. While most infectious diseases in this study require serological confirmation, we were not able to provide confirmation for these physician-coded diagnoses. The positive predictive values for each ICD-10 code are summarized in the **Supplementary Methods**. Despite exhaustive matching for comorbid conditions, underlying differences in cohort constitution may persist, influencing outcomes in ways unrelated to the index event, resulting in an assignment bias. Additionally, co-occuring infectious diseases were neither assessed nor excluded in the present study. Nevertheless, its strength lies in its large sample size, and inclusion of a wide range of infectious diseases. The multi-center character and inclusion of racially and ethnically diverse subjects ensure that these results are generalizable to patients at academic medical centers across the US.

In summary, this study is the first to use a nationwide network to compare long-term neurological and GI sequelae after COVID-19 to those after other infectious diseases. Our study shows that phenomena frequently ascribed to COVID-19 may manifest with comparable or even higher frequency after other infections, with the notable exception of ANSx. While these findings suggest a need to reassess the relative impact of COVID-19, it is equally crucial to recognize the significant and often overlooked role that common infectious diseases play in causing neurological or GI symptoms. However, despite any similarities in relative impact, the unparalleled number of COVID-19 infections makes postinfectious complications from COVID-19 a greater public health risk than those from other infections. In this regard, our study found prior vaccination for COVID-19 to be protective against the development of GI symptoms, and ANS and SNS disturbances, which, mindful of the spread of COVID-19, supports the notion that it had significant beneficial effects in preventing chronic sequelae.

## METHODS

### Study design

To investigate the association between a COVID-19 infection and the subsequent development of new onset GI symptoms and diagnoses (GISx), and symptoms referrable to the autonomic (ANSx), sensory (SNSx) and motor nervous systems (MNSx), we performed a retrospective cohort-study using Electronic Medical Health Record (EHR) data from the TriNetX Research Network (Cambridge, Massachusetts, USA). The network comprised over 84 million patients from 57 predominantly academic medical centers in the US at the time of data-collection. TriNetX provides a web-based platform to create and compare cohorts based on prespecified in- and exclusion criteria for the development of specified outcomes. The platform includes a built-in propensity score-matching (PSM) algorithm to match cohorts for potential confounders. More information about the network can be found in the online supplement (**Supplementary Methods**). Each participating organization confirmed obtaining consent to share aggregated de-identified participant data for research purposes. The Institute of Clinical and Translational Research (ICTR) at Johns Hopkins University managed access to end-users. This study was exempt from Institutional Review Board (IRB) review.

### Participants

First, we compared adult (≥ 18 years old) patients with a first diagnosis of COVID-19 with matched contemporary negative controls (NCs). COVID-19 cases were captured using the International Statistical Classification of Diseases and Related Health Problems, Tenth Revision (ICD-10) diagnosis of COVID-19 (U07.1) and 18 Current Procedural Terminology (CPT) codes capturing positive diagnostic tests, when documented between Jan 1, and Dec 31, 2020. Contemporary NCs comprised of patients with an ambulatory visit between July 1, 2020 and July 14, 2020, without a diagnosis of COVID-19. To determine COVID-19 specific sequelae, we not only compared COVID-19 cases with contemporary negative controls (NCs), but also with subjects who received a ICD-10 diagnosis of Influenza, Lyme’s disease (LD), or infections by several human herpesviruses (HHV), including Cytomegalovirus (CMV), Herpes Zoster (HZ; “shingles”), Varicella (VZ; “chickenpox”), and Infectious Mononucleosis (IM) between Jan 1, 2006 and Dec 31, 2020; more information about the ICD-10 codes and their positive predictive values (PPV) can be found in the **supplementary methods**. A minimum of one year of prospective follow-up was ensured for each cohort by requiring at least one documented ambulatory visit at least one year after the respective index diagnosis. This index diagnosis (i.e., first adult diagnosis of the respective disease or first of two ambulatory visits in NCs) was then used as the index event.

### Outcomes

We compared the COVID-19 cohort with matched control cohorts for the ‘prospective’ risk of developing new-onset GISx, ANSx, SNSx, and MNSx occurring between three months and one year after the respective diagnosis. In a pairwise fashion, COVID-19 patients were propensity score matched to each control cohort for age at diagnosis, sex (as reported in medical records), race and ethnicity, and risk factors associated with the development of COVID-19 and more severe forms of COVID-19.^13^ To investigate the temporal continuity of the observed trends, outcomes between 1 and 2 years post-index diagnosis were collected. In addition, patients with either anosmia during the acute phase or a prior COVID-19 vaccination were compared to those without. Similarly, we assessed the effects of vaccination on the development of new-onset symptoms after influenza. To examine the possibility of selection bias, historic NCs and contemporary NCs from multiple time periods were sampled and compared.

### Variables of interest

Matching covariates included fibrosis and cirrhosis of the liver, cerebral infarction, dementia, rheumatoid arthritis, lupus erythematosus, psoriasis, overweight and obesity, hypertension, diabetes mellitus, chronic kidney disease, asthma, chronic lower respiratory diseases, smoking, ischemic heart disease, heart disease, heart failure and neoplasms.^13^ Four primary composite outcomes consisted of GI symptoms and diagnoses (GISx), and symptoms referrable to the autonomic (ANSx), sensory (SNSx) and motor nervous systems (MNSx). GI symptoms included dysphagia, heartburn, nausea, vomiting, early satiety, abdominal and pelvic pain, bloating, constipation, and diarrhea, and ICD-10 based diagnoses included gastro-esophageal reflux disease, gastroparesis, functional dyspepsia, irritable bowel syndrome, and inflammatory bowel disease. ANS disturbances were categorized into 4 groups: postural symptoms (dizziness, syncope, visual disturbances, vertigo, gait abnormality, coordination difficulties, repeated falls, chronic fatigue, palpitations), urinary dysfunction (frequency, nocturia, urgency, stress incontinence, urinary retention, hesitancy, unspecified neuromuscular dysfunction of the bladder), sexual dysfunction (erectile dysfunction, atrophy of the vulva, decreased libido, unspecified sexual dysfunction) and sweat- and exocrine gland dysfunction (excessive or decreased sweating, dryness of mouth, dry eye syndrome). SNS disturbances included paresthesia and hyperesthesia of the skin and unspecified skin disturbances; MNS disturbances included generalized muscle weakness, muscle spasms and contractures, muscle wasting and atrophy, and paralytic syndrome. Additionally, to increase confidence in the veracity of our results, we also collected prescriptions commonly used for ANS disturbances as an independent validator. These prescriptions included fludrocortisone, betablockers, pyridostigmine and midodrine.

### Statistical analysis

Categorical and continuous variables are presented as number (percentage) and mean (standard deviation), respectively. Standardized Mean Differences (SMDs) were used to assess differences at baseline, an SMD of <0.2 was considered well balanced.^14^ A log-rank test was performed to compare outcomes, and a two-sided p-value of <0.05 was set as statistical significance. The step-up FDR controlling procedure from Benjamini & Yekutieli ^15^ was used to correct p-values for multiple comparisons, using the ‘stats’ package in R (v3.6.2; R Core Team; 2022). To capture new-onset outcomes, we excluded subjects with the respective outcome prior to the index event after performing propensity score matching (PSM).

## Acknowledgements

BK was supported by a Fellowship of the Belgian American Educational Foundation. We gratefully acknowledge Dr. Maxime Taquet, MD, PhD (University of Oxford, UK) for his statistical support.

## CRediT Author statement

**BK:** Conceptualization, Methodology, Formal analysis, Data curation, Validation, Visualization, Writing - Original draft, Writing – Review and Editing, **LV**: Conceptualization, Resources, Investigation, **RB:** Conceptualization, Resources, Investigation, **GB:** Conceptualization, Resources, Investigation, **MM:** Conceptualization, **KH:** Conceptualization, Methodology, Validation, **JT:** Writing – Review and Editing, **PJP:** Supervision, Conceptualization, Methodology, Validation, Writing – Review and Editing

## Data availability

Data were exported from TriNetX in .csv files and archived. Every (co-) author affiliated to Johns Hopkins University was granted access to the TriNetX Research network by the Institute of Clinical and Translational Research (ICTR). The supplementary materials contain an extensive list of tables representing the original data; researchers will be granted access to the original data for specific concerns upon approval from the corresponding author (PJP).

