## Supplementary material for "Long-term neurological and gastrointestinal sequelae of SARS-CoV-2, influenza, and other neurotropic infections with and without vaccination": Online Supplement

### **Supplementary Methods**

#### **Data source and data collection:**

The TriNetX Research network provides real-time access to aggregated longitudinal data, anonymized at a patient and organizational level. The participating centers of this Federated health research network primarily consisted of academic medical centers located in the USA. Captured information includes demographics, diagnoses, procedures, medications, and visits from patients independent of insurance status. Structured data in the medical records is mapped to standard and controlled clinical terms in the database (12) and is formatted in the most updated version of the of International Classification of Disease (ICD-10), Current Procedural Terminology (CPT 2021 version), Health Common Procedure Coding System (HCPCS 2021 version), Systematized Nomenclature of Medicine (SNOMED-CT 2021 version), the RxNorm coding language and native TriNetX (TNX) codes, allowing for reliable comparison of data between participating medical centers. ICD-9 codes are mapped to corresponding ICD-10 codes. These codes can be used as in- and exclusion criteria to define and query patient cohorts, after which they can be matched for confounding factors using a built-in propensity score matching (PSM) algorithm (see below). An index event is chosen for each cohort to determine starting point for the collection of outcomes over a predefined retrospective or prospective follow-up. Using the built-in web-based platform (live.trinetx.com), outcome data is generated based on pre-specified criteria. These criteria include the selection of comparator cohorts, matching covariates, follow-up duration, and the inclusion or exclusion of subjects with a documented outcome of interest prior to the investigated time window (distinguishing new-onset from non-new-onset outcomes). Subsequently, outcome measures are selected to generate the outcome data. Possible outcome measures include measures of association (MAO; odds ratios, absolute risk, relative risk), Kaplan-Meier data (hazard ratios, cumulative incidence), among others. Demographic and outcome data were collected as separate .csv files and summarized in an .xlsx file for further formatting and data visualization. The Institute of Clinical and Translational Research (ICTR) manages the TriNetX platform at our institution and provides access to the end-users. The use of aggregated, de-identified data from TriNetX did not require institutional board (IRB) review. The study was conducted according to the **STROBE** guidelines.

#### **Propensity score matching:**

Cohorts were matched using a built-in propensity score matching tool in TriNetX. Using logistic regression, a propensity score for each covariate in each patient was calculated using the ‘scikit-learn’ package in Python (version 3.7). Greedy nearest neighbor matching with a caliper of 0.1 pooled standard deviations of the aggregated propensity scores was used to create 1:1 matched subsets of patients. A randomization of the order of records was performed before performing nearest neighbor matching to reduce bias caused by order of rows in each covariate matrix after to centralization of pooled covariate matrices from each participating HCO.

#### **Positive predictive values for ICD-10 codes:**

The positive predictive value (PPV) of International Statistical Classification of Diseases and Related Health Problems, Tenth Revision (ICD-10) codes is the measure that quantifies the proportion of patients of those identified by the diagnostic code, that have the illness. Although the ICD-10 coding system is widely used in epidemiological research, it inherently suffers from certain limitations, most prominently being the lack of information on confirmatory diagnostic tests which have led up to a specific diagnosis. To validate the use of these diagnostic codes in epidemiological research, various validation studies have been set up for specific frequently used diagnostic codes. The paragraph below is a (non-systematic) summary of PPVs found in the literature for the diagnostic codes used for comparator cohorts.

**Influenza virus infection**: The codes used were J10.1 and J11.1. While J10.1 refers to an in-office diagnosis of a *specified* seasonal influenza variant with upper respiratory tract manifestations, J11.1 refers to an in-office diagnosis of an *unspecified* influenza variant with upper respiratory tract manifestations. Following the ICD-10 coding book, J11.1 is often referred to influenza not otherwise specified (NOS). In a study conducted by Hamilton et al, the PPV for the use of both the specified and unspecified influenza variants combined was 0.91 (95% CI: 0.91-0.92).[1]

**Cytomegalovirus infection**: The code used was B25.9. Of note, the code B25 (and its subcategories) exclude congenital CMV infection and CMV mononucleosis. This ICD-10 diagnosis is a clinician-based diagnosis for active CMV infection, but it is uncertain whether confirmative serology, PCR, or histopathology results were obtained. A study by # et al found that the PPV of the ICD-9 based diagnosis for CMV was 0.71 in predicting microbiologic, histologic, or ophthalmologic evidence of active CMV infection or disease.[2] No studies were found validating the ICD-10 diagnosis B25.9.

**Herpes Zoster (shingles)**: The code used was B02. The code B02 refers primarily to dermatological manifestations such as shingles and zona. In an earlier study, the ICD-9 code for HZ (053) had a PPV of 0.93 for detecting HZ infections.[3] A subsequent validation study in Korea found the PPV of the ICD-10 diagnosis B02 to detect definite cases to be 0.76, and definite and possible cases 0.86, respectively.[4]

**Varicella (chickenpox)**: The code used was B01. The code B01 refers to the manifestation of VZV referred to as chickenpox. The PPV of the ICD-10 diagnosis B01 for varicella/chickenpox was found to be about 60.5% in a CDC Report.[5] In the conclusion of the report, certain limitations regarding the use of this diagnostic code were mentioned: “This definition has some limitations, including some false positives that remain (though infrequently), including hives, insect bites, allergic reactions, and unspecified dermatitis. Additionally, varicella identified using this definition may reflect prior history of disease rather than acute disease. Some hospitals enter all diagnosis codes from a patient’s entire medical history, including prior history of varicella.”[5]

**Infectious mononucleosis**: The code used was B27.9. The most common causative agent for infectious mononucleosis (with a clinical triad of fever, lymphadenopathy and pharyngitis) is the Epstein-Barr Virus. However, using this code, serological confirmation of EBV infection is not available. To our knowledge, no studies were conducted investigating the PPV of B27.9 in predicting IM caused by EBV.

**Lyme’s disease**: The code used was A69.20, and more specifically corresponds to erythema chronicum migrans caused by Borrelia Burgdorferi’. A validation study using US based claims data found that this ICD-10 code has a PPV of 0.66 for correctly identifying a probable or confirmed diagnosis of Lyme’s disease. This increases to 0.93 when suspected cases are also included.[6]

#### **Diagnostic codes**

| Coding language | Code | Variable of interest |
| --- | --- | --- |
| COVID-19 | | |
| ICD-10 | U07.1 | COVID-19 |
| CPT | 87635 | Infectious agent detection by nucleic acid (DNA or RNA); severe acute respiratory syndrome coronavirus 2 (SARS-CoV-2) (Coronavirus disease [COVID-19]), amplified probe technique |
|  | U0001 | 2019 Novel Coronavirus Real Time RT-PCR Diagnostic Test Panel - CDC |
|  | U0002 | 2019 Novel Coronavirus Real Time RT-PCR Diagnostic Test Panel - non-CDC |
|  | 9088 | SARS coronavirus 2 and related RNA [Presence] |
|  | 94307-6 | SARS-CoV-2 (COVID-19) N gene [Presence] in Unspecified specimen by Nucleic acid amplification using CDC primer-probe set N1 |
|  | 94310-0 | SARS-like coronavirus N gene [Presence] in Unspecified specimen by NAA with probe detection |
|  | 94309-2 | SARS-CoV-2 (COVID-19) RNA [Presence] in Unspecified specimen by NAA with probe detection |
|  | 94314-2 | SARS-CoV-2 (COVID-19) RdRp gene [Presence] in Unspecified specimen by NAA with probe detection |
|  | 94315-9 | SARS-related coronavirus E gene [Presence] in Unspecified specimen by NAA with probe detection |
|  | 94316-7 | SARS-CoV-2 (COVID-19) N gene [Presence] in Unspecified specimen by NAA with probe detection |
|  | 94500-6 | SARS-CoV-2 (COVID-19) RNA [Presence] in Respiratory specimen by NAA with probe detection |
|  | 94502-2 | SARS-related coronavirus RNA [Presence] in Respiratory specimen by NAA with probe detection |
|  | 94533-7 | SARS-CoV-2 (COVID-19) N gene [Presence] in Respiratory specimen by NAA with probe detection |
|  | 94534-5 | SARS-CoV-2 (COVID-19) RdRp gene [Presence] in Respiratory specimen by NAA with probe detection |
|  | 94559-2 | SARS-CoV-2 (COVID-19) ORF1ab region [Presence] in Respiratory specimen by NAA with probe detection |
|  | 94565-9 | SARS-CoV-2 (COVID-19) RNA [Presence] in Nasopharynx by NAA with non-probe detection |
|  | 94758-0 | SARS-related coronavirus E gene [Presence] in Respiratory specimen by NAA with probe detection |
|  | 94759-8 | SARS-CoV-2 (COVID-19) RNA [Presence] in Nasopharynx by NAA with probe detection |
| Other diagnoses | | |
| ICD-10 | J10.1 | Influenza |
|  | J11.1 | Influenza |
|  | B02 | Herpes Zoster |
|  | B01 | Varicella zoster |
|  | B27.9 | Infectious mononucleosis |
|  | B25.9 | Cytomegalovirus |
|  | A69.20 | Lyme's disease |
| Outcomes | | |
| GI symptoms and diagnoses | | |
| ICD-10 | R11.0 | Nausea |
|  | R11.10 | Vomiting |
|  | R10.00 | Abdominal and pelvic pain |
|  | R68.81 | Early satiety or postprandial fulness |
|  | R14.0 | Bloating |
|  | R19.06 | Bloating |
|  | R12 | Heartburn |
|  | K59 | Constipation |
|  | R19.7 | Diarrhea |
|  | K30 | Functional dyspepsia |
|  | K31.84 | Gastroparesis |
|  | R13.1 | Dysphagia |
|  | K58.00 | Irritable bowel syndrome |
|  | K21.00 | Gastro-esophageal reflux disease |
|  | K50 + K51 | Inflammatory bowel disease |
| Autonomic nervous system disturbances | | |
| ICD-10 | G90 | Autonomic neuropathy |
| Postural symptoms | | |
| ICD-10 | R42 | Lightheadedness & dizziness |
|  | R55 | Fainting |
|  | H53.8 | Visual disturbances |
|  | H81.4 | Vertigo |
|  | R26.9 | Unsteady gait |
|  | R29.6 | Unsteady gait |
|  | R27.8 | Unsteady gait |
|  | R53.82 | Weakness |
|  | R00.2 | Palpitations |
| Urinary dysfunction | | |
| ICD-10 | R35.0 | Frequency |
|  | R35.1 | Excessive urinating at night |
|  | R39.15 | Urgency |
|  | R39.3 | Stress incontinence |
|  | R33.9 | Retention |
|  | R39.11 | Hesitancy |
|  | N31.9 | Unspecified neuromuscular dysfunction of bladder |
| Sexual dysfunction | | |
| ICD-10 | R37 | Sexual dysfunction, unspecified |
|  | N52.9 | Erectile dysfunction |
|  | N90.5 | Vaginal dryness |
|  | R68.82 | Decreased libido |
| Exocrine gland dysfunction | | |
| ICD-10 | R61 | Excessive or decreased sweating |
|  | L74.4 | Excessive or decreased sweating |
|  | R68.2 | Dryness of mouth |
|  | H04.12 | Dry eyes syndrome |
| Sensory nervous system disturbances | | |
| ICD-10 | R20.2 | Paresthesias: numbness or tingling in extremities |
|  | R20.3 | Hyperesthesia |
|  | R20.9 | Unspecified disturbances of skin sensation |
| Motor nervous system disturbances | | |
| ICD-10 | M62.81 | Muscle weakness |
|  | M62.83 | Painful cramps or fasciculations |
|  | M62.40 | Painful cramps or fasciculations |
|  | M62.50 | Muscle shrinking |
|  | G83.9 | Paralysis or loss of muscle control |
| Drugs for autonomic neuropathy: | | |
| TNX |  | Fludrocortisone |
|  |  | Beta-blockers |
|  |  | Pyridostigmine |
|  |  | Midodrine |
| Matchig covariates | | |
| TNX | AI | Age at Index |
|  | 2186-5 | Not Hispanic or Latino |
|  | 2106-3 | White |
|  | F | Female |
|  | M | Male |
|  | 2054-5 | Black or African American |
|  | UN | Unknown Ethnicity |
|  | 2135-2 | Hispanic or Latino |
| ICD-10 | I10 | Essential (primary) hypertension |
|  | C00-D49 | Neoplasms |
|  | E66 | Overweight and obesity |
|  | E08-E13 | Diabetes mellitus |
|  | I20-I25 | Ischemic heart diseases |
|  | J45 | Asthma |
|  | N18 | Chronic kidney disease (CKD) |
|  | F17 | Nicotine dependence |
|  | J44.9 | Chronic obstructive pulmory disease, unspecified |
|  | I63 | Cerebral infarction |
|  | M06.9 | Rheumatoid arthritis, unspecified |
|  | L40 | Psoriasis |
|  | K74 | Fibrosis and cirrhosis of liver |
|  | F03 | Unspecified dementia |
| Other baseline (Charleson) comorbidities, not included in matching covariates | | |
| ICD-10 | J40-J47 | Chronic lower respiratory diseases |
|  | I50 | Heart failure |
|  | N17 | Acute kidney failure |
|  | I73.9 | Peripheral vascular disease, unspecified |
|  | I21 | Acute myocardial infarction |
|  | I51.9 | Heart disease, unspecified |
|  | N18.6 | End stage rel disease |
|  | K27 | Peptic ulcer, site unspecified |
|  | K76.6 | Portal hypertension |
|  | B20 | Human immunodeficiency virus [HIV] disease |
| Influenza vaccination | | |
| CPT | 90630 | Influenza virus vaccine, quadrivalent (IIV4), split virus, preservative free, for intradermal use 90653 Influenza vaccine, inactivated (IIV), subunit, adjuvanted, for intramuscular use |
|  | 90654 | Influenza virus vaccine, trivalent (IIV3), split virus, preservative-free, for intradermal use OR |
|  | 90655 | Influenza virus vaccine, trivalent (IIV3), split virus, preservative free, 0.25 mL dosage, for intramuscular use |
|  | 90656 | Influenza virus vaccine, trivalent (IIV3), split virus, preservative free, 0.5 mL dosage, for intramuscular use |
|  | 90657 | Influenza virus vaccine, trivalent (IIV3), split virus, 0.25 mL dosage, for intramuscular use |
|  | 90658 | Influenza virus vaccine, trivalent (IIV3), split virus, 0.5 mL dosage, for intramuscular use |
|  | 90660 | Influenza virus vaccine, trivalent, live (LAIV3), for intranasal use |
|  | 90661 | Influenza virus vaccine, trivalent (ccIIV3), derived from cell cultures, subunit, preservative and antibiotic free, 0.5 mL dosage, for intramuscular use |
|  | 90662 | Influenza virus vaccine (IIV), split virus, preservative free, enhanced immunogenicity via increased antigen content, for intramuscular use |
|  | 90672 | Influenza virus vaccine, quadrivalent, live (LAIV4), for intranasal use |
|  | 90673 | Influenza virus vaccine, trivalent (RIV3), derived from recombinant DNA, hemagglutinin (HA) protein only, preservative and antibiotic free, for intramuscular use |
|  | 90674 | Influenza virus vaccine, quadrivalent (ccIIV4), derived from cell cultures, subunit, preservative and antibiotic free, 0.5 mL dosage, for intramuscular use |
|  | 90682 | Influenza virus vaccine, quadrivalent (RIV4), derived from recombinant DNA, hemagglutinin (HA) protein only, preservative and antibiotic free, for intramuscular use |
|  | 90685 | Influenza virus vaccine, quadrivalent (IIV4), split virus, preservative free, 0.25 mL dosage, for intramuscular use |
|  | 90686 | Influenza virus vaccine, quadrivalent (IIV4), split virus, preservative free, 0.5 mL dosage, for intramuscular use |
|  | 90687 | Influenza virus vaccine, quadrivalent (IIV4), split virus, 0.25 mL dosage, for intramuscular use 90688 Influenza virus vaccine, quadrivalent (IIV4), split virus, 0.5 mL dosage, for intramuscular use 90689 Influenza virus vaccine, quadrivalent (IIV4), inactivated, adjuvanted, preservative free, 0.25 mL dosage, for intramuscular use |
|  | 90694 | Influenza virus vaccine, quadrivalent (aIIV4), inactivated, adjuvanted, preservative free, 0.5 mL dosage, for intramuscular use |
|  | 90756 | Influenza virus vaccine, quadrivalent (ccIIV4), derived from cell cultures, subunit, antibiotic free, 0.5 mL dosage, for intramuscular use |
| HCPCS | Q2034 | Influenza virus vaccine, split virus, for intramuscular use (agriflu) |
|  | Q2035 | Influenza virus vaccine, split virus, when administered to individuals 3 years of age and older, for intramuscular use (afluria) |
|  | Q2036 | Influenza virus vaccine, split virus, when administered to individuals 3 years of age and older, for intramuscular use (flulaval) |
|  | Q2037 | Influenza virus vaccine, split virus, when administered to individuals 3 years of age and older, for intramuscular use (fluvirin) |
|  | Q2038 | Influenza virus vaccine, split virus, when administered to individuals 3 years of age and older, for intramuscular use (fluzone) |
|  | Q2039 | Influenza virus vaccine, not otherwise specified |
|  | G0008 | Administration of influenza virus vaccine |

### **Supplementary Figures**

#### **Supplementary Figure 1**. Odds ratios and absolute rates of new-onset outcomes between 3 months and one year after COVID-19, compared to influenza.

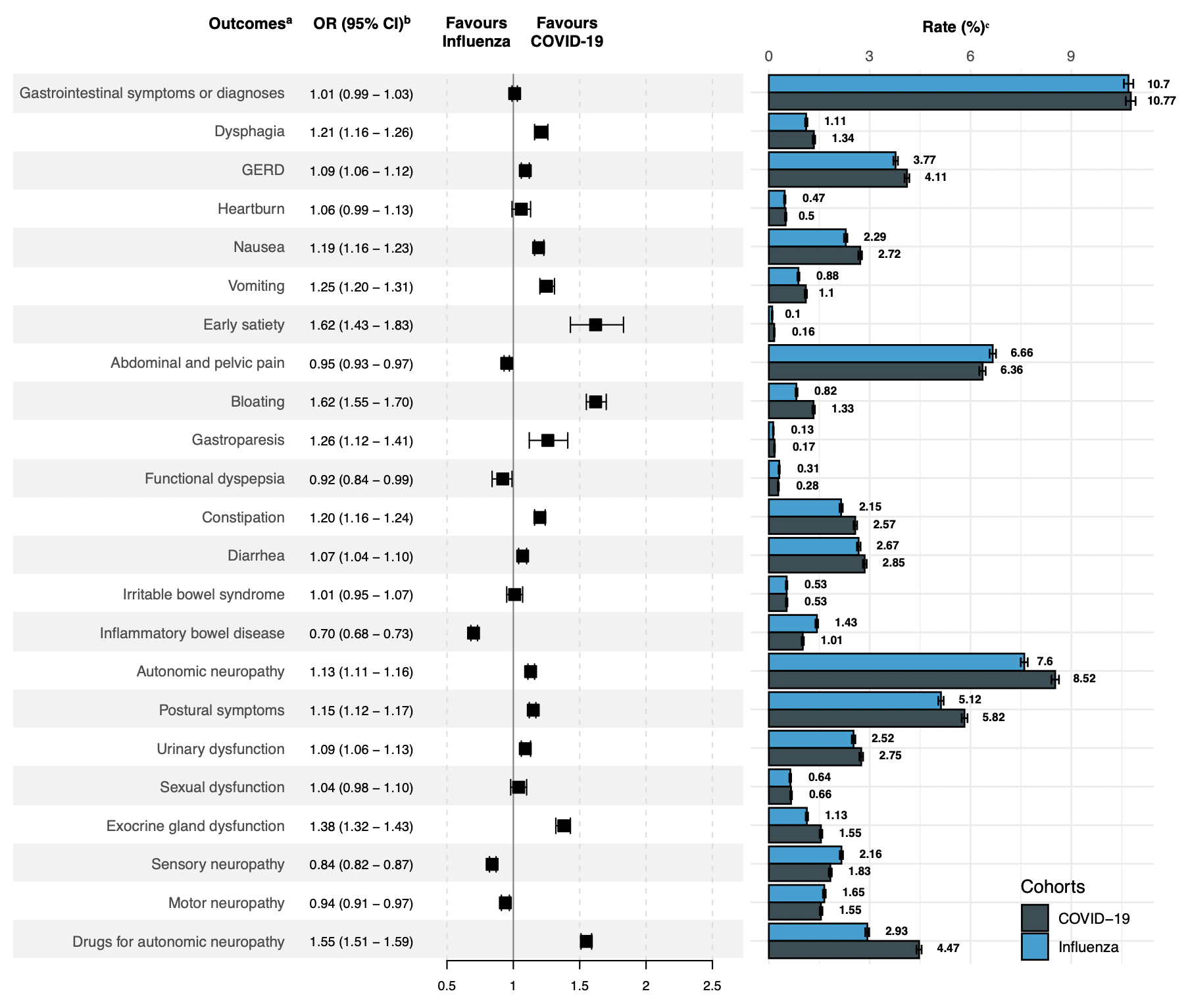

Abbreviations: GERD: gastro-esophageal reflux disease; OR: odds ratio; CI: confidence interval; NCs: negative controls

ᵃ Outcomes were identified using *International Statistical Classification of Diseases and Related Health Problems, Tenth Revision* (*ICD-10*) codes **(supplementary Methods).**

ᵇ ORs were calculated as follows: odds in COVID-19 cohort/ odds in contemporary NC cohort. NCs consisted of patients with an ambulatory visit documented between Juli 1, 2020 and Juli 14 2020, and a subsequent ambulatory visit at least one year thereafter.

ᶜ Cumulative incidence of new-onset outcomes during the follow-up.

### **Supplementary Tables**

#### **Supplementary Table 1.** Baseline demographics of COVID-19 compared to contemporary NCs and Influenza, before and after matching.

|  | **Before matching for baseline demographics and risk factors** | | | | | | **After matching for baseline demographics and risk factors ᵇ** | | | | | |
| --- | --- | --- | --- | --- | --- | --- | --- | --- | --- | --- | --- | --- |
| **Characteristics ᵃ** | COVID-19 | Contemporary negative controls |  | COVID-19 | Influenza |  | COVID-19 | Contemporary negative controls |  | COVID-19 | Influenza |  |
|  | No. (%) |  | SMD | No. (%) |  | SMD | No. (%) |  | SMD | No. (%) |  | SMD |
| **Number** | 649478 | 2890799 | - | 469233 | 462530 | - | 649311 | 649311 | - | 410458 | 410458 | - |
| **Age; mean (SD); y** | 49.2 (17.6) | 55.7 (17.5) | 0.4 | 49.4 (17.7) | 46.6 (17.3) | 0.2 | 49.2 (17.6) | 49.1 (17.5) | 0.005 | 47.8 (17.4) | 47.8 (17.3) | 0.0002 |
| **Sex** |  |  |  |  |  |  |  |  |  |  |  |  |
| Female | 399408 (61.5) | 1775777 (61.4) | 0.001 | 292696 (62.4) | 289062 (62.5) | 0.002 | 399302 (61.5) | 395457 (60.9) | 0.01 | 257944 (62.8) | 257151 (62.6) | 0.0003 |
| Male | 249976 (38.5) | 1114714 (38.6) | 0.001 | 176467 (37.6) | 173393 (37.5) | 0.002 | 249915 (38.5) | 253764 (39.1) | 0.01 | 152449 (37.1) | 153234 (37.3) | 0.0002 |
| **Race** |  |  |  |  |  |  |  |  |  |  |  |  |
| White | 450333 (69.3) | 2097083 (72.5) | 0.07 | 313989 (66.9) | 299565 (64.8) | 0.05 | 450253 (69.3) | 446495 (68.8) | 0.01 | 272669 (66.4) | 272977 (66.5) | 0.02 |
| Black or African American | 117279 (18.1) | 386394 (13.4) | 0.1 | 80276 (17.1) | 71883 (15.5) | 0.04 | 117247 (18.1) | 121390 (18.7) | 0.02 | 66354 (16.2) | 66585 (16.2) | 0.004 |
| **Ethnicity** |  |  |  |  |  |  |  |  |  |  |  |  |
| Hispanic or Latino | 59593 (9.2) | 119134 (4.1) | 0.2 | 49080 (10.5) | 42596 (9.2) | 0.04 | 59462 (9.2) | 61991 ( 9.5) | 0.01 | 40555 ( 9.9) | 40319 ( 9.8) | 0.01 |
| Not Hispanic of Latino | 458959 (70.7) | 2176594 (75.3) | 0.1 | 339264 (72.3) | 343493 (74.3) | 0.04 | 458923 (70.7) | 452526 (69.7) | 0.02 | 300465 (73.2) | 300863 (73.3) | 0.008 |
| Unknown | 130926 (20.2) | 595071 (20.6) | 0.01 | 80889 (17.2) | 76441 (16.5) | 0.02 | 130926 (20.2) | 134794 (20.8) | 0.01 | 69438 (16.9) | 69276 (16.9) | 0.001 |
| **Comorbidities** |  |  |  |  |  |  |  |  |  |  |  |  |
| Human immunodeficiency virus disease | 13168 (2.0) | 29842 (1.0) | 0.08 | 3728 (0.8) | 4328 (0.9) | 0.02 | 13111 (2.0) | 10633 (1.6) | 0.03 | 3069 (0.7) | 3955 (1.0) | 0.03 |
| Neoplasms | 209430 (32.2) | 1053786 (36.5) | 0.09 | 150215 (32.0) | 115483 (25.0) | 0.2 | 209328 (32.2) | 203635 (31.4) | 0.02 | 112002 (27.3) | 112584 (27.4) | 0.0004 |
| Diabetes mellitus | 131503 (20.2) | 551594 (19.1) | 0.03 | 87641 (18.7) | 69384 (15.0) | 0.1 | 131380 (20.2) | 126441 (19.5) | 0.02 | 66532 (16.2) | 66166 (16.1) | 0.0008 |
| Overweight and obesity | 168650 (26.0) | 649808 (22.5) | 0.08 | 122353 (26.1) | 94239 (20.4) | 0.1 | 168545 (26.0) | 168558 (26.0) | 0.00005 | 93203 (22.7) | 91581 (22.3) | 0.008 |
| Unspecified dementia | 7526 (1.2) | 26171 (0.9) | 0.03 | 4644 (1.0) | 2749 (0.6) | 0.04 | 7488 (1.2) | 6673 (1.0) | 0.01 | 2677 (0.7) | 2739 (0.7) | 0.007 |

#### **Supplementary Table 1 (continued).** Baseline demographics of COVID-19 compared to contemporary NCs and Influenza, before and after matching.

|  | **Before matching for baseline demographics and risk factors** | | | | | | **After matching for baseline demographics and risk factors ᵇ** | | | | | |
| --- | --- | --- | --- | --- | --- | --- | --- | --- | --- | --- | --- | --- |
| **Characteristics ᵃ** | COVID-19 | Contemporary negative controls |  | COVID-19 | Influenza |  | COVID-19 | Contemporary negative controls |  | COVID-19 | Influenza |  |
| Nicotine dependence | 106108 (16.3) | 349012 (12.1) | 0.1 | 61094 (13.0) | 58059 (12.6) | 0.01 | 105982 (16.3) | 102201 (15.7) | 0.02 | 51028 (12.4) | 51714 (12.6) | 0.003 |
| Paraplegia and quadriplegia | 3071 (0.5) | 11347 (0.4) | 0.01 | 2174 (0.5) | 1378 (0.3) | 0.03 | 3066 (0.5) | 3202 (0.5) | 0.003 | 1790 (0.4) | 1245 (0.3) | 0.01 |
| Essential hypertension | 255142 (39.3) | 1255731 (43.4) | 0.08 | 183361 (39.1) | 155563 (33.6) | 0.1 | 255027 (39.3) | 252325 (38.9) | 0.009 | 146996 (35.8) | 145812 (35.5) | 0.01 |
| Ischemic heart diseases | 86412 (13.3) | 397772 (13.8) | 0.01 | 59841 (12.8) | 44022 ( 9.5) | 0.1 | 86325 (13.3) | 82871 (12.8) | 0.02 | 42755 (10.4) | 42843 (10.4) | 0.007 |
| Acute myocardial infarction | 23812 (3.7) | 83162 (2.9) | 0.04 | 14273 (3.0) | 10019 (2.2) | 0.06 | 23760 (3.7) | 20137 (3.1) | 0.03 | 10064 (2.5) | 9755 (2.4) | 0.005 |
| Heart failure | 45610 (7.0) | 178405 (6.2) | 0.03 | 33873 (7.2) | 23580 (5.1) | 0.09 | 45569 (7.0) | 39677 (6.1) | 0.04 | 24002 (5.8) | 22774 (5.5) | 0.002 |
| Heart disease | 22735 (3.5) | 78118 (2.7) | 0.05 | 10448 (2.2) | 8584 (1.9) | 0.03 | 22620 (3.5) | 18776 (2.9) | 0.03 | 7840 (1.9) | 8006 (2.0) | 0.01 |
| Cerebral infarction | 34541 (5.3) | 104682 (3.6) | 0.08 | 15296 (3.3) | 9635 (2.1) | 0.07 | 34398 (5.3) | 30632 (4.7) | 0.03 | 9394 (2.3) | 9518 (2.3) | 0.01 |
| Peripheral vascular disease | 26907 (4.1) | 115164 (4.0) | 0.008 | 17919 (3.8) | 12122 (2.6) | 0.07 | 26892 (4.1) | 23978 (3.7) | 0.02 | 12591 (3.1) | 11750 (2.9) | 0.006 |
| Chronic lower respiratory diseases | 153849 (23.7) | 604552 (20.9) | 0.07 | 110410 (23.5) | 116497 (25.2) | 0.04 | 153732 (23.7) | 150162 (23.1) | 0.01 | 97564 (23.8) | 97402 (23.7) | 0.004 |
| Chronic obstructive pulmonary disease | 40766 (6.3) | 168054 (5.8) | 0.02 | 29734 (6.3) | 26202 (5.7) | 0.03 | 40747 (6.3) | 37089 (5.7) | 0.02 | 24035 (5.9) | 23916 (5.8) | 0.001 |
| Asthma | 95390 (14.7) | 353978 (12.2) | 0.07 | 66704 (14.2) | 70299 (15.2) | 0.03 | 95294 (14.7) | 92695 (14.3) | 0.01 | 59371 (14.5) | 59042 (14.4) | 0.003 |
| Peptic ulcer | 7097 (1.1) | 30666 (1.1) | 0.003 | 4680 (1.0) | 4021 (0.9) | 0.01 | 7087 (1.1) | 5924 (0.9) | 0.02 | 3618 (0.9) | 3761 (0.9) | 0.01 |
| Fibrosis and cirrhosis of liver | 12348 (1.9) | 43027 (1.5) | 0.03 | 8070 (1.7) | 4355 (0.9) | 0.07 | 12314 (1.9) | 10823 (1.7) | 0.02 | 4387 (1.1) | 4344 (1.1) | 0.01 |
| Portal hypertension | 4926 (0.8) | 15347 (0.5) | 0.03 | 3669 (0.8) | 1567 (0.3) | 0.06 | 4921 (0.8) | 3947 (0.6) | 0.02 | 2124 (0.5) | 1550 (0.4) | 0.01 |
| Psoriasis | 12755 (2.0) | 65580 (2.3) | 0.02 | 8896 (1.9) | 7394 (1.6) | 0.02 | 12750 (2.0) | 10742 (1.7) | 0.02 | 6824 (1.7) | 6975 (1.7) | 0.008 |
| Lupus erythematosus | 3624 (0.6) | 11871 (0.4) | 0.02 | 1953 (0.4) | 1506 (0.3) | 0.01 | 3616 (0.6) | 2755 (0.4) | 0.02 | 1406 (0.3) | 1418 (0.3) | 0.008 |
| Rheumatoid arthritis without rheumatoid factor | 1062 (0.2) | 6268 (0.2) | 0.01 | 821 (0.2) | 437 (0.09) | 0.02 | 1062 (0.2) | 1156 (0.2) | 0.004 | 673 (0.2) | 404 (0.1) | 0.01 |
| Rheumatoid arthritis | 12751 (2.0) | 61329 (2.1) | 0.01 | 8912 (1.9) | 7801 (1.7) | 0.02 | 12747 (2.0) | 10600 (1.6) | 0.02 | 7021 (1.7) | 7196 (1.8) | 0.009 |
| Acute kidney failure | 44492 (6.8) | 147813 (5.1) | 0.07 | 34492 (7.4) | 21138 (4.6) | 0.1 | 44462 (6.8) | 35923 (5.5) | 0.05 | 24224 (5.9) | 20425 (5.0) | 0.03 |
| Chronic kidney disease | 55055 (8.5) | 242791 (8.4) | 0.003 | 40414 (8.6) | 26452 (5.7) | 0.1 | 55014 (8.5) | 52336 (8.1) | 0.01 | 26039 (6.3) | 26134 (6.4) | 0.009 |
| End stage renal disease | 11409 (1.8) | 34423 (1.2) | 0.05 | 8956 (1.9) | 5713 (1.2) | 0.05 | 11392 (1.8) | 9329 (1.4) | 0.03 | 6058 (1.5) | 5626 (1.4) | 0.002 |

Abbreviations: SMD: standardized mean difference

ᵃ Patients were identified using *International Classification of Diseases and Related Health Problems, Tenth Revision* (*ICD-10*) and *Current Procedural Terminology (CPT)* codes. Characteristics were identified using electronic medical health record data from the TriNetX Research Network, and were recorded any time prior to the index event. Diagnostic coding can be found in the **online supplementary methods**.

ᵇ Matching covariates can be found in the **online supplementary** **methods**.

#### **Supplementary Table 2**: Baseline characteristics of COVID-19 compared to contemporary and historic negative controls, before matching.

|  | **Before matching for baseline demographics and risk factors** | | | | | | | | |
| --- | --- | --- | --- | --- | --- | --- | --- | --- | --- |
| **Characteristics ᵃ** | COVID-19 | Negative controls (Feb) |  | Negative controls (Jul) |  | Negative controls (Sept) |  | Negative controls (Nov) |  |
|  | No. (%) |  | SMD |  | SMD |  | SMD |  | SMD |
| **Number** | 652683 | 3122183 | - | 2890799 | - | 3039275 | - | 2326922 | - |
| **Age; mean (SD); y** | 49.2 (17.6) | 55.6 (17.3) | 0.4 | 55.7 (17.5) | 0.4 | 56.0 (17.3) | 0.4 | 56.1 (17.3) | 0.4 |
| **Sex** |  |  |  |  |  |  |  |  |  |
| Female | 401986 (61.6) | 1902589 (60.9) | 0.01 | 1775777 (61.4) | 0.001 | 1873397 (61.6) | 0.001 | 1433184 (61.6) | 3E-05 |
| Male | 250605 (38.4) | 1219346 (39.1) | 0.01 | 1114714 (38.6) | 0.001 | 1165644 (38.4) | 9E-04 | 893503 (38.4) | 5E-05 |
| **Race** |  |  |  |  |  |  |  |  |  |
| White | 456621 (70.0) | 2244924 (71.9) | 0.06 | 2097083 (72.5) | 0.07 | 2211281 (72.8) | 0.06 | 1697375 (72.9) | 0.07 |
| Black or African American | 115047 (17.6) | 428230 (13.7) | 0.1 | 386394 (13.4) | 0.1 | 406218 (13.4) | 0.1 | 299888 (12.9) | 0.1 |
| **Ethnicity** |  |  |  |  |  |  |  |  |  |
| Hispanic or Latino | 54879 (8.4) | 142140 (4.6) | 0.2 | 119134 (4.1) | 0.2 | 126472 (4.2) | 0.2 | 94171 (4.0) | 0.2 |
| Not Hispanic of Latino | 466645 (71.5) | 2335223 (74.8) | 0.09 | 2176594 (75.3) | 0.1 | 2322316 (76.4) | 0.1 | 1695475 (72.9) | 0.03 |
| Unknown | 131159 (20.1) | 644820 (20.7) | 0.01 | 595071 (20.6) | 0.01 | 590487 (19.4) | 0.02 | 537276 (23.1) | 0.07 |
| **Comorbidities** |  |  |  |  |  |  |  |  |  |
| Human immunodeficiency virus disease | 14261 (2.2) | 33486 (1.1) | 0.09 | 29842 (1.0) | 0.08 | 32513 (1.1) | 0.09 | 27576 (1.2) | 0.08 |
| Neoplasms | 213542 (32.7) | 1105384 (35.4) | 0.06 | 1053786 (36.5) | 0.09 | 1133154 (37.3) | 0.1 | 854544 (36.7) | 0.08 |
| Diabetes mellitus | 134086 (20.5) | 596808 (19.1) | 0.04 | 551594 (19.1) | 0.03 | 583154 (19.2) | 0.03 | 464603 (20.0) | 0.01 |
| Overweight and obesity | 171567 (26.3) | 695329 (22.3) | 0.09 | 649808 (22.5) | 0.08 | 689666 (22.7) | 0.08 | 539977 (23.2) | 0.07 |
| Unspecified dementia | 7808 (1.2) | 29086 (0.9) | 0.03 | 26171 (0.9) | 0.03 | 28399 (0.9) | 0.03 | 22996 (1.0) | 0.02 |
| Nicotine dependence | 109488 (16.8) | 369975 (11.8) | 0.1 | 349012 (12.1) | 0.1 | 361110 (11.9) | 0.1 | 292839 (12.6) | 0.1 |
| Paraplegia and quadriplegia | 3035 (0.5) | 12357 (0.4) | 0.01 | 11347 (0.4) | 0.01 | 11452 (0.4) | 0.01 | 9535 (0.4) | 0.008 |
| Essential hypertension | 257416 (39.4) | 1342990 (43.0) | 0.07 | 1255731 (43.4) | 0.08 | 1328534 (43.7) | 0.09 | 1028817 (44.2) | 0.1 |

#### **Supplementary Table 2 (continued)**: Baseline characteristics of COVID-19 compared to contemporary and historic negative controls, before matching.

| **Characteristics ᵃ** | COVID-19 | Negative controls (Feb) |  | Negative controls (Jul) |  | Negative controls (Sept) |  | Negative controls (Nov) |  |
| --- | --- | --- | --- | --- | --- | --- | --- | --- | --- |
|  | No. (%) |  | SMD |  | SMD |  | SMD |  | SMD |
| Ischemic heart diseases | 88480 (13.6) | 423248 (13.6) | 5E-06 | 397772 (13.8) | 0.01 | 423061 (13.9) | 0.01 | 328586 (14.1) | 0.02 |
| Acute myocardial infarction | 25017 (3.8) | 88963 (2.8) | 0.05 | 83162 (2.9) | 0.04 | 88402 (2.9) | 0.05 | 70257 (3.0) | 0.04 |
| Heart failure | 46271 (7.1) | 193379 (6.2) | 0.04 | 178405 (6.2) | 0.03 | 189606 (6.2) | 0.03 | 148471 (6.4) | 0.03 |
| Heart disease | 23784 (3.6) | 81443 (2.6) | 0.06 | 78118 (2.7) | 0.05 | 82579 (2.7) | 0.05 | 62444 (2.7) | 0.05 |
| Cerebral infarction | 37238 (5.7) | 113693 (3.6) | 0.09 | 104682 (3.6) | 0.08 | 113710 (3.7) | 0.09 | 91495 (3.9) | 0.08 |
| Peripheral vascular disease | 28805 (4.4) | 124239 (4.0) | 0.02 | 115164 (4.0) | 0.008 | 127003 (4.2) | 0.01 | 99668 (4.3) | 0.006 |
| Chronic lower respiratory diseases | 156589 (24.0) | 638822 (20.5) | 0.08 | 604552 (20.9) | 0.07 | 636394 (20.9) | 0.07 | 492059 (21.1) | 0.07 |
| Chronic obstructive pulmonary disease | 41927 (6.4) | 178577 (5.7) | 0.02 | 168054 (5.8) | 0.02 | 174985 (5.8) | 0.03 | 140549 (6.0) | 0.02 |
| Asthma | 96906 (14.8) | 370663 (11.9) | 0.09 | 353978 (12.2) | 0.07 | 379056 (12.5) | 0.07 | 287305 (12.3) | 0.07 |
| Peptic ulcer | 7203 (1.1) | 31503 (1.0) | 0.009 | 30666 (1.1) | 0.003 | 31999 (1.1) | 0.005 | 26211 (1.1) | 0.002 |
| Fibrosis and cirrhosis of liver | 12561 (1.9) | 45780 (1.5) | 0.04 | 43027 (1.5) | 0.03 | 44923 (1.5) | 0.03 | 35516 (1.5) | 0.03 |
| Portal hypertension | 4638 (0.7) | 16280 (0.5) | 0.03 | 15347 (0.5) | 0.03 | 15666 (0.5) | 0.03 | 12175 (0.5) | 0.02 |
| Psoriasis | 13186 (2.0) | 68158 (2.2) | 0.01 | 65580 (2.3) | 0.02 | 70592 (2.3) | 0.02 | 53725 (2.3) | 0.02 |
| Lupus erythematosus | 3794 (0.6) | 12894 (0.4) | 0.03 | 11871 (0.4) | 0.02 | 12659 (0.4) | 0.02 | 10083 (0.4) | 0.02 |
| Rheumatoid arthritis without rheumatoid factor | 1060 (0.2) | 6479 (0.2) | 0.01 | 6268 (0.2) | 0.01 | 6685 (0.2) | 0.01 | 5522 (0.2) | 0.02 |
| Rheumatoid arthritis | 12827 (2.0) | 64136 (2.1) | 0.005 | 61329 (2.1) | 0.01 | 63478 (2.1) | 0.009 | 50287 (2.2) | 0.01 |
| Acute kidney failure | 43308 (6.6) | 156480 (5.0) | 0.08 | 147813 (5.1) | 0.07 | 154983 (5.1) | 0.07 | 120213 (5.2) | 0.06 |
| Chronic kidney disease | 58779 (9.0) | 257759 (8.3) | 0.01 | 242791 (8.4) | 0.003 | 261316 (8.6) | 0.01 | 210960 (9.1) | 0.002 |
| End stage renal disease | 15756 (2.4) | 37714 (1.2) | 0.05 | 34423 (1.2) | 0.05 | 41474 (1.4) | 0.08 | 35479 (1.5) | 0.06 |

Abbreviations: SMD: standardized mean difference

ᵃ Patients were identified using *International Classification of Diseases and Related Health Problems, Tenth Revision* (*ICD-10*) and *Current Procedural Terminology (CPT)* codes. Characteristics were identified using electronic medical health record data from the TriNetX Research Network, and were recorded any time prior to the index event. Diagnostic coding can be found in the **online supplementary methods**.

#### **Supplementary Table 3**: Baseline characteristics of COVID-19 compared to contemporary and historic negative controls, after matching.

|  | **After matching for baseline demographics and risk factors** | | | | | | | | | | | |
| --- | --- | --- | --- | --- | --- | --- | --- | --- | --- | --- | --- | --- |
| **Characteristics ᵃ** | COVID-19 | Negative control (Feb) |  | COVID-19 | Negative control (Jul) |  | COVID-19 | Negative control (Sept) |  | COVID-19 | Negative control (Nov) |  |
|  | No. (%) |  | SMD | No. (%) |  | SMD | No. (%) |  | SMD | No. (%) |  | SMD |
| **Number** | 649714 | 649714 | - | 649311 | 649311 | - | 652240 | 652240 | - | 652086 | 652086 | - |
| **Age; mean (SD); y** | 49.2 (17.6) | 49.1 (17.5) | 0.008 | 49.2 (17.6) | 49.1 (17.5) | 0.005 | 49.2 (17.6) | 49.2 (17.6) | 0.003 | 49.2 (17.6) | 49.1 (17.6) | 0.007 |
| **Sex** |  |  |  |  |  |  |  |  |  |  |  |  |
| Female | 399497 (61.5) | 395906 (60.9) | 0.01 | 399302 (61.5) | 395457 (60.9) | 0.01 | 401712 (61.6) | 397942 (61.0) | 0.01 | 401621 (61.6) | 397951 (61.0) | 0.01 |
| Male | 250123 (38.5) | 253731 (39.1) | 0.01 | 249915 (38.5) | 253764 (39.1) | 0.01 | 250436 (38.4) | 254211 (39.0) | 0.01 | 250373 (38.4) | 254044 (39.0) | 0.01 |
| **Race** |  |  |  |  |  |  |  |  |  |  |  |  |
| White | 450600 (69.4) | 447963 (68.9) | 0.009 | 450253 (69.3) | 446495 (68.8) | 0.01 | 456360 (70.0) | 454022 (69.6) | 0.008 | 456327 (70.0) | 452972 (69.5) | 0.01 |
| Black or African American | 117280 (18.1) | 120904 (18.6) | 0.01 | 117247 (18.1) | 121390 (18.7) | 0.02 | 114945 (17.6) | 118015 (18.1) | 0.01 | 114857 (17.6) | 118774 (18.2) | 0.02 |
| **Ethnicity** |  |  |  |  |  |  |  |  |  |  |  |  |
| Hispanic or Latino | 59414 (9.1) | 62010 ( 9.5) | 0.01 | 59462 (9.2) | 61991 ( 9.5) | 0.01 | 54652 (8.4) | 57573 (8.8) | 0.02 | 54535 (8.4) | 57301 (8.8) | 0.02 |
| Not Hispanic of Latino | 459358 (70.7) | 453175 (69.8) | 0.02 | 458923 (70.7) | 452526 (69.7) | 0.02 | 466436 (71.5) | 459213 (70.4) | 0.02 | 466394 (71.5) | 459403 (70.5) | 0.02 |
| Unknown | 130942 (20.2) | 134529 (20.7) | 0.01 | 130926 (20.2) | 134794 (20.8) | 0.01 | 131152 (20.1) | 135454 (20.8) | 0.02 | 131157 (20.1) | 135382 (20.8) | 0.02 |
| **Comorbidities** |  |  |  |  |  |  |  |  |  |  |  |  |
| Human immunodeficiency virus disease | 13907 (2.1) | 10839 (1.7) | 0.03 | 13111 (2.0) | 10633 (1.6) | 0.03 | 14118 (2.2) | 10602 (1.6) | 0.04 | 14099 (2.2) | 11905 (1.8) | 0.02 |
| Neoplasms | 211765 (32.6) | 206155 (31.7) | 0.02 | 209328 (32.2) | 203635 (31.4) | 0.02 | 213247 (32.7) | 207272 (31.8) | 0.02 | 213200 (32.7) | 207831 (31.9) | 0.02 |
| Diabetes mellitus | 133521 (20.6) | 129153 (19.9) | 0.02 | 131380 (20.2) | 126441 (19.5) | 0.02 | 133734 (20.5) | 129710 (19.9) | 0.02 | 133681 (20.5) | 128679 (19.7) | 0.02 |
| Overweight and obesity | 169393 (26.1) | 169401 (26.1) | 3E-05 | 168545 (26.0) | 168558 (26.0) | 5E-05 | 171290 (26.3) | 171382 (26.3) | 3E-04 | 171238 (26.3) | 170373 (26.1) | 0.003 |
| Unspecified dementia | 7655 (1.2) | 6870 (1.1) | 0.01 | 7488 (1.2) | 6673 (1.0) | 0.01 | 7715 (1.2) | 6953 (1.1) | 0.01 | 7709 (1.2) | 7171 (1.1) | 0.008 |
| Nicotine dependence | 106989 (16.5) | 103481 (15.9) | 0.01 | 105982 (16.3) | 102201 (15.7) | 0.02 | 109139 (16.7) | 105244 (16.1) | 0.02 | 109090 (16.7) | 103911 (15.9) | 0.02 |
| Paraplegia and quadriplegia | 3135 (0.5) | 3236 (0.5) | 0.002 | 3066 (0.5) | 3202 (0.5) | 0.003 | 3023 (0.5) | 3145 (0.5) | 0.003 | 3022 (0.5) | 3412 (0.5) | 0.009 |
| Essential hypertension | 255930 (39.4) | 253289 (39.0) | 0.008 | 255027 (39.3) | 252325 (38.9) | 0.009 | 257090 (39.4) | 254953 (39.1) | 0.007 | 257040 (39.4) | 253690 (38.9) | 0.01 |

#### **Supplementary Table 3 (continued)**: Baseline characteristics of COVID-19 compared to contemporary and historic negative controls, after matching.

| **Characteristics ᵃ** | COVID-19 | Negative control (Feb) |  | COVID-19 | Negative control (Jul) |  | COVID-19 | Negative control (Sept) |  | COVID-19 | Negative control (Nov) |  |
| --- | --- | --- | --- | --- | --- | --- | --- | --- | --- | --- | --- | --- |
|  | No. (%) |  | SMD | No. (%) |  | SMD | No. (%) |  | SMD | No. (%) |  | SMD |
| Ischemic heart diseases | 87927 (13.5) | 84438 (13.0) | 0.02 | 86325 (13.3) | 82871 (12.8) | 0.02 | 88250 (13.5) | 85813 (13.2) | 0.01 | 88211 (13.5) | 84648 (13.0) | 0.02 |
| Acute myocardial infarction | 24306 (3.7) | 20656 (3.2) | 0.03 | 23760 (3.7) | 20137 (3.1) | 0.03 | 24883 (3.8) | 20918 (3.2) | 0.03 | 24863 (3.8) | 20955 (3.2) | 0.03 |
| Heart failure | 46385 (7.1) | 40900 (6.3) | 0.03 | 45569 (7.0) | 39677 (6.1) | 0.04 | 46130 (7.1) | 41212 (6.3) | 0.03 | 46123 (7.1) | 40157 (6.2) | 0.04 |
| Heart disease | 23056 (3.5) | 19344 (3.0) | 0.03 | 22620 (3.5) | 18776 (2.9) | 0.03 | 23497 (3.6) | 20310 (3.1) | 0.03 | 23449 (3.6) | 19692 (3.0) | 0.03 |
| Cerebral infarction | 36143 (5.6) | 33039 (5.1) | 0.02 | 34398 (5.3) | 30632 (4.7) | 0.03 | 36854 (5.7) | 33983 (5.2) | 0.02 | 36793 (5.6) | 33681 (5.2) | 0.02 |
| Peripheral vascular disease | 27878 (4.3) | 24714 (3.8) | 0.02 | 26892 (4.1) | 23978 (3.7) | 0.02 | 28747 (4.4) | 26076 (4.0) | 0.02 | 28749 (4.4) | 25383 (3.9) | 0.03 |
| Chronic lower respiratory diseases | 154688 (23.8) | 151366 (23.3) | 0.01 | 153732 (23.7) | 150162 (23.1) | 0.01 | 156273 (24.0) | 153416 (23.5) | 0.01 | 156196 (24.0) | 152757 (23.4) | 0.01 |
| Chronic obstructive pulmonary disease | 40959 (6.3) | 37904 (5.8) | 0.02 | 40747 (6.3) | 37089 (5.7) | 0.02 | 41858 (6.4) | 38973 (6.0) | 0.02 | 41852 (6.4) | 38189 (5.9) | 0.02 |
| Asthma | 95960 (14.8) | 93421 (14.4) | 0.01 | 95294 (14.7) | 92695 (14.3) | 0.01 | 96661 (14.8) | 93784 (14.4) | 0.01 | 96589 (14.8) | 93959 (14.4) | 0.01 |
| Peptic ulcer | 7151 (1.1) | 5906 (0.9) | 0.02 | 7087 (1.1) | 5924 (0.9) | 0.02 | 7172 (1.1) | 6104 (0.9) | 0.02 | 7168 (1.1) | 5888 (0.9) | 0.02 |
| Fibrosis and cirrhosis of liver | 12703 (2.0) | 11115 (1.7) | 0.02 | 12314 (1.9) | 10823 (1.7) | 0.02 | 12480 (1.9) | 10851 (1.7) | 0.02 | 12478 (1.9) | 10591 (1.6) | 0.02 |
| Portal hypertension | 4944 (0.8) | 4019 (0.6) | 0.02 | 4921 (0.8) | 3947 (0.6) | 0.02 | 4616 (0.7) | 3791 (0.6) | 0.02 | 4618 (0.7) | 3576 (0.5) | 0.02 |
| Psoriasis | 12903 (2.0) | 10948 (1.7) | 0.02 | 12750 (2.0) | 10742 (1.7) | 0.02 | 13177 (2.0) | 11139 (1.7) | 0.02 | 13173 (2.0) | 11334 (1.7) | 0.02 |
| Lupus erythematosus | 3804 (0.6) | 3008 (0.5) | 0.02 | 3616 (0.6) | 2755 (0.4) | 0.02 | 3763 (0.6) | 2802 (0.4) | 0.02 | 3754 (0.6) | 3005 (0.5) | 0.02 |
| Rheumatoid arthritis without rheumatoid factor | 1062 (0.2) | 1098 (0.2) | 0.001 | 1062 (0.2) | 1156 (0.2) | 0.004 | 1059 (0.2) | 1129 (0.2) | 0.003 | 1059 (0.2) | 1264 (0.2) | 0.007 |
| Rheumatoid arthritis | 12851 (2.0) | 10687 (1.6) | 0.02 | 12747 (2.0) | 10600 (1.6) | 0.02 | 12803 (2.0) | 10879 (1.7) | 0.02 | 12803 (2.0) | 10881 (1.7) | 0.02 |
| Acute kidney failure | 44474 (6.8) | 35842 (5.5) | 0.06 | 44462 (6.8) | 35923 (5.5) | 0.05 | 43197 (6.6) | 36826 (5.6) | 0.04 | 43186 (6.6) | 35495 (5.4) | 0.05 |
| Chronic kidney disease | 55354 (8.5) | 52819 (8.1) | 0.01 | 55014 (8.5) | 52336 (8.1) | 0.01 | 58642 (9.0) | 56554 (8.7) | 0.01 | 58628 (9.0) | 55747 (8.5) | 0.02 |
| End stage renal disease | 11407 (1.8) | 9662 (1.5) | 0.02 | 11392 (1.8) | 9329 (1.4) | 0.03 | 15698 (2.4) | 10903 (1.7) | 0.05 | 15692 (2.4) | 11636 (1.8) | 0.04 |

Abbreviations: SMD: standardized mean difference

ᵃ Patients were identified using *International Classification of Diseases and Related Health Problems, Tenth Revision* (*ICD-10*) and *Current Procedural Terminology (CPT)* codes. Characteristics were identified using electronic medical health record data from the TriNetX Research Network, and were recorded any time prior to the index event. Diagnostic coding can be found in the **online supplementary methods**.

#### **Supplementary Table 4**: Baseline characteristics of COVID-19 compared to other infectious diseases, before matching.

|  | **Before matching for baseline demographics and risk factors** | | | | | | | | | | | | | | |
| --- | --- | --- | --- | --- | --- | --- | --- | --- | --- | --- | --- | --- | --- | --- | --- |
| **Characteristics ᵃ** | COVID-19 | Lymes disease |  | COVID-19 | Herpes zoster |  | COVID-19 | Varicella Zoster |  | COVID-19 | Infectious mononucleosis |  | COVID-19 | Cytomegalovirus |  |
|  | No. (%) |  | SMD | No. (%) |  | SMD | No. (%) |  | SMD | No. (%) |  | SMD | No. (%) |  | SMD |
| **Number** | 649955 | 92989 | - | 649952 | 390439 | - | 643636 | 66790 | - | 649956 | 44243 | - | 649478 | 6503 | - |
| **Age; mean (SD); y** | 49.2 (17.6) | 52.1 (16.3) | 0.2 | 49.2 (17.6) | 57.4 (15.5) | 0.5 | 49.2 (17.6) | 41.0 (17.7) | 0.5 | 49.2 (17.6) | 30.6 (15.1) | 1.1 | 49.2 (17.6) | 53.8 (14.5) | 0.3 |
| **Sex** |  |  |  |  |  |  |  |  |  |  |  |  |  |  |  |
| Female | 399674 (61.5) | 50806 (54.6) | 0.1 | 399671 (61.5) | 249368 (63.9) | 0.05 | 395359 (61.4) | 46823 (70.1) | 0.2 | 399674 (61.5) | 28158 (63.6) | 0.04 | 399408 (61.5) | 2808 (43.2) | 0.4 |
| Male | 250187 (38.5) | 42179 (45.4) | 0.1 | 250187 (38.5) | 141048 (36.1) | 0.05 | 248183 (38.6) | 19964 (29.9) | 0.2 | 250188 (38.5) | 16077 (36.3) | 0.04 | 249976 (38.5) | 3695 (56.8) | 0.4 |
| **Race** |  |  |  |  |  |  |  |  |  |  |  |  |  |  |  |
| White | 450707 (69.3) | 79166 (85.1) | 0.4 | 450704 (69.3) | 302851 (77.6) | 0.2 | 446992 (69.4) | 50289 (75.3) | 0.1 | 450708 (69.3) | 36607 (82.7) | 0.3 | 450333 (69.3) | 3754 (57.7) | 0.2 |
| Black or African American | 117326 (18.1) | 2102 (2.3) | 0.5 | 117326 (18.1) | 41460 (10.6) | 0.2 | 115540 (18.0) | 8384 (12.6) | 0.2 | 117326 (18.1) | 2586 (5.8) | 0.4 | 117279 (18.1) | 1692 (26.0) | 0.2 |
| **Ethnicity** |  |  |  |  |  |  |  |  |  |  |  |  |  |  |  |
| Hispanic or Latino | 59618 (9.2) | 1575 (1.7) | 0.3 | 59618 (9.2) | 23501 (6.0) | 0.1 | 58932 (9.2) | 4137 (6.2) | 0.1 | 59618 (9.2) | 1666 (3.8) | 0.2 | 59593 (9.2) | 563 (8.7) | 0.02 |
| Not Hispanic of Latino | 459398 (70.7) | 76766 (82.6) | 0.3 | 459395 (70.7) | 294142 (75.3) | 0.1 | 454085 (70.5) | 58648 (87.8) | 0.4 | 459399 (70.7) | 35347 (79.9) | 0.2 | 458959 (70.7) | 5151 (79.2) | 0.2 |
| Unknown | 130939 (20.1) | 14648 (15.8) | 0.1 | 130939 (20.1) | 72796 (18.6) | 0.04 | 130619 (20.3) | 4005 (6.0) | 0.4 | 130939 (20.1) | 7230 (16.3) | 0.1 | 130926 (20.2) | 789 (12.1) | 0.2 |
| **Comorbidities** |  |  |  |  |  |  |  |  |  |  |  |  |  |  |  |
| Human immunodeficiency virus disease | 14033 (2.2) | 295 (0.3) | 0.2 | 14033 (2.2) | 7363 (1.9) | 0.02 | 14024 (2.2) | 2203 (3.3) | 0.07 | 14033 (2.2) | 284 (0.6) | 0.1 | 13168 (2.0) | 440 (6.8) | 0.2 |
| Neoplasms | 212029 (32.6) | 26073 (28.0) | 0.1 | 212029 (32.6) | 138897 (35.6) | 0.06 | 210814 (32.8) | 21814 (32.7) | 0.002 | 212029 (32.6) | 7195 (16.3) | 0.4 | 209430 (32.2) | 3302 (50.8) | 0.4 |
| Diabetes mellitus | 133821 (20.6) | 8326 (9.0) | 0.3 | 133821 (20.6) | 73539 (18.8) | 0.04 | 132829 (20.6) | 8773 (13.1) | 0.2 | 133821 (20.6) | 2400 (5.4) | 0.5 | 131503 (20.2) | 3366 (51.8) | 0.7 |
| Overweight and obesity | 169621 (26.1) | 10104 (10.9) | 0.4 | 169621 (26.1) | 70023 (17.9) | 0.2 | 167987 (26.1) | 10647 (15.9) | 0.3 | 169621 (26.1) | 3539 (8.0) | 0.5 | 168650 (26.0) | 2062 (31.7) | 0.1 |
| Unspecified dementia | 7740 (1.2) | 319 (0.3) | 0.1 | 7740 (1.2) | 3708 (0.9) | 0.02 | 7679 (1.2) | 557 (0.8) | 0.04 | 7740 (1.2) | 44 (0.1) | 0.1 | 7526 (1.2) | 47 (0.7) | 0.05 |
| Nicotine dependence | 107271 (16.5) | 8159 (8.8) | 0.2 | 107271 (16.5) | 44024 (11.3) | 0.2 | 106682 (16.6) | 11099 (16.6) | 0.001 | 107271 (16.5) | 2733 (6.2) | 0.3 | 106108 (16.3) | 892 (13.7) | 0.07 |
| Paraplegia and quadriplegia | 3146 (0.5) | 114 (0.1) | 0.07 | 3146 (0.5) | 1129 (0.3) | 0.03 | 3139 (0.5) | 188 (0.3) | 0.03 | 3146 (0.5) | 59 (0.1) | 0.06 | 3071 (0.5) | 71 (1.1) | 0.07 |
| Essential hypertension | 256173 (39.4) | 25115 (27.0) | 0.3 | 256174 (39.4) | 168718 (43.2) | 0.08 | 254118 (39.5) | 16400 (24.6) | 0.3 | 256174 (39.4) | 5106 (11.5) | 0.7 | 255142 (39.3) | 4961 (76.3) | 0.8 |
| Ischemic heart diseases | 88126 (13.6) | 7277 (7.8) | 0.2 | 88127 (13.6) | 51410 (13.2) | 0.01 | 87741 (13.6) | 4759 (7.1) | 0.2 | 88127 (13.6) | 1487 (3.4) | 0.4 | 86412 (13.3) | 2504 (38.5) | 0.6 |

#### **Supplementary Table 4 (continued)**: Baseline characteristics of COVID-19 compared to other infectious diseases, before matching.

| **Characteristics ᵃ** | COVID-19 | Lymes disease |  | COVID-19 | Herpes zoster |  | COVID-19 | Varicella Zoster |  | COVID-19 | Infectious mononucleosis |  | COVID-19 | Cytomegalovirus |  |
| --- | --- | --- | --- | --- | --- | --- | --- | --- | --- | --- | --- | --- | --- | --- | --- |
|  | No. (%) |  | SMD | No. (%) |  | SMD | No. (%) |  | SMD | No. (%) |  | SMD | No. (%) |  | SMD |
| Acute myocardial infarction | 24411 (3.8) | 1267 (1.4) | 0.2 | 24411 (3.8) | 10647 (2.7) | 0.06 | 24345 (3.8) | 1307 (2.0) | 0.1 | 24411 (3.8) | 296 (0.7) | 0.2 | 23812 (3.7) | 603 (9.3) | 0.2 |
| Heart failure | 46490 (7.2) | 2152 (2.3) | 0.2 | 46491 (7.2) | 23158 (5.9) | 0.05 | 46310 (7.2) | 2105 (3.2) | 0.2 | 46491 (7.2) | 755 (1.7) | 0.3 | 45610 (7.0) | 1897 (29.2) | 0.6 |
| Heart disease | 23306 (3.6) | 1299 (1.4) | 0.1 | 23307 (3.6) | 10694 (2.7) | 0.05 | 23230 (3.6) | 2771 (4.1) | 0.03 | 23307 (3.6) | 380 (0.9) | 0.2 | 22735 (3.5) | 588 (9.0) | 0.2 |
| Cerebral infarction | 36484 (5.6) | 1618 (1.7) | 0.2 | 36485 (5.6) | 15034 (3.9) | 0.08 | 36389 (5.7) | 3755 (5.6) | 0.001 | 36485 (5.6) | 431 (1.0) | 0.3 | 34541 (5.3) | 520 (8.0) | 0.1 |
| Peripheral vascular disease | 27928 (4.3) | 1599 (1.7) | 0.2 | 27929 (4.3) | 15059 (3.9) | 0.02 | 27830 (4.3) | 1396 (2.1) | 0.1 | 27929 (4.3) | 363 (0.8) | 0.2 | 26907 (4.1) | 641 ( 9.9) | 0.2 |
| Chronic lower respiratory diseases | 154953 (23.8) | 14451 (15.5) | 0.2 | 154954 (23.8) | 83849 (21.5) | 0.06 | 153744 (23.9) | 11993 (18.0) | 0.1 | 154954 (23.8) | 6893 (15.6) | 0.2 | 153849 (23.7) | 1923 (29.6) | 0.1 |
| Chronic obstructive pulmonary disease | 41021 (6.3) | 2799 (3.0) | 0.2 | 41021 (6.3) | 24975 (6.4) | 0.003 | 40787 (6.3) | 1886 (2.8) | 0.2 | 41021 (6.3) | 570 (1.3) | 0.3 | 40766 (6.3) | 810 (12.5) | 0.2 |
| Asthma | 96188 (14.8) | 8239 (8.9) | 0.2 | 96188 (14.8) | 45743 (11.7) | 0.09 | 95366 (14.8) | 8526 (12.8) | 0.06 | 96188 (14.8) | 4906 (11.1) | 0.1 | 95390 (14.7) | 836 (12.9) | 0.05 |
| Peptic ulcer | 7180 (1.1) | 595 (0.6) | 0.05 | 7180 (1.1) | 4339 (1.1) | 6E-04 | 7147 (1.1) | 458 (0.7) | 0.05 | 7180 (1.1) | 133 (0.3) | 0.1 | 7097 (1.1) | 167 (2.6) | 0.1 |
| Fibrosis and cirrhosis of liver | 12799 (2.0) | 528 (0.6) | 0.1 | 12799 (2.0) | 5263 (1.3) | 0.05 | 12770 (2.0) | 1116 (1.7) | 0.02 | 12799 (2.0) | 406 (0.9) | 0.09 | 12348 (1.9) | 861 (13.2) | 0.4 |
| Portal hypertension | 4963 (0.8) | 143 (0.2) | 0.09 | 4963 (0.8) | 2047 (0.5) | 0.03 | 4947 (0.8) | 406 (0.6) | 0.02 | 4963 (0.8) | 207 (0.5) | 0.04 | 4926 (0.8) | 596 (9.2) | 0.4 |
| Psoriasis | 12913 (2.0) | 1622 (1.7) | 0.02 | 12913 (2.0) | 8377 (2.1) | 0.01 | 12835 (2.0) | 1192 (1.8) | 0.02 | 12913 (2.0) | 401 (0.9) | 0.09 | 12755 (2.0) | 114 (1.8) | 0.02 |
| Lupus erythematosus | 3833 (0.6) | 220 (0.2) | 0.06 | 3833 (0.6) | 2387 (0.6) | 0.003 | 3824 (0.6) | 462 (0.7) | 0.01 | 3833 (0.6) | 100 (0.2) | 0.06 | 3624 (0.6) | 107 (1.6) | 0.1 |
| Rheumatoid arthritis without rheumatoid factor | 1062 (0.2) | 75 (0.08) | 0.02 | 1062 (0.2) | 610 (0.2) | 0.002 | 1052 (0.2) | 48 (0.07) | 0.03 | 1062 (0.2) | 10 (0.02) | 0.05 | 1062 (0.2) | 10 (0.2) | 0.002 |
| Rheumatoid arthritis | 12878 (2.0) | 1419 (1.5) | 0.03 | 12878 (2.0) | 11418 (2.9) | 0.06 | 12781 (2.0) | 1012 (1.5) | 0.04 | 12878 (2.0) | 340 (0.8) | 0.1 | 12751 (2.0) | 180 (2.8) | 0.05 |
| Acute kidney failure | 44557 (6.9) | 1503 (1.6) | 0.3 | 44558 (6.9) | 19331 (5.0) | 0.08 | 44369 (6.9) | 2164 (3.2) | 0.2 | 44558 (6.9) | 1288 (2.9) | 0.2 | 44492 (6.8) | 3513 (54.0) | 1.2 |
| Chronic kidney disease | 55467 (8.5) | 2725 (2.9) | 0.2 | 55468 (8.5) | 31432 (8.1) | 0.02 | 55183 (8.6) | 2940 (4.4) | 0.2 | 55468 (8.5) | 1546 (3.5) | 0.2 | 55055 (8.5) | 4204 (64.6) | 1.4 |
| End stage renal disease | 11447 (1.8) | 229 (0.2) | 0.2 | 11447 (1.8) | 5749 (1.5) | 0.02 | 11405 (1.8) | 698 (1.0) | 0.06 | 11447 (1.8) | 815 (1.8) | 0.006 | 11409 (1.8) | 2826 (43.5) | 1.1 |

Abbreviations: SMD: standardized mean difference

ᵃ Patients were identified using *International Classification of Diseases and Related Health Problems, Tenth Revision* (*ICD-10*) and *Current Procedural Terminology (CPT)* codes. Characteristics were identified using electronic medical health record data from the TriNetX Research Network, and were recorded any time prior to the index event. Diagnostic coding can be found in the **online supplementary methods**.

#### **Supplementary Table 5**: Baseline characteristics of COVID-19 compared to other infectious diseases, after matching.

|  | **After matching for baseline demographics and risk factorsᵉ** | | | | | | | | | | | | | | |
| --- | --- | --- | --- | --- | --- | --- | --- | --- | --- | --- | --- | --- | --- | --- | --- |
| **Characteristics ᵃ** | COVID-19 | Lymes disease |  | COVID-19 | Herpes zoster | SMD | COVID-19 | Varicella Zoster |  | COVID-19 | Infectious mononucleosis |  | COVID-19 | Cytomegalovirus |  |
|  | No. (%) |  | SMD | No. (%) |  | SMD | No. (%) |  | SMD | No. (%) |  | SMD | No. (%) |  | SMD |
| **Number** | 92989 | 92989 | - | 359377 | 359377 | - | 66785 | 66785 | - | 43553 | 43553 | - | 6499 | 6499 | - |
| **Age; mean (SD); y** | 51.9 (16.4) | 52.1 (16.3) | 0.008 | 56.4 (15.9) | 56.1 (15.3) | 0.02 | 41.2 (17.5) | 41.1 (17.7) | 0.007 | 30.8 (15.1) | 30.8 (15.2) | 0.001 | 55.3 (15.3) | 53.9 (14.5) | 0.1 |
| **Sex** |  |  |  |  |  |  |  |  |  |  |  |  |  |  |  |
| Female | 51044 (54.9) | 50806 (54.6) | 0.005 | 217973 (60.7) | 226255 (63.0) | 0.05 | 46759 (70.0) | 46818 (70.1) | 0.002 | 27831 (63.9) | 27855 (64.0) | 0.001 | 2732 (42.0) | 2808 (43.2) | 0.02 |
| Male | 41937 (45.1) | 42179 (45.4) | 0.005 | 141372 (39.3) | 133099 (37.0) | 0.05 | 20017 (30.0) | 19964 (29.9) | 0.002 | 15716 (36.1) | 15690 (36.0) | 0.001 | 3767 (58.0) | 3691 (56.8) | 0.02 |
| **Race** |  |  |  |  |  |  |  |  |  |  |  |  |  |  |  |
| White | 79410 (85.4) | 79166 (85.1) | 0.007 | 278283 (77.4) | 275375 (76.6) | 0.02 | 50512 (75.6) | 50286 (75.3) | 0.008 | 36165 (83.0) | 35919 (82.5) | 0.01 | 3752 (57.7) | 3753 (57.7) | 3E-04 |
| Black or African American | 2202 (2.4) | 2102 (2.3) | 0.007 | 42058 (11.7) | 41163 (11.5) | 0.008 | 8341 (12.5) | 8384 (12.6) | 0.002 | 2636 (6.1) | 2586 (5.9) | 0.005 | 1849 (28.5) | 1692 (26.0) | 0.05 |
| **Ethnicity** |  |  |  |  |  |  |  |  |  |  |  |  |  |  |  |
| Hispanic or Latino | 1619 (1.7) | 1575 (1.7) | 0.004 | 22643 (6.3) | 23180 (6.5) | 0.006 | 3886 (5.8) | 4137 (6.2) | 0.02 | 1610 (3.7) | 1666 (3.8) | 0.007 | 511 (7.9) | 562 (8.6) | 0.03 |
| Not Hispanic of Latino | 76853 (82.6) | 76766 (82.6) | 0.002 | 269279 (74.9) | 267655 (74.5) | 0.01 | 58852 (88.1) | 58643 (87.8) | 0.01 | 34884 (80.1) | 34657 (79.6) | 0.01 | 5267 (81.0) | 5149 (79.2) | 0.05 |
| Unknown | 14517 (15.6) | 14648 (15.8) | 0.004 | 67455 (18.8) | 68542 (19.1) | 0.008 | 4047 (6.1) | 4005 (6.0) | 0.003 | 7059 (16.2) | 7230 (16.6) | 0.01 | 721 (11.1) | 788 (12.1) | 0.03 |
| **Comorbidities** |  |  |  |  |  |  |  |  |  |  |  |  |  |  |  |
| Human immunodeficiency virus disease | 820 (0.9) | 295 (0.3) | 0.07 | 4879 (1.4) | 7328 (2.0) | 0.05 | 1778 (2.7) | 2203 (3.3) | 0.04 | 428 (1.0) | 284 (0.7) | 0.04 | 252 (3.9) | 440 (6.8) | 0.1 |
| Neoplasms | 25839 (27.8) | 26073 (28.0) | 0.006 | 127816 (35.6) | 126679 (35.2) | 0.007 | 21697 (32.5) | 21809 (32.7) | 0.004 | 7050 (16.2) | 7155 (16.4) | 0.007 | 3548 (54.6) | 3299 (50.8) | 0.08 |
| Diabetes mellitus | 8398 (9.0) | 8326 (9.0) | 0.003 | 71077 (19.8) | 69429 (19.3) | 0.01 | 8350 (12.5) | 8772 (13.1) | 0.02 | 2374 (5.5) | 2400 (5.5) | 0.003 | 3446 (53.0) | 3363 (51.7) | 0.03 |
| Overweight and obesity | 9978 (10.7) | 10104 (10.9) | 0.004 | 70644 (19.7) | 69210 (19.3) | 0.01 | 10311 (15.4) | 10647 (15.9) | 0.01 | 3528 (8.1) | 3539 (8.1) | 9E-04 | 2169 (33.4) | 2062 (31.7) | 0.04 |
| Unspecified dementia | 267 (0.3) | 319 (0.3) | 0.01 | 3670 (1.0) | 3631 (1.0) | 0.001 | 407 (0.6) | 557 (0.8) | 0.03 | 59 (0.1) | 44 (0.1) | 0.01 | 41 (0.6) | 47 (0.7) | 0.01 |
| Nicotine dependence | 8124 (8.7) | 8159 (8.8) | 0.001 | 44518 (12.4) | 43295 (12.0) | 0.01 | 10615 (15.9) | 11096 (16.6) | 0.02 | 2798 (6.4) | 2733 (6.3) | 0.006 | 993 (15.3) | 892 (13.7) | 0.04 |

#### **Supplementary Table 5 (continued)**: Baseline characteristics of COVID-19 compared to other infectious diseases, after matching.

| **Characteristics ᵃ** | COVID-19 | Lymes disease |  | COVID-19 | Herpes zoster | SMD | COVID-19 | Varicella Zoster |  | COVID-19 | Infectious mononucleosis |  | COVID-19 | Cytomegalovirus |  |
| --- | --- | --- | --- | --- | --- | --- | --- | --- | --- | --- | --- | --- | --- | --- | --- |
|  | No. (%) |  | SMD | No. (%) |  | SMD | No. (%) |  | SMD | No. (%) |  | SMD | No. (%) |  | SMD |
| Paraplegia and quadriplegia | 318 (0.3) | 114 (0.1) | 0.05 | 1490 (0.4) | 1070 (0.3) | 0.02 | 289 (0.4) | 188 (0.3) | 0.03 | 131 (0.3) | 58 (0.1) | 0.04 | 74 (1.1) | 71 (1.1) | 0.004 |
| Essential hypertension | 24996 (26.9) | 25115 (27.0) | 0.003 | 154675 (43.0) | 153395 (42.7) | 0.007 | 16094 (24.1) | 16400 (24.6) | 0.01 | 5110 (11.7) | 5101 (11.7) | 6E-04 | 5064 (77.9) | 4957 (76.3) | 0.04 |
| Ischemic heart diseases | 6892 (7.4) | 7277 (7.8) | 0.02 | 49726 (13.8) | 48462 (13.5) | 0.01 | 4475 (6.7) | 4759 (7.1) | 0.02 | 1432 (3.3) | 1486 (3.4) | 0.007 | 2562 (39.4) | 2501 (38.5) | 0.02 |
| Acute myocardial infarction | 1456 (1.6) | 1267 (1.4) | 0.02 | 11892 (3.3) | 10206 (2.8) | 0.03 | 1302 (1.9) | 1307 (2.0) | 5E-04 | 372 (0.9) | 295 (0.7) | 0.02 | 841 (12.9) | 602 (9.3) | 0.1 |
| Heart failure | 2980 (3.2) | 2152 (2.3) | 0.05 | 25339 (7.1) | 21707 (6.0) | 0.04 | 2298 (3.4) | 2104 (3.1) | 0.02 | 701 (1.6) | 753 (1.7) | 0.009 | 1651 (25.4) | 1894 (29.1) | 0.08 |
| Heart disease | 1113 (1.2) | 1299 (1.4) | 0.02 | 9915 (2.8) | 10265 (2.9) | 0.006 | 2451 (3.7) | 2766 (4.1) | 0.02 | 328 (0.8) | 380 (0.9) | 0.01 | 574 (8.8) | 587 (9.0) | 0.007 |
| Cerebral infarction | 1495 (1.6) | 1618 (1.7) | 0.01 | 14438 (4.0) | 14620 (4.1) | 0.003 | 3370 (5.0) | 3750 (5.6) | 0.03 | 432 (1.0) | 431 (1.0) | 2E-04 | 559 (8.6) | 520 (8.0) | 0.02 |
| Peripheral vascular disease | 2122 (2.3) | 1599 (1.7) | 0.04 | 16240 (4.5) | 13933 (3.9) | 0.03 | 1523 (2.3) | 1396 (2.1) | 0.01 | 426 (1.0) | 363 (0.8) | 0.02 | 809 (12.4) | 639 ( 9.8) | 0.08 |
| Chronic lower respiratory diseases | 14206 (15.3) | 14451 (15.5) | 0.007 | 80714 (22.5) | 78826 (21.9) | 0.01 | 11794 (17.7) | 11993 (18.0) | 0.008 | 6731 (15.5) | 6876 (15.8) | 0.009 | 1957 (30.1) | 1922 (29.6) | 0.01 |
| Chronic obstructive pulmonary disease | 3210 (3.5) | 2799 (3.0) | 0.02 | 24196 (6.7) | 23259 (6.5) | 0.01 | 1981 (3.0) | 1886 (2.8) | 0.008 | 602 (1.4) | 569 (1.3) | 0.007 | 781 (12.0) | 809 (12.4) | 0.01 |
| Asthma | 8157 (8.8) | 8239 (8.9) | 0.003 | 44660 (12.4) | 43600 (12.1) | 0.009 | 8333 (12.5) | 8526 (12.8) | 0.009 | 4830 (11.1) | 4904 (11.3) | 0.005 | 865 (13.3) | 836 (12.9) | 0.01 |
| Peptic ulcer | 579 (0.6) | 595 (0.6) | 0.002 | 3737 (1.0) | 4035 (1.1) | 0.008 | 514 (0.8) | 458 (0.7) | 0.01 | 181 (0.4) | 133 (0.3) | 0.02 | 189 (2.9) | 167 (2.6) | 0.02 |
| Fibrosis and cirrhosis of liver | 418 (0.5) | 528 (0.6) | 0.02 | 4893 (1.4) | 5171 (1.4) | 0.007 | 946 (1.4) | 1115 (1.7) | 0.02 | 341 (0.8) | 400 (0.9) | 0.01 | 818 (12.6) | 857 (13.2) | 0.02 |
| Portal hypertension | 232 (0.2) | 143 (0.2) | 0.02 | 2072 (0.6) | 2015 (0.6) | 0.002 | 287 (0.4) | 406 (0.6) | 0.02 | 132 (0.3) | 203 (0.5) | 0.03 | 383 (5.9) | 594 (9.1) | 0.1 |
| Psoriasis | 1264 (1.4) | 1622 (1.7) | 0.03 | 7505 (2.1) | 7680 (2.1) | 0.003 | 1092 (1.6) | 1192 (1.8) | 0.01 | 365 (0.8) | 401 (0.9) | 0.009 | 108 (1.7) | 114 (1.8) | 0.007 |
| Lupus erythematosus | 130 (0.1) | 220 (0.2) | 0.02 | 2109 (0.6) | 2196 (0.6) | 0.003 | 363 (0.5) | 462 (0.7) | 0.02 | 95 (0.2) | 100 (0.2) | 0.002 | 108 (1.7) | 107 (1.6) | 0.001 |

#### **Supplementary Table 5 (continued)**: Baseline characteristics of COVID-19 compared to other infectious diseases, after matching.

| **Characteristics ᵃ** | COVID-19 | Lymes disease |  | COVID-19 | Herpes zoster | SMD | COVID-19 | Varicella Zoster |  | COVID-19 | Infectious mononucleosis |  | COVID-19 | Cytomegalovirus |  |
| --- | --- | --- | --- | --- | --- | --- | --- | --- | --- | --- | --- | --- | --- | --- | --- |
|  | No. (%) |  | SMD | No. (%) |  | SMD | No. (%) |  | SMD | No. (%) |  | SMD | No. (%) |  | SMD |
| Rheumatoid arthritis without rheumatoid factor | 92 (0.1) | 75 (0.08) | 0.006 | 767 (0.2) | 529 (0.1) | 0.02 | 75 (0.1) | 48 (0.07) | 0.01 | 21 (0.05) | 10 (0.02) | 0.01 | 15 (0.2) | 10 (0.2) | 0.02 |
| Rheumatoid arthritis | 1053 (1.1) | 1419 (1.5) | 0.03 | 8933 (2.5) | 9774 (2.7) | 0.01 | 891 (1.3) | 1012 (1.5) | 0.02 | 306 (0.7) | 339 (0.8) | 0.009 | 190 (2.9) | 180 (2.8) | 0.009 |
| Acute kidney failure | 2745 (3.0) | 1503 (1.6) | 0.09 | 22875 (6.4) | 18475 (5.1) | 0.05 | 2462 (3.7) | 2164 (3.2) | 0.02 | 1025 (2.4) | 1283 (2.9) | 0.04 | 2335 (35.9) | 3509 (54.0) | 0.4 |
| Chronic kidney disease | 2675 (2.9) | 2725 (2.9) | 0.003 | 30265 (8.4) | 29576 (8.2) | 0.007 | 2743 (4.1) | 2939 (4.4) | 0.01 | 1477 (3.4) | 1537 (3.5) | 0.008 | 4285 (65.9) | 4200 (64.6) | 0.03 |
| End stage renal disease | 399 (0.4) | 229 (0.2) | 0.03 | 4973 (1.4) | 5620 (1.6) | 0.01 | 593 (0.9) | 698 (1.0) | 0.02 | 399 (0.9) | 812 (1.9) | 0.08 | 1382 (21.3) | 2823 (43.4) | 0.5 |

Abbreviations: SMD: standardized mean difference

ᵃ Patients were identified using *International Classification of Diseases and Related Health Problems, Tenth Revision* (*ICD-10*) and *Current Procedural Terminology (CPT)* codes. Characteristics were identified using electronic medical health record data from the TriNetX Research Network, and were recorded any time prior to the index event. Diagnostic coding can be found in the **online supplementary methods**.

#### **Supplementary Table 6**. ORs of new-onset outcomes between 3 months and one year after COVID-19, compared to contemporary negative controls and influenza patients.

|  | **Contemporary negative controls** | | | **Influenza** | | |
| --- | --- | --- | --- | --- | --- | --- |
|  | OR (95% CI) | P-value |  | OR (95% CI) | P-value |  |
| **Outcome ᵃ** |  | Uncorrected | Corrected |  | Uncorrected | Corrected |
| **Gastrointestinal symptoms or diagnoses** | 1.36 (1.33 - 1.38) | <.0001 | <.0001 | 1.01 (0.99 - 1.03) | 0,54 | 1 |
| Dysphagia | 1.56 (1.51 - 1.61) | <.0001 | <.0001 | 1.21 (1.16 - 1.26) | <.0001 | <.0001 |
| GERD | 1.30 (1.27 - 1.33) | <.0001 | <.0001 | 1.09 (1.06 - 1.12) | <.0001 | <.0001 |
| Heartburn | 1.42 (1.36 - 1.49) | <.0001 | <.0001 | 1.06 (0.99 - 1.13) | 0,08 | 0,35 |
| Nausea | 1.80 (1.75 - 1.84) | <.0001 | <.0001 | 1.19 (1.16 - 1.23) | <.0001 | <.0001 |
| Vomiting | 1.99 (1.92 - 2.06) | <.0001 | <.0001 | 1.25 (1.20 - 1.31) | <.0001 | <.0001 |
| Early satiety | 1.62 (1.46 - 1.79) | <.0001 | <.0001 | 1.62 (1.43 - 1.83) | <.0001 | <.0001 |
| Abdominal and pelvic pain | 1.46 (1.43 - 1.48) | <.0001 | <.0001 | 0.95 (0.93 - 0.97) | <.0001 | <.0001 |
| Bloating | 1.72 (1.66 - 1.78) | <.0001 | <.0001 | 1.62 (1.55 - 1.70) | <.0001 | <.0001 |
| Gastroparesis | 1.36 (1.24 - 1.49) | <.0001 | <.0001 | 1.26 (1.12 - 1.41) | <.0001 | 0,0004 |
| Functional dyspepsia | 1.56 (1.46 - 1.67) | <.0001 | <.0001 | 0.92 (0.84 - 0.99) | 0,03 | 0,15 |
| Constipation | 1.47 (1.44 - 1.51) | <.0001 | <.0001 | 1.20 (1.16 - 1.24) | <.0001 | <.0001 |
| Diarrhea | 1.74 (1.69 - 1.78) | <.0001 | <.0001 | 1.07 (1.04 - 1.10) | <.0001 | <.0001 |
| Irritable bowel syndrome | 1.05 (1.00 - 1.11) | 0,05 | 0,2 | 1.01 (0.95 - 1.07) | 0,87 | 1 |
| Inflammatory bowel disease | 1.56 (1.49 - 1.62) | <.0001 | <.0001 | 0.70 (0.68 - 0.73) | <.0001 | <.0001 |
| **Autonomic neuropathy** | 1.34 (1.31 - 1.36) | <.0001 | <.0001 | 1.13 (1.11 - 1.16) | <.0001 | <.0001 |
| Postural symptoms | 1.43 (1.40 - 1.45) | <.0001 | <.0001 | 1.15 (1.12 - 1.17) | <.0001 | <.0001 |
| Urinary dysfunction | 1.32 (1.29 - 1.35) | <.0001 | <.0001 | 1.09 (1.06 - 1.13) | <.0001 | <.0001 |
| Sexual dysfunction | 1.12 (1.07 - 1.17) | <.0001 | <.0001 | 1.04 (0.98 - 1.10) | 0,19 | 0,78 |
| Exocrine gland dysfunction | 1.58 (1.54 - 1.63) | <.0001 | <.0001 | 1.38 (1.32 - 1.43) | <.0001 | <.0001 |
| **Sensory neuropathy** | 1.35 (1.31 - 1.39) | <.0001 | <.0001 | 0.84 (0.82 - 0.87) | <.0001 | <.0001 |
| **Motor neuropathy** | 1.32 (1.28 - 1.36) | <.0001 | <.0001 | 0.94 (0.91 - 0.97) | 0,0008 | 0,004 |
| **Drugs for autonomic neuropathy** | 1.17 (1.15 - 1.20) | <.0001 | <.0001 | 1.55 (1.51 - 1.59) | <.0001 | <.0001 |
| Fludrocortisone | 1.58 (1.37 - 1.82) | <.0001 | <.0001 | 1.28 (1.08 - 1.50) | 0,003 | 0,02 |
| Beta-blockers | 1.17 (1.14 - 1.19) | <.0001 | <.0001 | 1.55 (1.51 - 1.59) | <.0001 | <.0001 |
| Pyridostigmine | 0.99 (0.79 - 1.26) | 0,95 | 1 | 2.22 (1.54 - 3.19) | <.0001 | <.0001 |
| Midodrine | 1.97 (1.81 - 2.14) | <.0001 | <.0001 | 2.38 (2.13 - 2.67) | <.0001 | <.0001 |

ᵃ Patients and outcomes were identified using *International Classification of Diseases and Related Health Problems, Tenth Revision* (*ICD-10*) and *Current Procedural Terminology (CPT)* codes. Diagnostic coding can be found in the **online supplementary methods**.

#### **Supplementary Table 7**. Rates of new-onset outcomes between 3 months and one year after COVID-19, compared to contemporary negative controls and influenza patients.

|  | **Contemporary negative controls** | | | | **Influenza** | | | |
| --- | --- | --- | --- | --- | --- | --- | --- | --- |
|  | No. Cases / No. Subjects (%) |  | P-value |  | No. Cases / No. Subjects (%) |  | P-value |  |
| **Outcome ᵃ** | COVID-19 | Negative controls | Uncorrected | Corrected | COVID-19 | Influenza | Uncorrected | Corrected |
| **Gastrointestinal symptoms or diagnoses** | 27,231 / 256,456 (10.62) | 24,332 / 302,502 (8.04) | <.0001 | <.0001 | 18,836 / 174,966 (10.77) | 19,746 / 184,493 (10.7) | 0.54 | 1 |
| Dysphagia | 8,614 / 588,637 (1.46) | 5,702 / 603,911 (.94) | <.0001 | <.0001 | 5,079 / 378,817 (1.34) | 4,287 / 384,787 (1.11) | <.0001 | <.0001 |
| GERD | 18,181 / 462,599 (3.93) | 14,664 / 480,401 (3.05) | <.0001 | <.0001 | 12,192 / 296,759 (4.11) | 11,515 / 305,216 (3.77) | <.0001 | <.0001 |
| Heartburn | 4,100 / 621,109 (.66) | 2,923 / 628,359 (.47) | <.0001 | <.0001 | 1,986 / 397,159 (.5) | 1,887 / 399,075 (.47) | 0.08 | 0.35 |
| Nausea | 15,588 / 525,402 (2.97) | 9,509 / 568,523 (1.67) | <.0001 | <.0001 | 9,498 / 349,691 (2.72) | 8,096 / 354,101 (2.29) | <.0001 | <.0001 |
| Vomiting | 9,209 / 572,507 (1.61) | 4,938 / 605,519 (.82) | <.0001 | <.0001 | 4,233 / 384,722 (1.1) | 3,357 / 381,657 (.88) | <.0001 | <.0001 |
| Early satiety | 981 / 644,193 (.15) | 607 / 645,722 (.09) | <.0001 | <.0001 | 649 / 407,819 (.16) | 403 / 408,851 (.1) | <.0001 | <.0001 |
| Abdominal and pelvic pain | 24,608 / 382,689 (6.43) | 19,342 / 428,862 (4.51) | <.0001 | <.0001 | 16,274 / 256,053 (6.36) | 17,502 / 262,692 (6.66) | <.0001 | <.0001 |
| Bloating | 8,870 / 604,175 (1.47) | 5,250 / 612,352 (.86) | <.0001 | <.0001 | 5,153 / 386,894 (1.33) | 3,206 / 388,711 (.82) | <.0001 | <.0001 |
| Gastroparesis | 1,054 / 641,927 (.16) | 778 / 643,372 (.12) | <.0001 | <.0001 | 678 / 406,270 (.17) | 540 / 406,898 (.13) | <.0001 | 0.0004 |
| Functional dyspepsia | 2,169 / 631,406 (.34) | 1,400 / 634,871 (.22) | <.0001 | <.0001 | 1,128 / 401,703 (.28) | 1,228 / 400,358 (.31) | 0.03 | 0.15 |
| Constipation | 15,399 / 528,863 (2.91) | 11,078 / 555,487 (1.99) | <.0001 | <.0001 | 9,001 / 349,786 (2.57) | 7,701 / 357,994 (2.15) | <.0001 | <.0001 |
| Diarrhea | 15,908 / 514,351 (3.09) | 10,066 / 558,169 (1.8) | <.0001 | <.0001 | 9,704 / 339,945 (2.85) | 9,206 / 344,212 (2.67) | <.0001 | <.0001 |
| Irritable bowel syndrome | 3,006 / 622,124 (.48) | 2,858 / 622,537 (.46) | 0.05 | 0.2 | 2,086 / 393,426 (.53) | 2,080 / 394,309 (.53) | 0.87 | 1 |
| Inflammatory bowel disease | 5,914 / 594,010 (1.) | 3,889 / 605,688 (.64) | <.0001 | <.0001 | 3,805 / 377,341 (1.01) | 5,330 / 373,895 (1.43) | <.0001 | <.0001 |
| **Autonomic neuropathy** | 29,503 / 346,915 (8.5) | 24,394 / 374,772 (6.51) | <.0001 | <.0001 | 20,166 / 236,722 (8.52) | 19,094 / 251,286 (7.6) | <.0001 | <.0001 |
| Postural symptoms | 25,400 / 418,279 (6.07) | 19,682 / 453,666 (4.34) | <.0001 | <.0001 | 16,559 / 284,328 (5.82) | 15,090 / 294,714 (5.12) | <.0001 | <.0001 |
| Urinary dysfunction | 14,650 / 536,156 (2.73) | 11,430 / 548,359 (2.08) | <.0001 | <.0001 | 9,452 / 343,899 (2.75) | 8,907 / 353,300 (2.52) | <.0001 | <.0001 |
| Sexual dysfunction | 4,211 / 615,922 (.68) | 3,765 / 614,667 (.61) | <.0001 | <.0001 | 2,586 / 391,382 (.66) | 2,498 / 392,299 (.64) | 0.19 | 0.78 |
| Exocrine gland dysfunction | 10,805 / 575,551 (1.88) | 7,055 / 591,333 (1.19) | <.0001 | <.0001 | 5,867 / 378,305 (1.55) | 4,326 / 382,210 (1.13) | <.0001 | <.0001 |

#### **Supplementary Table 7 (continued)**. Rates of new-onset outcomes between 3 months and one year after COVID-19, compared to contemporary negative controls and influenza patients.

|  | **Contemporary negative controls** | | | | **Influenza** | | | |
| --- | --- | --- | --- | --- | --- | --- | --- | --- |
|  | No. Cases / No. Subjects (%) |  | P-value |  | No. Cases / No. Subjects (%) |  | P-value |  |
| **Outcome ᵃ** | COVID-19 | Negative controls | Uncorrected | Corrected | COVID-19 | Influenza | Uncorrected | Corrected |
| **Sensory neuropathy** | 12,172 / 557,197 (2.18) | 9,252 / 568,570 (1.63) | <.0001 | <.0001 | 6,704 / 366,758 (1.83) | 7,864 / 364,457 (2.16) | <.0001 | <.0001 |
| **Motor neuropathy** | 9,814 / 573,809 (1.71) | 7,672 / 588,485 (1.3) | <.0001 | <.0001 | 5,772 / 371,470 (1.55) | 6,201 / 375,639 (1.65) | 0.0008 | 0.004 |
| **Drugs for autonomic neuropathy** | 18,537 / 447,128 (4.15) | 16,299 / 458,752 (3.55) | <.0001 | <.0001 | 12,701 / 284,217 (4.47) | 9,441 / 322,614 (2.93) | <.0001 | <.0001 |
| Fludrocortisone | 494 / 646,002 (.08) | 313 / 646,412 (.05) | <.0001 | <.0001 | 325 / 408,284 (.08) | 255 / 408,864 (.06) | 0.003 | 0.02 |
| Beta-blockers | 18,323 / 449,681 (4.07) | 16,182 / 461,069 (3.51) | <.0001 | <.0001 | 12,567 / 285,863 (4.4) | 9,342 / 323,708 (2.89) | <.0001 | <.0001 |
| Pyridostigmine | 139 / 648,237 (.02) | 140 / 648,282 (.02) | 0.95 | 1 | 93 / 409,742 (.02) | 42 / 410,007 (.01) | <.0001 | <.0001 |
| Midodrine | 1,639 / 642,857 (.25) | 838 / 645,077 (.13) | <.0001 | <.0001 | 999 / 406,650 (.25) | 422 / 408,637 (.1) | <.0001 | <.0001 |

ᵃ Patients and outcomes were identified using *International Classification of Diseases and Related Health Problems, Tenth Revision* (*ICD-10*) and *Current Procedural Terminology (CPT)* codes. Diagnostic coding can be found in the **online supplementary methods**.

#### **Supplementary Table 8**. ORs of new-onset outcomes between 3 months and one year after COVID-19, compared to other infectious diseases.

|  | **Lyme's disease** |  |  | **Herpes Zoster** |  |  | **Varicella Zoster** |  |  | **Infectious mononucleosis** |  |  | **Cytomegaloviral disease** |  |  |
| --- | --- | --- | --- | --- | --- | --- | --- | --- | --- | --- | --- | --- | --- | --- | --- |
|  | OR (95% CI) | P-value |  | OR (95% CI) | P-value |  | OR (95% CI) | P-value |  | OR (95% CI) | P-value |  | OR (95% CI) | P-value |  |
| **Outcome ᵃ** |  | Uncorrected | Corrected |  | Uncorrected | Corrected |  | Uncorrected | Corrected |  | Uncorrected | Corrected |  | Uncorrected | Corrected |
| **Gastrointestinal symptoms or diagnoses** | 1.40 (1.34 - 1.46) | <.0001 | <.0001 | 1.03 (1.00 - 1.05) | 0.02 | 0.1 | 1.17 (1.10 - 1.24) | <.0001 | 0.1 | 1.06 (1.00 - 1.13) | 0.07 | 0.53 | 0.86 (0.68 - 1.09) | 0.22 | 1 |
| Dysphagia | 1.41 (1.29 - 1.54) | <.0001 | <.0001 | 1.05 (1.01 - 1.09) | 0.02 | 0.13 | 0.71 (0.64 - 0.78) | <.0001 | 0.13 | 0.96 (0.82 - 1.11) | 0.55 | 1 | 0.96 (0.75 - 1.22) | 0.72 | 1 |
| GERD | 1.34 (1.27 - 1.42) | <.0001 | <.0001 | 1.04 (1.01 - 1.07) | 0.008 | 0.05 | 1.27 (1.19 - 1.37) | <.0001 | 0.05 | 1.07 (0.97 - 1.17) | 0.17 | 1 | 0.83 (0.68 - 1.00) | 0.05 | 0.47 |
| Heartburn | 1.36 (1.19 - 1.54) | <.0001 | <.0001 | 0.95 (0.90 - 1.01) | 0.1 | 0.48 | 0.42 (0.37 - 0.47) | <.0001 | 0.48 | 1.09 (0.90 - 1.31) | 0.39 | 1 | 0.70 (0.49 - 1.00) | 0.05 | 0.47 |
| Nausea | 1.71 (1.59 - 1.83) | <.0001 | <.0001 | 1.20 (1.16 - 1.24) | <.0001 | <.0001 | 0.69 (0.65 - 0.74) | <.0001 | <.0001 | 1.33 (1.21 - 1.45) | <.0001 | <.0001 | 0.98 (0.79 - 1.22) | 0.89 | 1 |
| Vomiting | 2.09 (1.91 - 2.30) | <.0001 | <.0001 | 1.22 (1.17 - 1.27) | <.0001 | <.0001 | 0.41 (0.38 - 0.44) | <.0001 | <.0001 | 1.49 (1.32 - 1.69) | <.0001 | <.0001 | 1.10 (0.85 - 1.42) | 0.46 | 1 |
| Early satiety | 1.63 (1.20 - 2.21) | 0.001 | 0.008 | 1.17 (1.03 - 1.32) | 0.01 | 0.08 | 0.63 (0.48 - 0.82) | 0.0007 | 0.08 | 1.05 (0.69 - 1.57) | 0.83 | 1 | 0.82 (0.44 - 1.52) | 0.52 | 1 |
| Abdominal and pelvic pain | 1.31 (1.25 - 1.38) | <.0001 | <.0001 | 0.99 (0.96 - 1.01) | 0.33 | 1 | 0.92 (0.87 - 0.97) | 0.003 | 1 | 1.02 (0.95 - 1.09) | 0.58 | 1 | 1.23 (1.02 - 1.48) | 0.03 | 0.32 |
| Bloating | 1.68 (1.53 - 1.84) | <.0001 | <.0001 | 1.31 (1.25 - 1.36) | <.0001 | <.0001 | 0.79 (0.72 - 0.86) | <.0001 | <.0001 | 1.21 (1.06 - 1.39) | 0.006 | 0.06 | 1.15 (0.90 - 1.47) | 0.26 | 1 |
| Gastroparesis | 1.16 (0.86 - 1.57) | 0.32 | 1 | 1.03 (0.92 - 1.16) | 0.6 | 1 | 1.10 (0.80 - 1.52) | 0.56 | 1 | 0.82 (0.55 - 1.21) | 0.31 | 1 | 0.66 (0.41 - 1.05) | 0.08 | 0.63 |
| Functional dyspepsia | 1.07 (0.91 - 1.26) | 0.43 | 1 | 0.84 (0.78 - 0.91) | <.0001 | <.0001 | 0.39 (0.33 - 0.46) | <.0001 | <.0001 | 0.74 (0.57 - 0.95) | 0.02 | 0.14 | 0.93 (0.60 - 1.45) | 0.75 | 1 |
| Constipation | 1.63 (1.52 - 1.75) | <.0001 | <.0001 | 1.15 (1.12 - 1.19) | <.0001 | <.0001 | 0.71 (0.66 - 0.76) | <.0001 | <.0001 | 1.28 (1.16 - 1.42) | <.0001 | <.0001 | 1.42 (1.15 - 1.76) | 0.001 | 0.03 |
| Diarrhea | 1.50 (1.40 - 1.60) | <.0001 | <.0001 | 1.08 (1.05 - 1.11) | <.0001 | <.0001 | 0.71 (0.67 - 0.76) | <.0001 | <.0001 | 1.05 (0.96 - 1.15) | 0.29 | 1 | 0.62 (0.51 - 0.76) | <.0001 | 0.0003 |
| Irritable bowel syndrome | 0.92 (0.81 - 1.05) | 0.21 | 0.97 | 0.85 (0.80 - 0.91) | <.0001 | <.0001 | 1.16 (1.00 - 1.35) | 0.05 | <.0001 | 0.81 (0.68 - 0.97) | 0.02 | 0.16 | 1.09 (0.65 - 1.81) | 0.74 | 1 |
| Inflammatory bowel disease | 1.33 (1.19 - 1.47) | <.0001 | <.0001 | 0.88 (0.84 - 0.92) | <.0001 | <.0001 | 0.95 (0.85 - 1.07) | 0.4 | <.0001 | 0.75 (0.65 - 0.86) | <.0001 | 0.0008 | 0.53 (0.40 - 0.70) | <.0001 | 0.0003 |

#### **Supplementary Table 8 (continued)**. ORs of new-onset outcomes between 3 months and one year after COVID-19, compared to other infectious diseases.

|  | **Lyme's disease** |  |  | **Herpes Zoster** |  |  | **Varicella Zoster** |  |  | **Infectious mononucleosis** |  |  | **Cytomegaloviral disease** |  |  |
| --- | --- | --- | --- | --- | --- | --- | --- | --- | --- | --- | --- | --- | --- | --- | --- |
|  | OR (95% CI) | P-value |  | OR (95% CI) | P-value |  | OR (95% CI) | P-value |  | OR (95% CI) | P-value |  | OR (95% CI) | P-value |  |
| **Outcome ᵃ** |  | Uncorrected | Corrected |  | Uncorrected | Corrected |  | Uncorrected | Corrected |  | Uncorrected | Corrected |  | Uncorrected | Corrected |
| **Autonomic neuropathy** | 1.21 (1.16 - 1.27) | <.0001 | <.0001 | 1.05 (1.03 - 1.08) | <.0001 | <.0001 | 0.95 (0.89 - 1.00) | 0.04 | <.0001 | 1.11 (1.03 - 1.18) | 0.003 | 0.04 | 1.23 (1.03 - 1.47) | 0.02 | 0.3 |
| Postural symptoms | 1.24 (1.19 - 1.31) | <.0001 | <.0001 | 1.12 (1.09 - 1.15) | <.0001 | <.0001 | 0.83 (0.79 - 0.88) | <.0001 | <.0001 | 1.21 (1.12 - 1.30) | <.0001 | <.0001 | 1.29 (1.07 - 1.56) | 0.006 | 0.11 |
| Urinary dysfunction | 1.17 (1.10 - 1.25) | <.0001 | <.0001 | 1.02 (0.99 - 1.05) | 0.28 | 1 | 0.89 (0.83 - 0.96) | 0.003 | 1 | 0.96 (0.87 - 1.07) | 0.47 | 1 | 1.20 (0.97 - 1.47) | 0.09 | 0.65 |
| Sexual dysfunction | 1.18 (1.05 - 1.32) | 0.005 | 0.03 | 0.99 (0.94 - 1.05) | 0.72 | 1 | 0.92 (0.78 - 1.07) | 0.29 | 1 | 0.93 (0.73 - 1.18) | 0.54 | 1 | 0.97 (0.72 - 1.32) | 0.87 | 1 |
| Exocrine gland dysfunction | 1.37 (1.26 - 1.48) | <.0001 | <.0001 | 0.99 (0.96 - 1.03) | 0.6 | 1 | 0.53 (0.49 - 0.57) | <.0001 | 1 | 1.02 (0.90 - 1.16) | 0.73 | 1 | 1.40 (1.11 - 1.76) | 0.005 | 0.1 |
| **Sensory neuropathy** | 1.06 (0.99 - 1.14) | 0.08 | 0.39 | 0.92 (0.89 - 0.95) | <.0001 | <.0001 | 0.49 (0.46 - 0.53) | <.0001 | <.0001 | 1.00 (0.89 - 1.12) | 0.97 | 1 | 1.37 (1.08 - 1.74) | 0.01 | 0.14 |
| **Motor neuropathy** | 0.99 (0.91 - 1.07) | 0.73 | 1 | 0.88 (0.84 - 0.91) | <.0001 | <.0001 | 0.60 (0.55 - 0.65) | <.0001 | <.0001 | 1.00 (0.87 - 1.14) | 0.96 | 1 | 1.30 (1.01 - 1.67) | 0.04 | 0.45 |
| **Drugs for autonomic neuropathy** | 1.55 (1.46 - 1.64) | <.0001 | <.0001 | 1.29 (1.25 - 1.33) | <.0001 | <.0001 | 1.68 (1.56 - 1.81) | <.0001 | <.0001 | 1.52 (1.37 - 1.68) | <.0001 | <.0001 | 0.87 (0.68 - 1.12) | 0.28 | 1 |
| Fludrocortisone | 0.93 (0.67 - 1.30) | 0.67 | 1 | 1.17 (0.99 - 1.38) | 0.06 | 0.32 | 0.74 (0.49 - 1.10) | 0.13 | 0.32 | 0.43 (0.27 - 0.70) | 0.0005 | 0.006 | 0.38 (0.22 - 0.67) | 0.0004 | 0.01 |
| Beta-blockers | 1.58 (1.49 - 1.68) | <.0001 | <.0001 | 1.28 (1.25 - 1.32) | <.0001 | <.0001 | 1.69 (1.56 - 1.82) | <.0001 | <.0001 | 1.58 (1.42 - 1.75) | <.0001 | <.0001 | 0.86 (0.67 - 1.09) | 0.21 | 1 |
| Pyridostigmine | 0.59 (0.32 - 1.10) | 0.09 | 0.44 | 1.19 (0.87 - 1.62) | 0.27 | 1 | 1.23 (0.59 - 2.56) | 0.58 | 1 | 1.00 (0.42 - 2.40) | 1 | 1 | 1.00 (0.42 - 2.41) | 1 | 1 |
| Midodrine | 1.90 (1.48 - 2.44) | <.0001 | <.0001 | 2.24 (2.01 - 2.51) | <.0001 | <.0001 | 1.71 (1.26 - 2.33) | 0.0006 | <.0001 | 1.70 (1.12 - 2.56) | 0.01 | 0.11 | 1.29 (0.87 - 1.90) | 0.21 | 1 |

Abbreviations: OR: odds ratio

ᵃ Patients and outcomes were identified using *International Classification of Diseases and Related Health Problems, Tenth Revision* (*ICD-10*) and *Current Procedural Terminology (CPT)* codes. Diagnostic coding can be found in the **online supplementary methods**.

#### **Supplementary Table 9**. Rates of new-onset outcomes between 3 months and one year after COVID-19, compared to other infectious diseases.

|  | **Lyme's disease** |  | **Herpes Zoster** |  | **Varicella Zoster** |  | **Infectious mononucleosis** |  | **Cytomegaloviral disease** |  |
| --- | --- | --- | --- | --- | --- | --- | --- | --- | --- | --- |
|  | No. Cases / No. Subjects (%) |  | No. Cases / No. Subjects (%) |  | No. Cases / No. Subjects (%) |  | No. Cases / No. Subjects (%) |  | No. Cases / No. Subjects (%) |  |
| **Outcomeᵃ** | COVID-19 | Lyme's disease | COVID-19 | Herpes Zoster | COVID-19 | Varicella Zoster | COVID-19 | Infectious mononucleosis | COVID-19 | Cytomegaloviral disease |
| **Gastrointestinal symptoms or diagnoses** | 4,553 / 45,148 (10.08) | 3,841 / 51,679 (7.43) | 15,565 / 144,347 (10.78) | 16,596 / 157,809 (10.52) | 2,869 / 28,118 (10.2) | 2,225 / 25,057 (8.88) | 2,109 / 23,152 (9.11) | 2,028 / 23,492 (8.63) | 189 / 1,454 (13.) | 144 / 977 (14.74) |
| Dysphagia | 1,213 / 86,154 (1.41) | 884 / 88,278 (1.) | 5,295 / 322,979 (1.64) | 5,182 / 330,190 (1.57) | 734 / 61,203 (1.2) | 1,020 / 60,545 (1.68) | 345 / 41,355 (.83) | 359 / 41,167 (.87) | 135 / 5,325 (2.54) | 136 / 5,138 (2.65) |
| GERD | 2,685 / 71,464 (3.76) | 2,098 / 74,332 (2.82) | 10,719 / 250,898 (4.27) | 10,611 / 257,375 (4.12) | 1,750 / 51,831 (3.38) | 1,400 / 52,480 (2.67) | 960 / 37,107 (2.59) | 909 / 37,418 (2.43) | 212 / 3,591 (5.9) | 222 / 3,144 (7.06) |
| Heartburn | 541 / 89,950 (.6) | 403 / 90,805 (.44) | 2,265 / 343,175 (.66) | 2,390 / 344,884 (.69) | 386 / 63,512 (.61) | 899 / 62,260 (1.44) | 228 / 42,297 (.54) | 212 / 42,665 (.5) | 52 / 6,017 (.86) | 74 / 6,020 (1.23) |
| Nausea | 2,196 / 80,096 (2.74) | 1,373 / 84,526 (1.62) | 8,801 / 295,983 (2.97) | 7,674 / 308,228 (2.49) | 1,603 / 52,618 (3.05) | 2,111 / 48,498 (4.35) | 1,115 / 36,828 (3.03) | 879 / 38,246 (2.3) | 166 / 4,550 (3.65) | 168 / 4,536 (3.7) |
| Vomiting | 1,334 / 85,190 (1.57) | 668 / 88,612 (.75) | 5,357 / 320,937 (1.67) | 4,527 / 329,040 (1.38) | 932 / 56,905 (1.64) | 2,001 / 51,358 (3.9) | 599 / 38,852 (1.54) | 420 / 40,474 (1.04) | 123 / 5,123 (2.4) | 119 / 5,440 (2.19) |
| Early satiety | 109 / 92,529 (.12) | 67 / 92,748 (.07) | 545 / 356,614 (.15) | 467 / 357,351 (.13) | 84 / 66,322 (.13) | 134 / 66,215 (.2) | 47 / 43,368 (.11) | 45 / 43,399 (.1) | 18 / 6,363 (.28) | 22 / 6,344 (.35) |
| Abdominal and pelvic pain | 3,670 / 63,285 (5.8) | 3,043 / 67,865 (4.48) | 13,798 / 221,791 (6.22) | 14,613 / 232,272 (6.29) | 2,540 / 39,337 (6.46) | 2,593 / 37,069 (7.) | 1,763 / 29,307 (6.02) | 1,769 / 29,949 (5.91) | 254 / 2,961 (8.58) | 235 / 3,318 (7.08) |
| Bloating | 1,196 / 88,038 (1.36) | 728 / 89,414 (.81) | 5,132 / 333,847 (1.54) | 3,983 / 337,120 (1.18) | 873 / 62,210 (1.4) | 1,066 / 60,043 (1.78) | 457 / 41,542 (1.1) | 382 / 41,951 (.91) | 140 / 5,554 (2.52) | 120 / 5,467 (2.19) |
| Gastroparesis | 92 / 92,465 (.1) | 79 / 92,438 (.09) | 547 / 355,949 (.15) | 530 / 356,157 (.15) | 78 / 66,218 (.12) | 71 / 66,305 (.11) | 45 / 43,291 (.1) | 55 / 43,242 (.13) | 30 / 6,190 (.48) | 44 / 6,011 (.73) |
| Functional dyspepsia | 298 / 91,275 (.33) | 278 / 90,936 (.31) | 1,237 / 350,035 (.35) | 1,459 / 348,347 (.42) | 199 / 64,978 (.31) | 499 / 63,916 (.78) | 108 / 42,879 (.25) | 146 / 42,768 (.34) | 38 / 6,144 (.62) | 41 / 6,169 (.66) |
| Constipation | 2,109 / 81,192 (2.6) | 1,370 / 85,128 (1.61) | 9,081 / 295,751 (3.07) | 8,165 / 305,029 (2.68) | 1,414 / 54,748 (2.58) | 1,877 / 51,820 (3.62) | 832 / 37,974 (2.19) | 679 / 39,482 (1.72) | 216 / 4,363 (4.95) | 154 / 4,361 (3.53) |
| Diarrhea | 2,308 / 78,418 (2.94) | 1,643 / 82,786 (1.98) | 9,016 / 288,080 (3.13) | 8,719 / 299,594 (2.91) | 1,554 / 52,541 (2.96) | 1,992 / 48,715 (4.09) | 982 / 36,682 (2.68) | 969 / 37,962 (2.55) | 183 / 4,082 (4.48) | 228 / 3,249 (7.02) |
| Irritable bowel syndrome | 438 / 89,551 (.49) | 471 / 88,663 (.53) | 1,640 / 343,913 (.48) | 1,922 / 343,167 (.56) | 360 / 63,912 (.56) | 309 / 63,666 (.49) | 217 / 42,197 (.51) | 266 / 41,881 (.64) | 31 / 6,189 (.5) | 29 / 6,296 (.46) |
| Inflammatory bowel disease | 782 / 87,070 (.9) | 598 / 88,058 (.68) | 3,261 / 331,094 (.98) | 3,710 / 332,530 (1.12) | 571 / 61,395 (.93) | 595 / 60,911 (.98) | 338 / 40,573 (.83) | 449 / 40,411 (1.11) | 79 / 5,548 (1.42) | 141 / 5,333 (2.64) |

#### **Supplementary Table 9 (continued)**. Rates of new-onset outcomes between 3 months and one year after COVID-19, compared to other infectious diseases.

|  | **Lyme's disease** |  | **Herpes Zoster** |  | **Varicella Zoster** |  | **Infectious mononucleosis** |  | **Cytomegaloviral disease** |  |
| --- | --- | --- | --- | --- | --- | --- | --- | --- | --- | --- |
|  | No. Cases / No. Subjects (%) |  | No. Cases / No. Subjects (%) |  | No. Cases / No. Subjects (%) |  | No. Cases / No. Subjects (%) |  | No. Cases / No. Subjects (%) |  |
| **Outcomeᵃ** | COVID-19 | Lyme's disease | COVID-19 | Herpes Zoster | COVID-19 | Varicella Zoster | COVID-19 | Infectious mononucleosis | COVID-19 | Cytomegaloviral disease |
| **Autonomic neuropathy** | 4,570 / 56,513 (8.09) | 3,949 / 58,287 (6.78) | 17,425 / 188,751 (9.23) | 17,960 / 203,831 (8.81) | 2,921 / 37,943 (7.7) | 2,667 / 32,893 (8.11) | 1,869 / 29,944 (6.24) | 1,762 / 31,023 (5.68) | 270 / 2,266 (11.92) | 268 / 2,706 (9.9) |
| Postural symptoms | 3,839 / 66,467 (5.78) | 3,201 / 68,175 (4.7) | 15,040 / 230,401 (6.53) | 14,452 / 246,341 (5.87) | 2,433 / 44,422 (5.48) | 2,558 / 39,328 (6.5) | 1,573 / 33,025 (4.76) | 1,359 / 34,167 (3.98) | 250 / 3,159 (7.91) | 231 / 3,707 (6.23) |
| Urinary dysfunction | 2,016 / 80,165 (2.51) | 1,758 / 81,701 (2.15) | 8,780 / 294,020 (2.99) | 8,902 / 302,896 (2.94) | 1,379 / 56,667 (2.43) | 1,522 / 56,113 (2.71) | 716 / 39,361 (1.82) | 756 / 40,052 (1.89) | 205 / 4,731 (4.33) | 180 / 4,931 (3.65) |
| Sexual dysfunction | 655 / 88,625 (.74) | 557 / 88,465 (.63) | 2,442 / 339,750 (.72) | 2,471 / 340,324 (.73) | 301 / 64,594 (.47) | 326 / 64,253 (.51) | 125 / 42,823 (.29) | 135 / 42,860 (.31) | 83 / 5,747 (1.44) | 90 / 6,074 (1.48) |
| Exocrine gland dysfunction | 1,487 / 85,172 (1.75) | 1,111 / 86,466 (1.28) | 6,569 / 316,974 (2.07) | 6,677 / 319,306 (2.09) | 987 / 59,043 (1.67) | 1,723 / 55,200 (3.12) | 476 / 40,796 (1.17) | 469 / 41,111 (1.14) | 167 / 5,242 (3.19) | 127 / 5,518 (2.3) |
| **Sensory neuropathy** | 1,666 / 83,096 (2.) | 1,544 / 81,898 (1.89) | 7,085 / 309,266 (2.29) | 7,613 / 307,481 (2.48) | 1,166 / 57,823 (2.02) | 2,182 / 54,077 (4.03) | 584 / 40,226 (1.45) | 591 / 40,628 (1.45) | 154 / 5,131 (3.) | 125 / 5,659 (2.21) |
| **Motor neuropathy** | 1,292 / 85,507 (1.51) | 1,314 / 85,785 (1.53) | 5,579 / 318,959 (1.75) | 6,432 / 322,834 (1.99) | 922 / 59,689 (1.54) | 1,476 / 57,992 (2.55) | 449 / 40,964 (1.1) | 455 / 41,384 (1.1) | 137 / 5,329 (2.57) | 112 / 5,617 (1.99) |
| **Drugs for autonomic neuropathy** | 2,690 / 69,348 (3.88) | 1,944 / 76,441 (2.54) | 11,261 / 237,108 (4.75) | 9,992 / 268,397 (3.72) | 1,710 / 51,838 (3.3) | 1,116 / 56,126 (1.99) | 912 / 37,473 (2.43) | 632 / 39,068 (1.62) | 155 / 2,287 (6.78) | 117 / 1,521 (7.69) |
| Fludrocortisone | 67 / 92,681 (.07) | 72 / 92,598 (.08) | 302 / 357,625 (.08) | 258 / 357,983 (.07) | 42 / 66,468 (.06) | 57 / 66,517 (.09) | 24 / 43,374 (.06) | 55 / 43,237 (.13) | 18 / 6,330 (.28) | 44 / 5,983 (.74) |
| Beta-blockers | 2,664 / 69,698 (3.82) | 1,883 / 76,803 (2.45) | 11,100 / 238,598 (4.65) | 9,870 / 269,550 (3.66) | 1,679 / 52,138 (3.22) | 1,091 / 56,365 (1.94) | 904 / 37,638 (2.4) | 602 / 39,263 (1.53) | 156 / 2,343 (6.66) | 127 / 1,651 (7.69) |
| Pyridostigmine | 16 / 92,853 (.02) | 27 / 92,793 (.03) | 88 / 358,710 (.02) | 74 / 358,822 (.02) | 16 / 66,679 (.02) | 13 / 66,707 (.02) | 10 / 43,503 (.02) | 10 / 43,505 (.02) | 10 / 6,472 (.15) | 10 / 6,490 (.15) |
| Midodrine | 176 / 92,400 (.19) | 93 / 92,673 (.1) | 992 / 355,933 (.28) | 445 / 357,633 (.12) | 109 / 66,278 (.16) | 64 / 66,475 (.1) | 61 / 43,301 (.14) | 36 / 43,297 (.08) | 60 / 6,138 (.98) | 44 / 5,774 (.76) |

ᵃ Patients and outcomes were identified using *International Classification of Diseases and Related Health Problems, Tenth Revision* (*ICD-10*) and *Current Procedural Terminology (CPT)* codes. Diagnostic coding can be found in the **online supplementary methods**.

#### **Supplementary table 10.** Baseline characteristics of influenza compared to contemporary negative controls, before and after matching.

|  | **Before matching** | | | **After matching** | | |
| --- | --- | --- | --- | --- | --- | --- |
| **Characteristics ^a^** | Influenza | Negative controls | SMD | Influenza | Negative controls | SMD |
| **Number** | 500854 | 2505344 | - | 500852 | 500852 | - |
| **Age; mean (SD); y** | 45.5 (17.1) | 54.7 (17.5) | 0.5 | 45.5 (17.1) | 45.5 (17.2) | 1 |
| **Sex** |  |  |  |  |  |  |
| Female | 300433 (60.0) | 1470931 (58.7) | 0.03 | 300433 (60.0) | 297379 (59.4) | 0.01 |
| Male | 184307 (36.8) | 928323 (37.1) | 5 | 184305 (36.8) | 187044 (37.3) | 0.01 |
| **Race** |  |  |  |  |  |  |
| White | 324561 (64.8) | 1660944 (66.3) | 0.03 | 324561 (64.8) | 326870 (65.3) | 0.01 |
| Black or African American | 67901 (13.6) | 268029 (10.7) | 0.09 | 67900 (13.6) | 65448 (13.1) | 0.01 |
| **Ethnicity** |  |  |  |  |  |  |
| Hispanic or Latino | 46285 (9.2) | 149589 (6.0) | 0.1 | 46284 (9.2) | 45416 (9.1) | 6 |
| Not Hispanic of Latino | 355491 (71.0) | 1723238 (68.8) | 0.05 | 355490 (71.0) | 357646 (71.4) | 0.01 |
| Unknown | 99078 (19.8) | 632517 (25.2) | 0.1 | 99078 (19.8) | 97790 (19.5) | 6 |
| **Comorbidities** |  |  |  |  |  |  |
| Human immunodeficiency virus [HIV] disease | 4829 (1.0) | 19307 (0.8) | 0.02 | 4829 (1.0) | 4033 (0.8) | 0.02 |
| Neoplasms | 122345 (24.4) | 907417 (36.2) | 0.3 | 122345 (24.4) | 120010 (24.0) | 0.01 |
| Diabetes mellitus | 70449 (14.1) | 480845 (19.2) | 0.1 | 70449 (14.1) | 67157 (13.4) | 0.02 |
| Overweight and obesity | 92896 (18.5) | 572853 (22.9) | 0.1 | 92896 (18.5) | 91667 (18.3) | 6 |
| Unspecified dementia | 2829 (0.6) | 23544 (0.9) | 0.04 | 2829 (0.6) | 2588 (0.5) | 7 |
| Nicotine dependence | 63188 (12.6) | 313163 (12.5) | 4 | 63188 (12.6) | 62168 (12.4) | 6 |
| Paraplegia (paraparesis) and quadriplegia (quadriparesis) | 1473 (0.3) | 10978 (0.4) | 0.02 | 1473 (0.3) | 1400 (0.3) | 3 |
| Essential (primary) hypertension | 153316 (30.6) | 1053055 (42.0) | 0.2 | 153315 (30.6) | 150853 (30.1) | 0.01 |
| Ischemic heart diseases | 44553 (8.9) | 346742 (13.8) | 0.2 | 44553 (8.9) | 41507 (8.3) | 0.02 |
| Acute myocardial infarction | 10381 (2.1) | 75260 (3.0) | 0.06 | 10381 (2.1) | 9319 (1.9) | 0.02 |
| Heart failure | 23340 (4.7) | 166045 (6.6) | 0.09 | 23340 (4.7) | 20213 (4.0) | 0.03 |
| Heart disease; unspecified | 7720 (1.5) | 55309 (2.2) | 0.05 | 7720 (1.5) | 6716 (1.3) | 0.02 |
| Cerebral infarction | 9712 (1.9) | 82529 (3.3) | 0.08 | 9712 (1.9) | 8628 (1.7) | 0.02 |
| Peripheral vascular disease; unspecified | 11242 (2.2) | 95868 (3.8) | 0.09 | 11242 (2.2) | 10122 (2.0) | 0.02 |
| Chronic obstructive pulmonary disease; unspecified | 26406 (5.3) | 151699 (6.1) | 0.03 | 26404 (5.3) | 23977 (4.8) | 0.02 |
| Asthma | 77742 (15.5) | 317747 (12.7) | 0.08 | 77740 (15.5) | 74247 (14.8) | 0.02 |
| Peptic ulcer; site unspecified | 4693 (0.9) | 29703 (1.2) | 0.02 | 4693 (0.9) | 3996 (0.8) | 0.02 |
| Fibrosis and cirrhosis of liver | 4613 (0.9) | 40943 (1.6) | 0.06 | 4613 (0.9) | 3855 (0.8) | 0.02 |
| Portal hypertension | 1584 (0.3) | 16110 (0.6) | 0.05 | 1584 (0.3) | 1366 (0.3) | 8 |
| Psoriasis | 7653 (1.5) | 54361 (2.2) | 0.05 | 7653 (1.5) | 7291 (1.5) | 6 |
| Lupus erythematosus | 1434 (0.3) | 9139 (0.4) | 0.01 | 1434 (0.3) | 1267 (0.3) | 6 |
| Rheumatoid arthritis without rheumatoid factor; multiple sites | 427 (0.09) | 5487 (0.2) | 0.03 | 427 (0.09) | 556 (0.1) | 8 |
| Rheumatoid arthritis; unspecified | 7734 (1.5) | 55554 (2.2) | 0.05 | 7734 (1.5) | 6961 (1.4) | 0.01 |
| Acute kidney failure | 21298 (4.3) | 151487 (6.0) | 0.08 | 21298 (4.3) | 18547 (3.7) | 0.03 |
| Chronic kidney disease (CKD) | 27084 (5.4) | 228968 (9.1) | 0.1 | 27084 (5.4) | 24263 (4.8) | 0.03 |
| End stage renal disease | 6349 (1.3) | 39594 (1.6) | 0.03 | 6349 (1.3) | 5164 (1.0) | 0.02 |

Abbreviations: SMD: standardized mean difference

ᵃ Patients were identified using *International Classification of Diseases and Related Health Problems, Tenth Revision* (*ICD-10*) and *Current Procedural Terminology (CPT)* codes. Characteristics were identified using electronic medical health record data from the TriNetX Research Network, and were recorded any time prior to the index event. Diagnostic coding can be found in the **online supplementary methods**.

#### **Supplementary table 11.** Rates of new-onset inflammatory bowel disease (IBD) between 3 months and one year after influenza, compared to contemporary negative controls, after matching.

| **Outcome** | **OR (95% CI)** | **Rate (%)** | | | **No. Cases / No. Subjects** | | **P-value** | |
| --- | --- | --- | --- | --- | --- | --- | --- | --- |
| **Outcome^a^** |  | **Influenza** | **Contemporary negative controls** | **Excess** | **Influenza** | **Contemporary negative controls** | **Uncorrected** | **Corrected** |
| **Inflammatory bowel disease** | 1.87 (1.79 - 1.96) | 1.21 | 0.65 | 0.56 | 5,582 / 459,813 | 3,060 / 469,837 | <.0001 | <.0001 |

ᵃ Patients and outcomes were identified using *International Classification of Diseases and Related Health Problems, Tenth Revision* (*ICD-10*) and *Current Procedural Terminology (CPT)* codes. Diagnostic coding can be found in the **online supplementary methods**.

#### **Supplementary Table 12**: Baseline characteristics for COVID-19 patients with and without anosmia or prior vaccination, and influenza patients with and without prior vaccination, before matching.

|  | **Before matching for baseline demographics and risk factors** | | | | | | | | |
| --- | --- | --- | --- | --- | --- | --- | --- | --- | --- |
| **Characteristics ᵃ** | COVID-19 with anosmia | COVID-19 without anosmia |  | COVID-19 after prior vaccination | COVID-19 without prior vaccination |  | Influenza after prior vaccination | Influenza without prior vaccination |  |
|  | No. (%) |  | SMD | No. (%) |  | SMD | No. (%) |  | SMD |
| **Number** | 12207 | 637271 | - | 18101 | 625373 | - | 89940 | 347965 | - |
| **Age; mean (SD); y** | 44.7 (16.2) | 50.5 (17.6) | 0.3 | 52.2 (17.7) | 49.1 (17.6) | 0.2 | 50.4 (17.8) | 44.2 (16.8) | 0.4 |
| **Sex** |  |  |  |  |  |  |  |  |  |
| Female | 8055 (66.0) | 391353 (61.4) | 0.1 | 11560 (63.9) | 383713 (61.4) | 0.05 | 58493 (65.0) | 215152 (61.8) | 0.07 |
| Male | 4146 (34.0) | 245830 (38.6) | 0.1 | 6539 (36.1) | 241568 (38.6) | 0.05 | 31442 (35.0) | 132725 (38.1) | 0.07 |
| **Race** |  |  |  |  |  |  |  |  |  |
| White | 8010 (65.6) | 442323 (69.4) | 0.08 | 13188 (72.9) | 433665 (69.3) | 0.08 | 67450 (75.0) | 224156 (64.4) | 0.2 |
| Black or African American | 2389 (19.6) | 114890 (18.0) | 0.04 | 3246 (17.9) | 112290 (18.0) | 6E-04 | 11814 (13.1) | 52602 (15.1) | 0.06 |
| **Ethnicity** |  |  |  |  |  |  |  |  |  |
| Hispanic or Latino | 1320 (10.8) | 58273 (9.1) | 0.06 | 2558 (14.1) | 56360 (9.0) | 0.2 | 7785 (8.7) | 29966 (8.6) | 0.002 |
| Not Hispanic of Latino | 8483 (69.5) | 450476 (70.7) | 0.03 | 14961 (82.7) | 438975 (70.2) | 0.3 | 73010 (81.2) | 235223 (67.6) | 0.3 |
| Unknown | 2404 (19.7) | 128522 (20.2) | 0.01 | 582 (3.2) | 130038 (20.8) | 0.6 | 9145 (10.2) | 82776 (23.8) | 0.4 |
| **Comorbidities** |  |  |  |  |  |  |  |  |  |
| Human immunodeficiency virus disease | 320 (2.6) | 14359 (2.3) | 0.02 | 845 (4.7) | 13199 (2.1) | 0.1 | 1685 (1.9) | 2750 (0.8) | 0.09 |
| Neoplasms | 4475 (36.7) | 238160 (37.4) | 0.01 | 9761 (53.9) | 201893 (32.3) | 0.4 | 38018 (42.3) | 67927 (19.5) | 0.5 |
| Diabetes mellitus | 2422 (19.8) | 151050 (23.7) | 0.09 | 6097 (33.7) | 127187 (20.3) | 0.3 | 21707 (24.1) | 39913 (11.5) | 0.3 |
| Overweight and obesity | 4149 (34.0) | 201112 (31.6) | 0.05 | 7550 (41.7) | 161471 (25.8) | 0.3 | 29762 (33.1) | 51306 (14.7) | 0.4 |
| Unspecified dementia | 97 (0.8) | 11268 (1.8) | 0.09 | 490 (2.7) | 7298 (1.2) | 0.1 | 951 (1.1) | 1438 (0.4) | 0.08 |
| Nicotine dependence | 2460 (20.2) | 119148 (18.7) | 0.04 | 4700 (26.0) | 102280 (16.4) | 0.2 | 16234 (18.1) | 39178 (11.3) | 0.2 |
| Paraplegia and quadriplegia | 54 (0.4) | 3857 (0.6) | 0.02 | 159 (0.9) | 3002 (0.5) | 0.05 | 489 (0.5) | 823 (0.2) | 0.05 |

#### **Supplementary Table 12 (continued)**: Baseline characteristics for COVID-19 patients with and without anosmia or prior vaccination, and influenza patients with and without prior vaccination, before matching.

| **Characteristics ᵃ** | COVID-19 with anosmia | COVID-19 without anosmia |  | COVID-19 after prior vaccination | COVID-19 without prior vaccination |  | Influenza after prior vaccination | Influenza without prior vaccination |  |
| --- | --- | --- | --- | --- | --- | --- | --- | --- | --- |
|  | No. (%) |  | SMD | No. (%) |  | SMD | No. (%) |  | SMD |
| Essential hypertension | 4663 (38.2) | 290232 (45.5) | 0.1 | 10024 (55.4) | 244915 (39.2) | 0.3 | 44471 (49.4) | 90836 (26.1) | 0.5 |
| Ischemic heart diseases | 1369 (11.2) | 108778 (17.1) | 0.2 | 4946 (27.3) | 83439 (13.3) | 0.4 | 14418 (16.0) | 24645 (7.1) | 0.3 |
| Acute myocardial infarction | 431 (3.5) | 32824 (5.2) | 0.08 | 1906 (10.5) | 22755 (3.6) | 0.3 | 3349 (3.7) | 5384 (1.5) | 0.1 |
| Heart failure | 655 (5.4) | 61859 ( 9.7) | 0.2 | 3218 (17.8) | 43666 (7.0) | 0.3 | 7887 (8.8) | 12908 (3.7) | 0.2 |
| Heart disease | 420 (3.4) | 25949 (4.1) | 0.03 | 1818 (10.0) | 21522 (3.4) | 0.3 | 3266 (3.6) | 3567 (1.0) | 0.2 |
| Cerebral infarction | 628 (5.1) | 43422 (6.8) | 0.07 | 2576 (14.2) | 34064 (5.4) | 0.3 | 3170 (3.5) | 5135 (1.5) | 0.1 |
| Peripheral vascular disease | 394 (3.2) | 34256 (5.4) | 0.1 | 1876 (10.4) | 26231 (4.2) | 0.2 | 4193 (4.7) | 5914 (1.7) | 0.2 |
| Chronic lower respiratory diseases | 3518 (28.8) | 177681 (27.9) | 0.02 | 6782 (37.5) | 147628 (23.6) | 0.3 | 37750 (42.0) | 71967 (20.7) | 0.5 |
| Chronic obstructive pulmonary disease | 592 (4.8) | 50481 (7.9) | 0.1 | 2249 (12.4) | 38849 (6.2) | 0.2 | 9612 (10.7) | 14882 (4.3) | 0.2 |
| Asthma | 2367 (19.4) | 109639 (17.2) | 0.06 | 4283 (23.7) | 91479 (14.6) | 0.2 | 23356 (26.0) | 43749 (12.6) | 0.3 |
| Peptic ulcer | 142 (1.2) | 8468 (1.3) | 0.01 | 612 (3.4) | 6596 (1.1) | 0.2 | 1514 (1.7) | 2580 (0.7) | 0.09 |
| Fibrosis and cirrhosis of liver | 187 (1.5) | 15387 (2.4) | 0.06 | 815 (4.5) | 12097 (1.9) | 0.1 | 1428 (1.6) | 2609 (0.8) | 0.08 |
| Portal hypertension | 52 (0.4) | 6431 (1.0) | 0.07 | 351 (1.9) | 4667 (0.7) | 0.1 | 535 (0.6) | 865 (0.2) | 0.05 |
| Psoriasis | 282 (2.3) | 14892 (2.3) | 0.002 | 645 (3.6) | 12273 (2.0) | 0.1 | 2504 (2.8) | 4105 (1.2) | 0.1 |
| Lupus erythematosus | 89 (0.7) | 4137 (0.6) | 0.01 | 252 (1.4) | 3591 (0.6) | 0.08 | 489 (0.5) | 807 (0.2) | 0.05 |
| Rheumatoid arthritis without rheumatoid factor | 24 (0.2) | 1294 (0.2) | 0.001 | 48 (0.3) | 1016 (0.2) | 0.02 | 164 (0.2) | 224 (0.06) | 0.03 |
| Rheumatoid arthritis | 225 (1.8) | 14757 (2.3) | 0.03 | 700 (3.9) | 12154 (1.9) | 0.1 | 2695 (3.0) | 4182 (1.2) | 0.1 |
| Acute kidney failure | 806 (6.6) | 67106 (10.5) | 0.1 | 3284 (18.1) | 41956 (6.7) | 0.4 | 7339 (8.2) | 11597 (3.3) | 0.2 |
| Chronic kidney disease | 880 (7.2) | 72241 (11.3) | 0.1 | 3329 (18.4) | 52398 (8.4) | 0.3 | 9129 (10.2) | 14692 (4.2) | 0.2 |
| End stage renal disease | 162 (1.3) | 16236 (2.5) | 0.09 | 724 (4.0) | 10799 (1.7) | 0.1 | 1623 (1.8) | 3926 (1.1) | 0.06 |

Abbreviations: SMD: standardized mean difference

ᵃ Patients were identified using *International Classification of Diseases and Related Health Problems, Tenth Revision* (*ICD-10*) and *Current Procedural Terminology (CPT)* codes. Characteristics were identified using electronic medical health record data from the TriNetX Research Network, and were recorded any time prior to the index event. Diagnostic coding can be found in the **online supplementary methods**.

#### **Supplementary Table 13**: Baseline characteristics for COVID-19 patients with anosmia or prior vaccination compared to without anosmia or prior vaccination, after matching.

|  | **After matching for baseline demographics and risk factors** | | | | | | | | |
| --- | --- | --- | --- | --- | --- | --- | --- | --- | --- |
| **Characteristics ᵃ** | COVID-19 with anosmia | COVID-19 without anosmia |  | COVID-19 after prior vaccination | COVID-19 without prior vaccination |  | Influenza after prior vaccination | Influenza without prior vaccination |  |
|  | No. (%) |  | SMD | No. (%) |  | SMD | No. (%) |  | SMD |
| **Number** | 12207 | 12207 | - | 18101 | 18101 | - | 87936 | 87936 | - |
| **Age; mean (SD); y** | 44.7 (16.2) | 44.8 (16.2) | 0.005 | 52.2 (17.7) | 51.6 (17.4) | 0.04 | 50.1 (17.8) | 49.7 (17.4) | 0.02 |
| **Sex** |  |  |  |  |  |  |  |  |  |
| Female | 8055 (66.0) | 8043 (65.9) | 0.002 | 11560 (63.9) | 11796 (65.2) | 0.03 | 57084 (64.9) | 57704 (65.6) | 0.01 |
| Male | 4146 (34.0) | 4157 (34.1) | 0.002 | 6539 (36.1) | 6304 (34.8) | 0.03 | 30847 (35.1) | 30224 (34.4) | 0.01 |
| **Race** |  |  |  |  |  |  |  |  |  |
| White | 8010 (65.6) | 8122 (66.5) | 0.02 | 13188 (72.9) | 13453 (74.3) | 0.03 | 65620 (74.6) | 65462 (74.4) | 0.004 |
| Black or African American | 2389 (19.6) | 2423 (19.8) | 0.007 | 3246 (17.9) | 2995 (16.5) | 0.04 | 11698 (13.3) | 11980 (13.6) | 0.009 |
| **Ethnicity** |  |  |  |  |  |  |  |  |  |
| Hispanic or Latino | 1320 (10.8) | 1245 (10.2) | 0.02 | 2558 (14.1) | 2455 (13.6) | 0.02 | 7681 (8.7) | 7757 (8.8) | 0.003 |
| Not Hispanic of Latino | 8483 (69.5) | 8534 (69.9) | 0.009 | 14961 (82.7) | 15075 (83.3) | 0.02 | 71113 (80.9) | 71309 (81.1) | 0.006 |
| Unknown | 2404 (19.7) | 2428 (19.9) | 0.005 | 582 (3.2) | 571 (3.2) | 0.003 | 9142 (10.4) | 8870 (10.1) | 0.01 |
| **Comorbidities** |  |  |  |  |  |  |  |  |  |
| Human immunodeficiency virus disease | 320 (2.6) | 311 (2.5) | 0.005 | 845 (4.7) | 769 (4.2) | 0.02 | 1665 (1.9) | 849 (1.0) | 0.08 |
| Neoplasms | 4475 (36.7) | 4410 (36.1) | 0.01 | 9761 (53.9) | 9894 (54.7) | 0.01 | 36061 (41.0) | 35893 (40.8) | 0.004 |
| Diabetes mellitus | 2422 (19.8) | 2333 (19.1) | 0.02 | 6097 (33.7) | 6053 (33.4) | 0.005 | 20303 (23.1) | 20153 (22.9) | 0.004 |
| Overweight and obesity | 4149 (34.0) | 4131 (33.8) | 0.003 | 7550 (41.7) | 7587 (41.9) | 0.004 | 27916 (31.7) | 28251 (32.1) | 0.008 |
| Unspecified dementia | 97 (0.8) | 83 (0.7) | 0.01 | 490 (2.7) | 438 (2.4) | 0.02 | 852 (1.0) | 737 (0.8) | 0.01 |
| Nicotine dependence | 2460 (20.2) | 2500 (20.5) | 0.008 | 4700 (26.0) | 4610 (25.5) | 0.01 | 15458 (17.6) | 15574 (17.7) | 0.003 |
| Paraplegia and quadriplegia | 54 (0.4) | 72 (0.6) | 0.02 | 159 (0.9) | 153 (0.8) | 0.004 | 458 (0.5) | 277 (0.3) | 0.03 |
| Essential hypertension | 4663 (38.2) | 4657 (38.1) | 0.001 | 10024 (55.4) | 9971 (55.1) | 0.006 | 42509 (48.3) | 42823 (48.7) | 0.007 |

#### **Supplementary Table 13 (continued)**: Baseline characteristics for COVID-19 patients with anosmia or prior vaccination compared to without anosmia or prior vaccination, after matching.

| **Characteristics ᵃ** | COVID-19 with anosmia | COVID-19 without anosmia |  | COVID-19 after prior vaccination | COVID-19 without prior vaccination |  | Influenza after prior vaccination | Influenza without prior vaccination |  |
| --- | --- | --- | --- | --- | --- | --- | --- | --- | --- |
|  | No. (%) |  | SMD | No. (%) |  | SMD | No. (%) |  | SMD |
| Ischemic heart diseases | 1369 (11.2) | 1310 (10.7) | 0.02 | 4946 (27.3) | 4972 (27.5) | 0.003 | 13332 (15.2) | 12749 (14.5) | 0.02 |
| Acute myocardial infarction | 431 (3.5) | 388 (3.2) | 0.02 | 1906 (10.5) | 1654 (9.1) | 0.05 | 3034 (3.5) | 2923 (3.3) | 0.007 |
| Heart failure | 655 (5.4) | 747 (6.1) | 0.03 | 3218 (17.8) | 2733 (15.1) | 0.07 | 7082 (8.1) | 7043 (8.0) | 0.002 |
| Heart disease | 420 (3.4) | 346 (2.8) | 0.03 | 1818 (10.0) | 1764 ( 9.7) | 0.01 | 2740 (3.1) | 2321 (2.6) | 0.03 |
| Cerebral infarction | 628 (5.1) | 530 (4.3) | 0.04 | 2576 (14.2) | 2501 (13.8) | 0.01 | 2884 (3.3) | 2542 (2.9) | 0.02 |
| Peripheral vascular disease | 394 (3.2) | 393 (3.2) | 5E-04 | 1876 (10.4) | 1571 (8.7) | 0.06 | 3766 (4.3) | 3245 (3.7) | 0.03 |
| Chronic lower respiratory diseases | 3518 (28.8) | 3484 (28.5) | 0.006 | 6782 (37.5) | 6804 (37.6) | 0.003 | 35820 (40.7) | 36056 (41.0) | 0.005 |
| Chronic obstructive pulmonary disease | 592 (4.8) | 658 (5.4) | 0.02 | 2249 (12.4) | 2253 (12.4) | 7E-04 | 8696 ( 9.9) | 8884 (10.1) | 0.007 |
| Asthma | 2367 (19.4) | 2347 (19.2) | 0.004 | 4283 (23.7) | 4412 (24.4) | 0.02 | 22052 (25.1) | 22498 (25.6) | 0.01 |
| Peptic ulcer | 142 (1.2) | 138 (1.1) | 0.003 | 612 (3.4) | 415 (2.3) | 0.07 | 1400 (1.6) | 1133 (1.3) | 0.03 |
| Fibrosis and cirrhosis of liver | 187 (1.5) | 145 (1.2) | 0.03 | 815 (4.5) | 756 (4.2) | 0.02 | 1334 (1.5) | 1179 (1.3) | 0.01 |
| Portal hypertension | 52 (0.4) | 65 (0.5) | 0.02 | 351 (1.9) | 295 (1.6) | 0.02 | 503 (0.6) | 432 (0.5) | 0.01 |
| Psoriasis | 282 (2.3) | 271 (2.2) | 0.006 | 645 (3.6) | 574 (3.2) | 0.02 | 2266 (2.6) | 2146 (2.4) | 0.009 |
| Lupus erythematosus | 89 (0.7) | 69 (0.6) | 0.02 | 252 (1.4) | 218 (1.2) | 0.02 | 449 (0.5) | 381 (0.4) | 0.01 |
| Rheumatoid arthritis without rheumatoid factor | 24 (0.2) | 17 (0.1) | 0.01 | 48 (0.3) | 38 (0.2) | 0.01 | 148 (0.2) | 116 (0.1) | 0.009 |
| Rheumatoid arthritis | 225 (1.8) | 201 (1.6) | 0.02 | 700 (3.9) | 564 (3.1) | 0.04 | 2437 (2.8) | 2255 (2.6) | 0.01 |
| Acute kidney failure | 806 (6.6) | 835 (6.8) | 0.009 | 3284 (18.1) | 2580 (14.3) | 0.1 | 6704 (7.6) | 6153 (7.0) | 0.02 |
| Chronic kidney disease | 880 (7.2) | 810 (6.6) | 0.02 | 3329 (18.4) | 3325 (18.4) | 6E-04 | 8338 (9.5) | 7719 (8.8) | 0.02 |
| End stage renal disease | 162 (1.3) | 232 (1.9) | 0.05 | 724 (4.0) | 709 (3.9) | 0.004 | 1505 (1.7) | 1859 (2.1) | 0.03 |

Abbreviations: SMD: standardized mean difference

ᵃ Patients were identified using *International Classification of Diseases and Related Health Problems, Tenth Revision* (*ICD-10*) and *Current Procedural Terminology (CPT)* codes. Characteristics were identified using electronic medical health record data from the TriNetX Research Network, and were recorded any time prior to the index event. Diagnostic coding can be found in the **online supplementary methods**.

#### **Supplementary Table 14**. Sensitivity analyses. ORs and (excess) rates of new-onset outcomes between 3 months and one year after COVID-19 with prior vaccination compared to without, and influenza with prior vaccination compared to without.

|  | **Influenza with prior vaccination vs without prior vaccination** | | | | | **COVID-19 with prior vaccination vs without prior vaccination** | | | | |
| --- | --- | --- | --- | --- | --- | --- | --- | --- | --- | --- |
|  | OR (95% CI) | No. Cases / No. Subjects (%) |  | P-value |  | OR (95% CI) | No. Cases / No. Subjects (%) |  | P-value |  |
| **Outcomeᵃ** |  | Influenza with vaccination | Influenza without vaccination | Uncorrected | Corrected |  | COVID-19 with vaccination | COVID-19 without vaccination | Uncorrected | Corrected |
| **Gastrointestinal symptoms or diagnoses** | 1.00 (0.95 - 1.05) | 2,703 / 22,703 (11,91) | 3,869 / 32,438 (11,93) | 0.94 | 1 | 0.55 (0.47 - 0.64) | 258 / 3,833 (6.73) | 556 / 4,772 (11.65) | <.0001 | <.0001 |
| Dysphagia | 1.11 (1.02 - 1.21) | 1,145 / 78,922 (1,45) | 1,060 / 81,065 (1,31) | 0.01 | 0.21 | 0.68 (0.57 - 0.82) | 186 / 14,456 (1.29) | 282 / 15,062 (1.87) | <.0001 | 0.0004 |
| GERD | 1.02 (0.96 - 1.08) | 2,287 / 52,803 (4,33) | 2,533 / 59,686 (4,24) | 0.47 | 1 | 0.63 (0.55 - 0.73) | 305 / 10,137 (3.01) | 505 / 10,817 (4.67) | <.0001 | <.0001 |
| Heartburn | 1.24 (1.09 - 1.42) | 494 / 83,715 (,59) | 405 / 85,335 (,47) | 1 | 0.02 | 0.59 (0.45 - 0.77) | 82 / 16,119 (.51) | 141 / 16,369 (.86) | 0.0001 | 0.0008 |
| Nausea | 1.06 (0.99 - 1.13) | 1,942 / 69,688 (2,79) | 1,962 / 74,519 (2,63) | 0.07 | 0.75 | 0.65 (0.56 - 0.76) | 277 / 11,215 (2.47) | 461 / 12,316 (3.74) | <.0001 | <.0001 |
| Vomiting | 1.05 (0.96 - 1.16) | 839 / 79,293 (1,06) | 822 / 81,710 (1,01) | 0.3 | 1 | 0.48 (0.39 - 0.58) | 138 / 12,921 (1.07) | 310 / 13,958 (2.22) | <.0001 | <.0001 |
| Early satiety | 1.08 (0.83 - 1.42) | 109 / 87,340 (,12) | 101 / 87,522 (,12) | 0.57 | 1 | 0.91 (0.58 - 1.42) | 37 / 17,655 (.21) | 41 / 17,825 (.23) | 0.68 | 1 |
| Abdominal and pelvic pain | 0.97 (0.93 - 1.02) | 3,111 / 42,221 (7,37) | 3,930 / 51,927 (7,57) | 0.25 | 1 | 0.65 (0.56 - 0.74) | 331 / 6,826 (4.85) | 595 / 8,148 (7.3) | <.0001 | <.0001 |
| Bloating | 1.19 (1.08 - 1.30) | 928 / 81,346 (1,14) | 807 / 83,679 (,96) | 0.0004 | 0.01 | 0.45 (0.37 - 0.55) | 143 / 15,016 (.95) | 326 / 15,702 (2.08) | <.0001 | <.0001 |
| Gastroparesis | 1.08 (0.86 - 1.35) | 161 / 86,606 (,19) | 150 / 86,957 (,17) | 0.51 | 1 | 0.96 (0.62 - 1.47) | 41 / 17,635 (.23) | 43 / 17,681 (.24) | 0.84 | 1 |
| Functional dyspepsia | 1.08 (0.93 - 1.26) | 335 / 83,960 (,4) | 315 / 85,505 (,37) | 0.31 | 1 | 0.76 (0.55 - 1.05) | 63 / 16,846 (.37) | 84 / 17,083 (.49) | 0.1 | 0.55 |
| Constipation | 1.08 (1.01 - 1.15) | 1,922 / 69,429 (2,77) | 1,929 / 74,814 (2,58) | 0.03 | 0.31 | 0.71 (0.61 - 0.82) | 286 / 11,502 (2.49) | 435 / 12,476 (3.49) | <.0001 | <.0001 |
| Diarrhea | 1.07 (1.01 - 1.14) | 2,199 / 66,026 (3,33) | 2,235 / 71,768 (3,11) | 0.02 | 0.3 | 0.71 (0.61 - 0.83) | 279 / 10,852 (2.57) | 432 / 12,107 (3.57) | <.0001 | 0.0001 |
| Irritable bowel syndrome | 1.15 (1.01 - 1.31) | 513 / 82,106 (,62) | 455 / 83,778 (,54) | 0.03 | 0.34 | 0.85 (0.61 - 1.18) | 66 / 16,869 (.39) | 78 / 16,992 (.46) | 0.34 | 1 |
| Inflammatory bowel disease | 1.03 (0.95 - 1.12) | 1,087 / 75,581 (1,44) | 1,111 / 79,408 (1,4) | 0.52 | 1 | 0.87 (0.71 - 1.07) | 170 / 15,252 (1.11) | 201 / 15,719 (1.28) | 0.18 | 0.97 |

#### **Supplementary Table 14 (continued)**. Sensitivity analyses. ORs and (excess) rates of new-onset outcomes between 3 months and one year after COVID-19 with prior vaccination compared to without, and influenza with prior vaccination compared to without.

|  | **Influenza with prior vaccination vs without prior vaccination** | | | | | **COVID-19 with prior vaccination vs without prior vaccination** | | | | |
| --- | --- | --- | --- | --- | --- | --- | --- | --- | --- | --- |
|  | OR (95% CI) | No. Cases / No. Subjects (%) |  | P-value |  | OR (95% CI) | No. Cases / No. Subjects (%) |  | P-value |  |
| **Outcome ᵃ** |  | Influenza with vaccination | Influenza without vaccination | Uncorrected | Corrected |  | COVID-19 with vaccination | COVID-19 without vaccination | Uncorrected | Corrected |
| **Autonomic neuropathy** | 1.12 (1.06 - 1.17) | 3,509 / 37,242 (9,42) | 4,095 / 48,024 (8,53) | <.0001 | 0.0004 | 0.73 (0.64 - 0.84) | 378 / 5,400 (7.) | 641 / 6,870 (9.33) | <.0001 | <.0001 |
| Postural symptoms | 1.07 (1.02 - 1.13) | 3,165 / 50,982 (6,21) | 3,392 / 58,479 (5,8) | 5 | 0.08 | 0.71 (0.62 - 0.81) | 361 / 7,202 (5.01) | 617 / 8,876 (6.95) | <.0001 | <.0001 |
| Urinary dysfunction | 1.19 (1.12 - 1.26) | 2,234 / 66,232 (3,37) | 2,098 / 73,648 (2,85) | <.0001 | <.0001 | 0.92 (0.79 - 1.06) | 339 / 12,317 (2.75) | 392 / 13,094 (2.99) | 0.25 | 1 |
| Sexual dysfunction | 1.24 (1.10 - 1.40) | 607 / 81,247 (,75) | 506 / 83,914 (,6) | 0.0003 | 0.01 | 0.71 (0.55 - 0.93) | 93 / 16,291 (.57) | 133 / 16,681 (.8) | 0.01 | 0.08 |
| Exocrine gland dysfunction | 1.21 (1.11 - 1.31) | 1,313 / 77,281 (1,7) | 1,143 / 81,001 (1,41) | <.0001 | 0.0004 | 0.85 (0.72 - 1.00) | 262 / 12,978 (2.02) | 337 / 14,177 (2.38) | 0.04 | 0.26 |
| **Sensory neuropathy** | 1.12 (1.05 - 1.21) | 1,591 / 72,374 (2,2) | 1,521 / 77,535 (1,96) | 1 | 0.02 | 0.64 (0.54 - 0.76) | 221 / 12,957 (1.71) | 361 / 13,662 (2.64) | <.0001 | <.0001 |
| **Motor neuropathy** | 1.16 (1.08 - 1.24) | 1,602 / 75,584 (2,12) | 1,450 / 78,955 (1,84) | <.0001 | 3 | 0.93 (0.79 - 1.10) | 264 / 13,073 (2.02) | 314 / 14,519 (2.16) | 0.41 | 1 |
| **Drugs for autonomic neuropathy** | 0.89 (0.84 - 0.95) | 1,895 / 60,485 (3,13) | 2,213 / 63,142 (3,5) | 0.0003 | 9 | 0.51 (0.44 - 0.61) | 225 / 9,835 (2.29) | 451 / 10,364 (4.35) | <.0001 | <.0001 |
| Fludrocortisone | 0.93 (0.65 - 1.34) | 56 / 87,363 (,06) | 60 / 87,464 (,07) | 0.71 | 1 | 0.90 (0.47 - 1.72) | 17 / 17,925 (.09) | 19 / 17,942 (.11) | 0.74 | 1 |
| Beta-blockers | 0.89 (0.83 - 0.95) | 1,873 / 60,827 (3,08) | 2,191 / 63,448 (3,45) | 0.0002 | 8 | 0.51 (0.43 - 0.60) | 220 / 9,922 (2.22) | 444 / 10,449 (4.25) | <.0001 | <.0001 |
| Pyridostigmine | 1.50 (0.67 - 3.34) | 15 / 87,719 (,02) | 10 / 87,719 (,01) | 0.32 | 1 | 1.00 (0.42 - 2.41) | 10 / 18,038 (.06) | 10 / 18,059 (.06) | 1 | 1 |
| Midodrine | 0.80 (0.62 - 1.03) | 104 / 87,395 (,12) | 130 / 87,366 (,15) | 0.09 | 0.88 | 1.54 (1.11 - 2.14) | 90 / 17,606 (.51) | 59 / 17,763 (.33) | 0.009 | 0.06 |

Abbreviations: OR: odds ratio

ᵃ Patients and outcomes were identified using *International Classification of Diseases and Related Health Problems, Tenth Revision* (*ICD-10*) and *Current Procedural Terminology (CPT)* codes. Diagnostic coding can be found in the **online supplementary methods**.

#### **Supplementary Table 15**. ORs and (excess) rates of new-onset outcomes between 3 months and one year after COVID-19 with anosmia compared to without.

|  | **COVID-19 with anosmia vs without anosmia** | | | | |
| --- | --- | --- | --- | --- | --- |
|  | OR (95% CI) | No. Cases / No. Subjects (%) |  | P-value |  |
| **Outcomeᵃ** |  | COVID-19 with anosmia | COVID-19 without anosmia | Uncorrected | Corrected |
| **Gastrointestinal symptoms or diagnoses** | 0.99 (0.77 - 1.27) | 114 / 3,374 (3.38) | 142 / 4,144 (3.43) | 0.91 | 1 |
| Dysphagia | 0.91 (0.61 - 1.36) | 46 / 10,929 (.42) | 51 / 11,040 (.46) | 0.65 | 1 |
| GERD | 0.95 (0.73 - 1.23) | 112 / 8,398 (1.33) | 119 / 8,493 (1.4) | 0.71 | 1 |
| Heartburn | 0.48 (0.27 - 0.85) | 17 / 11,437 (.15) | 36 / 11,593 (.31) | 0.01 | 0.54 |
| Nausea | 1.07 (0.78 - 1.47) | 74 / 8,511 (.87) | 77 / 9,451 (.81) | 0.69 | 1 |
| Vomiting | 0.82 (0.49 - 1.35) | 27 / 9,919 (.27) | 35 / 10,507 (.33) | 0.43 | 1 |
| Early satiety | 1.00 (0.42 - 2.41) | 10 / 12,078 (.08) | 10 / 12,113 (.08) | 0.99 | 1 |
| Abdominal and pelvic pain | 1.00 (0.79 - 1.26) | 136 / 5,719 (2.38) | 151 / 6,325 (2.39) | 0.97 | 1 |
| Bloating | 1.07 (0.70 - 1.63) | 45 / 10,963 (.41) | 43 / 11,197 (.38) | 0.75 | 1 |
| Gastroparesis | 0.91 (0.38 - 2.13) | 10 / 12,095 (.08) | 11 / 12,059 (.09) | 0.82 | 1 |
| Functional dyspepsia | 0.67 (0.30 - 1.49) | 10 / 11,747 (.09) | 15 / 11,833 (.13) | 0.33 | 1 |
| Constipation | 1.25 (0.90 - 1.73) | 79 / 9,264 (.85) | 66 / 9,622 (.69) | 0.19 | 1 |
| Diarrhea | 0.97 (0.72 - 1.30) | 80 / 8,057 (.99) | 96 / 9,340 (1.03) | 0.82 | 1 |
| Irritable bowel syndrome | 1.00 (0.54 - 1.86) | 20 / 11,581 (.17) | 20 / 11,582 (.17) | 1 | 1 |
| Inflammatory bowel disease | 1.48 (0.97 - 2.23) | 55 / 10,816 (.51) | 38 / 11,008 (.35) | 0.06 | 1 |
| **Autonomic neuropathy** | 0.93 (0.75 - 1.17) | 141 / 5,192 (2.72) | 176 / 6,065 (2.9) | 0.55 | 1 |
| Postural symptoms | 0.87 (0.69 - 1.11) | 121 / 6,685 (1.81) | 154 / 7,457 (2.07) | 0.27 | 1 |
| Urinary dysfunction | 0.84 (0.65 - 1.09) | 104 / 9,452 (1.1) | 129 / 9,869 (1.31) | 0.19 | 1 |
| Sexual dysfunction | 0.80 (0.48 - 1.33) | 27 / 11,463 (.24) | 34 / 11,601 (.29) | 0.39 | 1 |
| Exocrine gland dysfunction | 1.36 (0.95 - 1.93) | 70 / 10,083 (.69) | 55 / 10,726 (.51) | 0.09 | 1 |
| **Sensory neuropathy** | 1.31 (0.94 - 1.82) | 79 / 9,681 (.82) | 64 / 10,250 (.62) | 0.11 | 1 |
| **Motor neuropathy** | 1.12 (0.81 - 1.56) | 75 / 10,457 (.72) | 68 / 10,635 (.64) | 0.49 | 1 |
| **Drugs for autonomic neuropathy** | 0.81 (0.63 - 1.04) | 113 / 8,986 (1.26) | 134 / 8,608 (1.56) | 0.09 | 1 |
| Fludrocortisone | 1.00 (0.42 - 2.40) | 10 / 12,162 (.08) | 10 / 12,147 (.08) | 1 | 1 |
| Beta-blockers | 0.82 (0.64 - 1.06) | 113 / 9,017 (1.25) | 132 / 8,653 (1.53) | 0.12 | 1 |
| Pyridostigmine | - | 0 / 12207 (0) | 10 / 12,193 (.08) | 0.002 | 0.16 |
| Midodrine | 1.00 (0.42 - 2.40) | 10 / 12,127 (.08) | 10 / 12,100 (.08) | 1 | 1 |

Abbreviations: OR: odds ratio

ᵃ Patients and outcomes were identified using *International Classification of Diseases and Related Health Problems, Tenth Revision* (*ICD-10*) and *Current Procedural Terminology (CPT)* codes. Diagnostic coding can be found in the **online supplementary methods**.

#### **Supplementary Table 16**. ORs of new-onset outcomes between 3 months and one year after COVID-19, compared to historic and contemporary negative controls.

|  | **Negative controls (February)** |  |  | **Negative controls (Juli)** |  |  | **Negative controls (September)** |  |  | **Negative controls (November)** |  |  |
| --- | --- | --- | --- | --- | --- | --- | --- | --- | --- | --- | --- | --- |
|  | OR (95% CI) | P-value |  | OR (95% CI) | P-value |  | OR (95% CI) | P-value |  | OR (95% CI) | P-value |  |
| **Outcomeᵃ** |  | Uncorrected | Corrected |  | Uncorrected | Corrected |  | Uncorrected | Corrected |  | Uncorrected | Corrected |
| **Gastrointestinal symptoms or diagnoses** | 1.53 (1.51 - 1.56) | <.0001 | <.0001 | 1.36 (1.33 - 1.38) | <.0001 | <.0001 | 1.36 (1.33 - 1.38) | <.0001 | <.0001 | 1.26 (1.24 - 1.28) | <.0001 | <.0001 |
| Dysphagia | 1.88 (1.81 - 1.95) | <.0001 | <.0001 | 1.56 (1.51 - 1.61) | <.0001 | <.0001 | 1.70 (1.65 - 1.76) | <.0001 | <.0001 | 1.57 (1.52 - 1.62) | <.0001 | <.0001 |
| GERD | 1.41 (1.38 - 1.44) | <.0001 | <.0001 | 1.30 (1.27 - 1.33) | <.0001 | <.0001 | 1.33 (1.30 - 1.36) | <.0001 | <.0001 | 1.24 (1.21 - 1.27) | <.0001 | <.0001 |
| Heartburn | 1.96 (1.86 - 2.06) | <.0001 | <.0001 | 1.42 (1.36 - 1.49) | <.0001 | <.0001 | 2.02 (1.93 - 2.11) | <.0001 | <.0001 | 1.77 (1.69 - 1.85) | <.0001 | <.0001 |
| Nausea | 2.27 (2.20 - 2.33) | <.0001 | <.0001 | 1.80 (1.75 - 1.84) | <.0001 | <.0001 | 2.03 (1.98 - 2.08) | <.0001 | <.0001 | 1.76 (1.72 - 1.81) | <.0001 | <.0001 |
| Vomiting | 3.37 (3.23 - 3.51) | <.0001 | <.0001 | 1.99 (1.92 - 2.06) | <.0001 | <.0001 | 2.52 (2.43 - 2.61) | <.0001 | <.0001 | 2.20 (2.12 - 2.27) | <.0001 | <.0001 |
| Early satiety | 1.75 (1.57 - 1.94) | <.0001 | <.0001 | 1.62 (1.46 - 1.79) | <.0001 | <.0001 | 1.61 (1.46 - 1.78) | <.0001 | <.0001 | 1.28 (1.17 - 1.41) | <.0001 | <.0001 |
| Abdominal and pelvic pain | 1.69 (1.66 - 1.72) | <.0001 | <.0001 | 1.46 (1.43 - 1.48) | <.0001 | <.0001 | 1.49 (1.46 - 1.52) | <.0001 | <.0001 | 1.36 (1.33 - 1.39) | <.0001 | <.0001 |
| Bloating | 2.23 (2.15 - 2.32) | <.0001 | <.0001 | 1.72 (1.66 - 1.78) | <.0001 | <.0001 | 1.99 (1.93 - 2.06) | <.0001 | <.0001 | 1.79 (1.74 - 1.85) | <.0001 | <.0001 |
| Gastroparesis | 1.50 (1.37 - 1.65) | <.0001 | <.0001 | 1.36 (1.24 - 1.49) | <.0001 | <.0001 | 1.51 (1.37 - 1.66) | <.0001 | <.0001 | 1.39 (1.27 - 1.53) | <.0001 | <.0001 |
| Functional dyspepsia | 1.92 (1.78 - 2.06) | <.0001 | <.0001 | 1.56 (1.46 - 1.67) | <.0001 | <.0001 | 2.06 (1.93 - 2.20) | <.0001 | <.0001 | 1.76 (1.66 - 1.87) | <.0001 | <.0001 |
| Constipation | 1.83 (1.78 - 1.88) | <.0001 | <.0001 | 1.47 (1.44 - 1.51) | <.0001 | <.0001 | 1.65 (1.61 - 1.69) | <.0001 | <.0001 | 1.51 (1.48 - 1.55) | <.0001 | <.0001 |
| Diarrhea | 2.04 (1.99 - 2.09) | <.0001 | <.0001 | 1.74 (1.69 - 1.78) | <.0001 | <.0001 | 1.87 (1.82 - 1.91) | <.0001 | <.0001 | 1.69 (1.65 - 1.73) | <.0001 | <.0001 |
| Irritable bowel syndrome | 1.24 (1.17 - 1.31) | <.0001 | <.0001 | 1.05 (1.00 - 1.11) | 0.05 | 0.2 | 1.15 (1.10 - 1.21) | <.0001 | <.0001 | 1.06 (1.01 - 1.11) | 0.02 | 0.09 |
| Inflammatory bowel disease | 1.72 (1.65 - 1.79) | <.0001 | <.0001 | 1.56 (1.49 - 1.62) | <.0001 | <.0001 | 1.58 (1.51 - 1.64) | <.0001 | <.0001 | 1.48 (1.42 - 1.54) | <.0001 | <.0001 |

#### **Supplementary Table 16 (continued)**. ORs of new-onset outcomes between 3 months and one year after COVID-19, compared to historic and contemporary negative controls.

|  | **Negative controls (February)** |  |  | **Negative controls (Juli)** |  |  | **Negative controls (September)** |  |  | **Negative controls (November)** |  |  |
| --- | --- | --- | --- | --- | --- | --- | --- | --- | --- | --- | --- | --- |
|  | OR (95% CI) | P-value |  | OR (95% CI) | P-value |  | OR (95% CI) | P-value |  | OR (95% CI) | P-value |  |
| **Outcome ᵃ** |  | Uncorrected | Corrected |  | Uncorrected | Corrected |  | Uncorrected | Corrected |  | Uncorrected | Corrected |
| **Autonomic neuropathy** | 1.57 (1.55 - 1.60) | <.0001 | <.0001 | 1.34 (1.31 - 1.36) | <.0001 | <.0001 | 1.37 (1.34 - 1.39) | <.0001 | <.0001 | 1.23 (1.21 - 1.25) | <.0001 | <.0001 |
| Postural symptoms | 1.75 (1.71 - 1.78) | <.0001 | <.0001 | 1.43 (1.40 - 1.45) | <.0001 | <.0001 | 1.49 (1.46 - 1.52) | <.0001 | <.0001 | 1.35 (1.32 - 1.37) | <.0001 | <.0001 |
| Urinary dysfunction | 1.46 (1.43 - 1.50) | <.0001 | <.0001 | 1.32 (1.29 - 1.35) | <.0001 | <.0001 | 1.40 (1.36 - 1.43) | <.0001 | <.0001 | 1.26 (1.23 - 1.29) | <.0001 | <.0001 |
| Sexual dysfunction | 1.28 (1.22 - 1.34) | <.0001 | <.0001 | 1.12 (1.07 - 1.17) | <.0001 | <.0001 | 1.19 (1.14 - 1.24) | <.0001 | <.0001 | 1.10 (1.06 - 1.15) | <.0001 | <.0001 |
| Exocrine gland dysfunction | 2.07 (2.00 - 2.14) | <.0001 | <.0001 | 1.58 (1.54 - 1.63) | <.0001 | <.0001 | 1.83 (1.78 - 1.89) | <.0001 | <.0001 | 1.58 (1.53 - 1.63) | <.0001 | <.0001 |
| **Sensory neuropathy** | 1.65 (1.61 - 1.70) | <.0001 | <.0001 | 1.35 (1.31 - 1.39) | <.0001 | <.0001 | 1.55 (1.51 - 1.60) | <.0001 | <.0001 | 1.43 (1.40 - 1.47) | <.0001 | <.0001 |
| **Motor neuropathy** | 1.59 (1.54 - 1.65) | <.0001 | <.0001 | 1.32 (1.28 - 1.36) | <.0001 | <.0001 | 1.52 (1.48 - 1.57) | <.0001 | <.0001 | 1.32 (1.28 - 1.36) | <.0001 | <.0001 |
| **Drugs for autonomic neuropathy** | 1.27 (1.24 - 1.30) | <.0001 | <.0001 | 1.17 (1.15 - 1.20) | <.0001 | <.0001 | 1.17 (1.14 - 1.19) | <.0001 | <.0001 | 1.09 (1.07 - 1.11) | <.0001 | <.0001 |
| Fludrocortisone | 1.79 (1.55 - 2.08) | <.0001 | <.0001 | 1.58 (1.37 - 1.82) | <.0001 | <.0001 | 1.47 (1.28 - 1.69) | <.0001 | <.0001 | 1.43 (1.24 - 1.65) | <.0001 | <.0001 |
| Beta-blockers | 1.26 (1.23 - 1.29) | <.0001 | <.0001 | 1.17 (1.14 - 1.19) | <.0001 | <.0001 | 1.17 (1.14 - 1.19) | <.0001 | <.0001 | 1.08 (1.06 - 1.11) | <.0001 | <.0001 |
| Pyridostigmine | 1.23 (0.96 - 1.57) | 0.1 | 0.4 | 0.99 (0.79 - 1.26) | 0.95 | 1 | 1.14 (0.89 - 1.46) | 0.29 | 1 | 0.95 (0.75 - 1.20) | 0.68 | 1 |
| Midodrine | 2.20 (2.02 - 2.40) | <.0001 | <.0001 | 1.97 (1.81 - 2.14) | <.0001 | <.0001 | 1.71 (1.57 - 1.85) | <.0001 | <.0001 | 1.71 (1.58 - 1.85) | <.0001 | <.0001 |

Abbreviations: OR: odds ratio

ᵃ Patients and outcomes were identified using *International Classification of Diseases and Related Health Problems, Tenth Revision* (*ICD-10*) and *Current Procedural Terminology (CPT)* codes. Diagnostic coding can be found in the **online supplementary methods**.

#### **Supplementary Table 17**. Rates of new-onset outcomes between 3 months and one year after COVID-19, compared to historic and contemporary negative controls.

|  | **Negative controls (February)** |  | **Negative controls (Juli)** |  | **Negative controls (September)** |  | **Negative controls (November)** |  |
| --- | --- | --- | --- | --- | --- | --- | --- | --- |
|  | No. Cases / No. Subjects (%) |  | No. Cases / No. Subjects (%) |  | No. Cases / No. Subjects (%) |  | No. Cases / No. Subjects (%) |  |
| **Outcome ᵃ** | COVID-19 | Negative controls | COVID-19 | Negative controls | COVID-19 | Negative controls | COVID-19 | Negative controls |
| **Gastrointestinal symptoms or diagnoses** | 26,961 / 253,855 (10.62) | 21,995 / 305,855 (7.19) | 27,231 / 256,456 (10.62) | 24,332 / 302,502 (8.04) | 26,589 / 251,470 (10.57) | 23,940 / 298,548 (8.02) | 26,588 / 251,446 (10.57) | 25,393 / 295,585 (8.59) |
| Dysphagia | 8,531 / 584,713 (1.46) | 4,715 / 602,614 (.78) | 8,614 / 588,637 (1.46) | 5,702 / 603,911 (.94) | 9,382 / 584,329 (1.61) | 5,721 / 603,412 (.95) | 9,377 / 584,206 (1.61) | 6,210 / 602,100 (1.03) |
| GERD | 18,169 / 462,664 (3.93) | 13,661 / 483,759 (2.82) | 18,181 / 462,599 (3.93) | 14,664 / 480,401 (3.05) | 18,404 / 462,922 (3.98) | 14,444 / 479,577 (3.01) | 18,403 / 462,812 (3.98) | 15,487 / 479,714 (3.23) |
| Heartburn | 4,070 / 616,802 (.66) | 2,121 / 627,382 (.34) | 4,100 / 621,109 (.66) | 2,923 / 628,359 (.47) | 5,366 / 615,862 (.87) | 2,725 / 628,289 (.43) | 5,366 / 615,731 (.87) | 3,093 / 626,463 (.49) |
| Nausea | 15,344 / 518,865 (2.96) | 7,519 / 566,606 (1.33) | 15,588 / 525,402 (2.97) | 9,509 / 568,523 (1.67) | 15,784 / 513,164 (3.08) | 8,725 / 566,369 (1.54) | 15,782 / 513,083 (3.08) | 9,926 / 560,968 (1.77) |
| Vomiting | 9,107 / 564,277 (1.61) | 2,917 / 601,867 (.48) | 9,209 / 572,507 (1.61) | 4,938 / 605,519 (.82) | 10,142 / 557,556 (1.82) | 4,408 / 603,212 (.73) | 10,141 / 557,477 (1.82) | 5,007 / 598,578 (.84) |
| Early satiety | 978 / 644,375 (.15) | 562 / 646,152 (.09) | 981 / 644,193 (.15) | 607 / 645,722 (.09) | 993 / 647,041 (.15) | 617 / 648,526 (.1) | 993 / 646,888 (.15) | 776 / 647,962 (.12) |
| Abdominal and pelvic pain | 24,346 / 378,612 (6.43) | 16,819 / 430,162 (3.91) | 24,608 / 382,689 (6.43) | 19,342 / 428,862 (4.51) | 24,436 / 375,252 (6.51) | 19,050 / 425,791 (4.47) | 24,435 / 375,217 (6.51) | 20,609 / 422,736 (4.88) |
| Bloating | 8,805 / 600,721 (1.47) | 4,054 / 612,551 (.66) | 8,870 / 604,175 (1.47) | 5,250 / 612,352 (.86) | 10,157 / 600,160 (1.69) | 5,246 / 612,247 (.86) | 10,156 / 600,035 (1.69) | 5,809 / 610,787 (.95) |
| gastroparesis | 1,051 / 642,274 (.16) | 701 / 643,706 (.11) | 1,054 / 641,927 (.16) | 778 / 643,372 (.12) | 1,103 / 644,871 (.17) | 733 / 645,906 (.11) | 1,100 / 644,720 (.17) | 791 / 645,968 (.12) |
| Functional dyspepsia | 2,163 / 631,071 (.34) | 1,137 / 634,685 (.18) | 2,169 / 631,406 (.34) | 1,400 / 634,871 (.22) | 2,807 / 631,996 (.44) | 1,376 / 636,908 (.22) | 2,807 / 631,855 (.44) | 1,610 / 636,700 (.25) |
| Constipation | 15,296 / 524,628 (2.92) | 8,973 / 555,910 (1.61) | 15,399 / 528,863 (2.91) | 11,078 / 555,487 (1.99) | 16,255 / 522,220 (3.11) | 10,624 / 554,813 (1.91) | 16,248 / 522,136 (3.11) | 11,419 / 549,422 (2.08) |
| Diarrhea | 15,702 / 509,933 (3.08) | 8,559 / 558,104 (1.53) | 15,908 / 514,351 (3.09) | 10,066 / 558,169 (1.8) | 16,319 / 504,117 (3.24) | 9,802 / 556,702 (1.76) | 16,318 / 504,037 (3.24) | 10,734 / 552,333 (1.94) |
| Irritable bowel syndrome | 3,008 / 622,304 (.48) | 2,437 / 623,507 (.39) | 3,006 / 622,124 (.48) | 2,858 / 622,537 (.46) | 3,135 / 623,924 (.5) | 2,722 / 624,462 (.44) | 3,134 / 623,775 (.5) | 2,962 / 624,785 (.47) |
| Inflammatory bowel disease | 5,905 / 594,114 (.99) | 3,523 / 605,869 (.58) | 5,914 / 594,010 (1.) | 3,889 / 605,688 (.64) | 6,156 / 595,756 (1.03) | 3,999 / 607,660 (.66) | 6,156 / 595,628 (1.03) | 4,267 / 608,087 (.7) |

#### **Supplementary Table 17 (continued)**. Rates of new-onset outcomes between 3 months and one year after COVID-19, compared to historic and contemporary negative controls.

|  | **Negative controls (February)** |  | **Negative controls (Juli)** |  | **Negative controls (September)** |  | **Negative controls (November)** |  |
| --- | --- | --- | --- | --- | --- | --- | --- | --- |
|  | No. Cases / No. Subjects (%) |  | No. Cases / No. Subjects (%) |  | No. Cases / No. Subjects (%) |  | No. Cases / No. Subjects (%) |  |
| **Outcome ᵃ** | COVID-19 | Negative controls | COVID-19 | Negative controls | COVID-19 | Negative controls | COVID-19 | Negative controls |
| **Autonomic neuropathy** | 29,198 / 343,440 (8.5) | 21,131 / 379,085 (5.57) | 29,503 / 346,915 (8.5) | 24,394 / 374,772 (6.51) | 28,678 / 339,690 (8.44) | 23,494 / 371,479 (6.32) | 28,673 / 339,644 (8.44) | 25,523 / 366,682 (6.96) |
| Postural symptoms | 25,080 / 413,111 (6.07) | 16,256 / 455,288 (3.57) | 25,400 / 418,279 (6.07) | 19,682 / 453,666 (4.34) | 24,760 / 408,235 (6.07) | 18,668 / 450,084 (4.15) | 24,757 / 408,179 (6.07) | 20,387 / 445,964 (4.57) |
| Urinary dysfunction | 14,572 / 534,545 (2.73) | 10,340 / 550,480 (1.88) | 14,650 / 536,156 (2.73) | 11,430 / 548,359 (2.08) | 15,201 / 533,887 (2.85) | 11,286 / 548,690 (2.06) | 15,198 / 533,778 (2.85) | 12,454 / 545,943 (2.28) |
| Sexual dysfunction | 4,208 / 615,623 (.68) | 3,305 / 616,262 (.54) | 4,211 / 615,922 (.68) | 3,765 / 614,667 (.61) | 4,370 / 617,705 (.71) | 3,679 / 617,077 (.6) | 4,369 / 617,557 (.71) | 3,958 / 616,604 (.64) |
| Exocrine gland dysfunction | 10,688 / 569,897 (1.88) | 5,416 / 591,267 (.92) | 10,805 / 575,551 (1.88) | 7,055 / 591,333 (1.19) | 11,884 / 568,207 (2.09) | 6,811 / 591,047 (1.15) | 11,881 / 568,092 (2.09) | 7,807 / 585,171 (1.33) |
| **Sensory neuropathy** | 12,054 / 553,393 (2.18) | 7,552 / 568,817 (1.33) | 12,172 / 557,197 (2.18) | 9,252 / 568,570 (1.63) | 13,286 / 550,551 (2.41) | 8,903 / 568,499 (1.57) | 13,286 / 550,445 (2.41) | 9,608 / 566,785 (1.7) |
| **Motor neuropathy** | 9,762 / 571,863 (1.71) | 6,335 / 587,767 (1.08) | 9,814 / 573,809 (1.71) | 7,672 / 588,485 (1.3) | 10,784 / 570,150 (1.89) | 7,360 / 588,125 (1.25) | 10,781 / 570,029 (1.89) | 8,421 / 585,114 (1.44) |
| **Drugs for autonomic neuropathy** | 18,541 / 447,455 (4.14) | 15,198 / 460,957 (3.3) | 18,537 / 447,128 (4.15) | 16,299 / 458,752 (3.55) | 18,402 / 450,293 (4.09) | 16,192 / 460,507 (3.52) | 18,398 / 450,158 (4.09) | 17,276 / 458,869 (3.76) |
| Fludrocortisone | 494 / 646,398 (.08) | 276 / 646,835 (.04) | 494 / 646,002 (.08) | 313 / 646,412 (.05) | 473 / 649,017 (.07) | 322 / 649,367 (.05) | 473 / 648,866 (.07) | 331 / 649,110 (.05) |
| Beta-blockers | 18,326 / 450,014 (4.07) | 15,083 / 463,227 (3.26) | 18,323 / 449,681 (4.07) | 16,182 / 461,069 (3.51) | 18,180 / 452,816 (4.01) | 16,034 / 462,757 (3.46) | 18,176 / 452,680 (4.02) | 17,124 / 461,232 (3.71) |
| Pyridostigmine | 140 / 648,635 (.02) | 114 / 648,626 (.02) | 139 / 648,237 (.02) | 140 / 648,282 (.02) | 138 / 651,205 (.02) | 121 / 651,194 (.02) | 138 / 651,051 (.02) | 145 / 651,013 (.02) |
| Midodrine | 1,638 / 643,250 (.25) | 749 / 645,663 (.12) | 1,639 / 642,857 (.25) | 838 / 645,077 (.13) | 1,618 / 645,917 (.25) | 953 / 647,997 (.15) | 1,617 / 645,764 (.25) | 951 / 647,783 (.15) |

ᵃ Patients and outcomes were identified using *International Classification of Diseases and Related Health Problems, Tenth Revision* (*ICD-10*) and *Current Procedural Terminology (CPT)* codes. Diagnostic coding can be found in the **online supplementary methods**.

**Supplementary Table 18.** ORs of new-onset outcomes between 1 year and 2 years after COVID-19, compared to contemporary negative controls and influenza.

|  | **Contemporary negative controls** |  |  | **Influenza** |  |  |
| --- | --- | --- | --- | --- | --- | --- |
|  | OR (95% CI) | P-value |  | OR (95% CI) | P-value |  |
| **Outcome ᵃ** |  | Uncorrected | Corrected |  | Uncorrected | Corrected |
| **Gastrointestinal symptoms or diagnoses** | 1.42 (1.37 - 1.47) | <.0001 | <.0001 | 1.05 (1.01 - 1.08) | 0.006 | 0.03 |
| Dysphagia | 1.47 (1.40 - 1.54) | <.0001 | <.0001 | 0.87 (0.84 - 0.91) | <.0001 | <.0001 |
| GERD | 1.39 (1.35 - 1.44) | <.0001 | <.0001 | 1.08 (1.04 - 1.11) | <.0001 | <.0001 |
| Heartburn | 1.23 (1.15 - 1.31) | <.0001 | <.0001 | 0.58 (0.55 - 0.62) | <.0001 | <.0001 |
| Nausea | 1.72 (1.65 - 1.79) | <.0001 | <.0001 | 0.86 (0.83 - 0.89) | <.0001 | <.0001 |
| Vomiting | 1.80 (1.71 - 1.89) | <.0001 | <.0001 | 0.67 (0.64 - 0.70) | <.0001 | <.0001 |
| Early satiety | 1.28 (1.13 - 1.45) | 0.0001 | 0.0006 | 1.03 (0.92 - 1.16) | 0.61 | 1 |
| Abdominal and pelvic pain | 1.40 (1.35 - 1.44) | <.0001 | <.0001 | 0.90 (0.88 - 0.93) | <.0001 | <.0001 |
| Bloating | 1.55 (1.48 - 1.63) | <.0001 | <.0001 | 0.91 (0.87 - 0.95) | <.0001 | <.0001 |
| Gastroparesis | 1.65 (1.45 - 1.88) | <.0001 | <.0001 | 1.09 (0.97 - 1.22) | 0.16 | 0.7 |
| Functional dyspepsia | 1.37 (1.25 - 1.49) | <.0001 | <.0001 | 0.61 (0.57 - 0.66) | <.0001 | <.0001 |
| Constipation | 1.38 (1.33 - 1.44) | <.0001 | <.0001 | 0.90 (0.87 - 0.93) | <.0001 | <.0001 |
| Diarrhea | 1.69 (1.63 - 1.75) | <.0001 | <.0001 | 0.85 (0.82 - 0.88) | <.0001 | <.0001 |
| Irritable bowel syndrome | 1.10 (1.02 - 1.18) | 0.01 | 0.05 | 0.92 (0.85 - 0.98) | 0.02 | 0.08 |
| Inflammatory bowel disease | 1.59 (1.50 - 1.69) | <.0001 | <.0001 | 0.76 (0.73 - 0.80) | <.0001 | <.0001 |
| **Autonomic neuropathy** | 1.37 (1.33 - 1.41) | <.0001 | <.0001 | 1.06 (1.03 - 1.09) | 0.0003 | 0.002 |
| Postural symptoms | 1.46 (1.41 - 1.50) | <.0001 | <.0001 | 0.98 (0.95 - 1.01) | 0.22 | 0.95 |
| Urinary dysfunction | 1.30 (1.26 - 1.35) | <.0001 | <.0001 | 0.99 (0.95 - 1.02) | 0.46 | 1 |
| Sexual dysfunction | 1.10 (1.03 - 1.17) | 0.004 | 0.02 | 0.94 (0.88 - 0.99) | 0.03 | 0.15 |
| Exocrine gland dysfunction | 1.44 (1.38 - 1.50) | <.0001 | <.0001 | 0.83 (0.80 - 0.87) | <.0001 | <.0001 |
| **Sensory neuropathy** | 1.25 (1.20 - 1.30) | <.0001 | <.0001 | 0.68 (0.65 - 0.70) | <.0001 | <.0001 |
| **Motor neuropathy** | 1.32 (1.27 - 1.38) | <.0001 | <.0001 | 0.73 (0.70 - 0.76) | <.0001 | <.0001 |
| **Drugs for autonomic neuropathy** | 1.26 (1.22 - 1.31) | <.0001 | <.0001 | 1.34 (1.29 - 1.39) | <.0001 | <.0001 |
| Fludrocortisone | 1.69 (1.37 - 2.09) | <.0001 | <.0001 | 1.24 (1.02 - 1.50) | 0.03 | 0.15 |
| Beta-blockers | 1.26 (1.22 - 1.30) | <.0001 | <.0001 | 1.33 (1.29 - 1.38) | <.0001 | <.0001 |
| Pyridostigmine | 1.24 (0.90 - 1.72) | 0.19 | 0.73 | 1.95 (1.35 - 2.84) | 0.0003 | 0.002 |
| Midodrine | 2.34 (2.07 - 2.66) | <.0001 | <.0001 | 1.91 (1.70 - 2.15) | <.0001 | <.0001 |

Abbreviations: OR: odds ratio

ᵃ Patients and outcomes were identified using *International Classification of Diseases and Related Health Problems, Tenth Revision* (*ICD-10*) and *Current Procedural Terminology (CPT)* codes. Diagnostic coding can be found in the **online supplementary methods**.

#### **Supplementary Table 19**. ORs of new-onset outcomes between 1 year and 2 years after COVID-19, compared to other infectious diseases.

|  | **Lyme’s disease** |  |  | **Herpes Zoster** |  |  | **Varicella Zoster** |  |  | **Infectious mononucleosis** |  |  | **Cytomegaloviral disease** |  |  |
| --- | --- | --- | --- | --- | --- | --- | --- | --- | --- | --- | --- | --- | --- | --- | --- |
|  | OR (95% CI) | P-value |  | OR (95% CI) | P-value |  | OR (95% CI) | P-value |  | OR (95% CI) | P-value |  | OR (95% CI) | P-value |  |
| **Outcomeᵃ** |  | Uncorrected | Corrected |  | Uncorrected | Corrected |  | Uncorrected | Corrected |  | Uncorrected | Corrected |  | Uncorrected | Corrected |
| **Gastrointestinal symptoms or diagnoses** | 1.73 (1.66 – 1.81) | <.0001 | <.0001 | 1.25 (1.21 – 1.29) | <.0001 | <.0001 | 1.64 (1.56 – 1.72) | <.0001 | <.0001 | 1.41 (1.32 – 1.50) | <.0001 | <.0001 | 0.99 (0.70 – 1.41) | 0.96 | 1 |
| Dysphagia | 1.42 (1.33 – 1.53) | <.0001 | <.0001 | 1.06 (1.02 – 1.11) | 0.008 | 0.04 | 0.81 (0.76 – 0.87) | <.0001 | <.0001 | 1.19 (1.05 – 1.34) | 0.005 | 0.03 | 0.97 (0.72 – 1.30) | 0.82 | 1 |
| GERD | 1.59 (1.52 – 1.67) | <.0001 | <.0001 | 1.16 (1.12 – 1.20) | <.0001 | <.0001 | 1.53 (1.45 – 1.61) | <.0001 | <.0001 | 1.36 (1.26 – 1.47) | <.0001 | <.0001 | 1.00 (0.77 – 1.30) | 0.99 | 1 |
| Heartburn | 1.17 (1.06 – 1.30) | 0.002 | 0.009 | 0.80 (0.76 – 0.86) | <.0001 | <.0001 | 0.44 (0.41 – 0.48) | <.0001 | <.0001 | 1.10 (0.94 – 1.28) | 0.23 | 0.99 | 0.90 (0.60 – 1.35) | 0.61 | 1 |
| Nausea | 1.74 (1.65 – 1.84) | <.0001 | <.0001 | 1.18 (1.13 – 1.22) | <.0001 | <.0001 | 0.84 (0.80 – 0.89) | <.0001 | <.0001 | 1.44 (1.33 – 1.56) | <.0001 | <.0001 | 1.07 (0.82 – 1.38) | 0.62 | 1 |
| Vomiting | 1.61 (1.49 – 1.73) | <.0001 | <.0001 | 0.98 (0.94 – 1.03) | 0.45 | 1 | 0.45 (0.42 – 0.48) | <.0001 | <.0001 | 1.37 (1.22 – 1.53) | <.0001 | <.0001 | 1.11 (0.82 – 1.51) | 0.5 | 1 |
| Early satiety | 1.85 (1.48 – 2.31) | <.0001 | <.0001 | 1.23 (1.08 – 1.41) | 0.001 | 0.008 | 0.85 (0.70 – 1.02) | 0.08 | 0.33 | 1.58 (1.14 – 2.18) | 0.005 | 0.03 | 0.47 (0.22 – 1.01) | 0.05 | 0.61 |
| Abdominal and pelvic pain | 1.44 (1.38 – 1.51) | <.0001 | <.0001 | 1.11 (1.07 – 1.14) | <.0001 | <.0001 | 1.20 (1.15 – 1.26) | <.0001 | <.0001 | 1.27 (1.19 – 1.36) | <.0001 | <.0001 | 1.14 (0.90 – 1.46) | 0.27 | 1 |
| Bloating | 1.66 (1.54 – 1.78) | <.0001 | <.0001 | 1.24 (1.19 – 1.30) | <.0001 | <.0001 | 0.88 (0.82 – 0.94) | <.0001 | 0.0005 | 1.41 (1.26 – 1.58) | <.0001 | <.0001 | 1.48 (1.08 – 2.02) | 0.01 | 0.38 |
| Gastroparesis | 1.34 (1.08 – 1.65) | 0.007 | 0.03 | 1.04 (0.92 – 1.17) | 0.57 | 1 | 1.45 (1.17 – 1.79) | 0.0007 | 0.003 | 1.24 (0.92 – 1.69) | 0.16 | 0.74 | 0.80 (0.42 – 1.51) | 0.48 | 1 |
| Functional dyspepsia | 1.22 (1.08 – 1.39) | 0.002 | 0.01 | 0.81 (0.74 – 0.88) | <.0001 | <.0001 | 0.50 (0.45 – 0.56) | <.0001 | <.0001 | 1.12 (0.92 – 1.37) | 0.25 | 1 | 0.78 (0.46 – 1.32) | 0.35 | 1 |
| Constipation | 1.66 (1.57 – 1.75) | <.0001 | <.0001 | 1.14 (1.10 – 1.19) | <.0001 | <.0001 | 0.83 (0.79 – 0.88) | <.0001 | <.0001 | 1.38 (1.26 – 1.50) | <.0001 | <.0001 | 1.09 (0.82 – 1.44) | 0.55 | 1 |
| Diarrhea | 1.55 (1.48 – 1.64) | <.0001 | <.0001 | 1.11 (1.07 – 1.14) | <.0001 | <.0001 | 0.85 (0.80 – 0.89) | <.0001 | <.0001 | 1.24 (1.14 – 1.34) | <.0001 | <.0001 | 0.74 (0.57 – 0.96) | 0.02 | 0.44 |
| Irritable bowel syndrome | 0.98 (0.89 – 1.09) | 0.75 | 1 | 0.96 (0.89 – 1.04) | 0.3 | 1 | 1.25 (1.12 – 1.40) | <.0001 | 0.0003 | 1.02 (0.88 – 1.17) | 0.83 | 1 | 1.59 (0.93 – 2.73) | 0.09 | 0.93 |
| Inflammatory bowel disease | 1.36 (1.26 – 1.48) | <.0001 | <.0001 | 0.91 (0.86 – 0.96) | 0.0002 | 0.001 | 1.11 (1.02 – 1.21) | 0.01 | 0.06 | 0.96 (0.85 – 1.08) | 0.47 | 1 | 0.74 (0.55 – 1.01) | 0.06 | 0.67 |

#### **Supplementary Table 19 (continued)**. ORs of new-onset outcomes between 1 year and 2 years after COVID-19, compared to other infectious diseases.

|  | **Lyme’s disease** |  |  | **Herpes Zoster** |  |  | **Varicella Zoster** |  |  | **Infectious mononucleosis** |  |  | **Cytomegaloviral disease** |  |  |
| --- | --- | --- | --- | --- | --- | --- | --- | --- | --- | --- | --- | --- | --- | --- | --- |
|  | OR (95% CI) | P-value |  | OR (95% CI) | P-value |  | OR (95% CI) | P-value |  | OR (95% CI) | P-value |  | OR (95% CI) | P-value |  |
| **Outcome ᵃ** |  | Uncorrected | Corrected |  | Uncorrected | Corrected |  | Uncorrected | Corrected |  | Uncorrected | Corrected |  | Uncorrected | Corrected |
| **Autonomic neuropathy** | 1.49 (1.43 - 1.55) | <.0001 | <.0001 | 1.18 (1.14 - 1.21) | <.0001 | <.0001 | 1.26 (1.21 - 1.32) | <.0001 | <.0001 | 1.35 (1.27 - 1.43) | <.0001 | <.0001 | 1.51 (1.18 - 1.92) | 0.0009 | 0.04 |
| Postural symptoms | 1.42 (1.36 - 1.48) | <.0001 | <.0001 | 1.21 (1.17 - 1.24) | <.0001 | <.0001 | 1.10 (1.05 - 1.15) | <.0001 | 0.0003 | 1.34 (1.25 - 1.43) | <.0001 | <.0001 | 1.76 (1.39 - 2.23) | <.0001 | 0.0003 |
| Urinary dysfunction | 1.31 (1.25 - 1.38) | <.0001 | <.0001 | 1.10 (1.06 - 1.14) | <.0001 | <.0001 | 1.11 (1.05 - 1.17) | 0.0002 | 0.001 | 1.27 (1.16 - 1.38) | <.0001 | <.0001 | 1.20 (0.93 - 1.53) | 0.16 | 1 |
| Sexual dysfunction | 1.29 (1.17 - 1.41) | <.0001 | <.0001 | 1.02 (0.96 - 1.09) | 0.54 | 1 | 1.12 (1.01 - 1.25) | 0.03 | 0.13 | 1.21 (1.01 - 1.45) | 0.04 | 0.18 | 0.94 (0.66 - 1.34) | 0.75 | 1 |
| Exocrine gland dysfunction | 1.54 (1.44 - 1.63) | <.0001 | <.0001 | 1.07 (1.03 - 1.12) | 0.0007 | 0.004 | 0.63 (0.59 - 0.66) | <.0001 | <.0001 | 1.20 (1.08 - 1.33) | 0.0004 | 0.003 | 1.25 (0.95 - 1.65) | 0.11 | 1 |
| **Sensory neuropathy** | 1.07 (1.01 - 1.14) | 0.02 | 0.07 | 0.85 (0.81 - 0.88) | <.0001 | <.0001 | 0.59 (0.56 - 0.62) | <.0001 | <.0001 | 1.15 (1.04 - 1.26) | 0.004 | 0.03 | 1.52 (1.13 - 2.05) | 0.005 | 0.18 |
| **Motor neuropathy** | 1.05 (0.99 - 1.12) | 0.11 | 0.47 | 0.88 (0.85 - 0.92) | <.0001 | <.0001 | 0.70 (0.66 - 0.74) | <.0001 | <.0001 | 1.18 (1.06 - 1.31) | 0.002 | 0.01 | 1.43 (1.04 - 1.95) | 0.03 | 0.44 |
| **Drugs for autonomic neuropathy** | 1.72 (1.63 - 1.80) | <.0001 | <.0001 | 1.36 (1.31 - 1.40) | <.0001 | <.0001 | 1.96 (1.85 - 2.08) | <.0001 | <.0001 | 1.76 (1.62 - 1.92) | <.0001 | <.0001 | 1.11 (0.81 - 1.51) | 0.52 | 1 |
| Fludrocortisone | 0.95 (0.71 - 1.26) | 0.71 | 1 | 1.04 (0.85 - 1.26) | 0.72 | 1 | 1.13 (0.85 - 1.52) | 0.4 | 1 | 0.59 (0.40 - 0.87) | 0.008 | 0.04 | 0.46 (0.22 - 0.99) | 0.04 | 0.61 |
| Beta-blockers | 1.74 (1.65 - 1.83) | <.0001 | <.0001 | 1.35 (1.31 - 1.40) | <.0001 | <.0001 | 1.97 (1.86 - 2.09) | <.0001 | <.0001 | 1.81 (1.66 - 1.97) | <.0001 | <.0001 | 1.12 (0.83 - 1.51) | 0.45 | 1 |
| Pyridostigmine | 1.10 (0.71 - 1.70) | 0.66 | 1 | 1.72 (1.20 - 2.47) | 0.003 | 0.02 | 2.30 (1.30 - 4.06) | 0.003 | 0.01 | 1.84 (0.91 - 3.71) | 0.09 | 0.41 | 1.00 (0.42 - 2.41) | 1 | 1 |
| Midodrine | 2.69 (2.15 - 3.36) | <.0001 | <.0001 | 1.90 (1.68 - 2.15) | <.0001 | <.0001 | 2.04 (1.63 - 2.55) | <.0001 | <.0001 | 1.71 (1.23 - 2.39) | 0.001 | 0.009 | 0.98 (0.60 - 1.61) | 0.93 | 1 |

Abbreviations: OR: odds ratio

ᵃ Patients and outcomes were identified using *International Classification of Diseases and Related Health Problems, Tenth Revision* (*ICD-10*) and *Current Procedural Terminology (CPT)* codes. Diagnostic coding can be found in the **online supplementary methods**.
